# Machine learning applications in vascular neuroimaging for the diagnosis and prognosis of cognitive impairment and dementia: a systematic review and meta-analysis

**DOI:** 10.1101/2024.12.17.24319166

**Authors:** Valerie Lohner, Amanpreet Badhwar, Flavie E. Detcheverry, Cindy L. García, Helena M. Gellersen, Zahra Khodakarami, René Lattmann, Rui Li, Audrey Low, Claudia Mazo, Amelie Metz, Olivier Parent, Veronica Phillips, Usman Saeed, Sean YW Tan, Stefano Tamburin, David J. Llewellyn, Timothy Rittman, Sheena Waters, Jose Bernal

**Author notes:** Corresponding author: Dr Valerie Lohner, Klinik III für Innere Medizin, Universitätsklinikum Köln (AöR) Kerpener Str. 62, 50937 Cologne, Germany, Mail.

## Abstract

**Introduction:** Machine learning (ML) algorithms using neuroimaging markers of cerebral small vessel disease (CSVD) are a promising approach for classifying cognitive impairment and dementia.

**Methods:** We systematically reviewed and meta-analysed studies that leveraged CSVD features for ML-based diagnosis and/or prognosis of cognitive impairment and dementia.

**Results:** We identified 75 relevant studies: 43 on diagnosis, 27 on prognosis, and 5 on both. CSVD markers are becoming important in ML-based classifications of neurodegenerative diseases, mainly Alzheimer’s dementia, with nearly 60% of studies published in the last two years. Regression and support vector machine techniques were more common than other approaches such as ensemble and deep-learning algorithms. ML-based classification performed well for both Alzheimer’s dementia (AUC 0.88 [95%-CI 0.85–0.92]) and cognitive impairment (AUC 0.84 [95%-CI 0.74–0.95]). Of 75 studies, only 16 were suitable for meta-analysis, only 11 used multiple datasets for training and validation, and six lacked clear definitions of diagnostic criteria.

**Discussion:** ML-based models using CSVD neuroimaging markers perform well in classifying cognitive impairment and dementia. However, challenges in inconsistent reporting, limited generalisability, and potential biases hinder adoption. Our targeted recommendations provide a roadmap to accelerate the integration of ML into clinical practice.

## I. INTRODUCTION

Cerebral small vessel disease (CSVD) describes multiple dynamic pathological processes that impair the optimal functioning of perforating arterioles, capillaries, and venules in the brain [1–3]. CSVD is among the most common conditions encountered by neurologists in clinical practice [4] and a significant contributor to major healthcare challenges. CSVD causes 25% of ischaemic strokes, the majority of intracerebral haemorrhages in individuals over 65 years old, and most cases of vascular dementia [2, 3]; contributes to around 45% of all dementia cases worldwide [4]; and leads to mobility and gait issues, neurobehavioural changes, and mood disorders [5]. The relationship between CSVD and Alzheimer’s disease (AD) has been recognised since the earliest days of AD research [6], and is now included in the Alzheimer’s Society’s most recent revised criteria for diagnosing and staging AD [7]. This coexistence between CSVD and AD has taken on a new significance in recent years, as anti-amyloid monoclonal antibody trials have revealed that individuals with cerebral amyloid angiopathy — a form of CSVD — are at risk of developing brain swelling or haemorrhages during the course of the treatment, making CSVD assessments and studies crucial for patient stratification and minimising treatment risks [8].

Although direct assessment of the human cerebral microvasculature *in vivo* remains challenging with standard imaging technologies, its chronic dysfunction leads to changes that can be detected through magnetic resonance imaging (MRI) and computed tomography (**Supplementary BOX 1**). Assessing CSVD has traditionally focused on evaluating discrete lesions, such as white matter hyperintensities (WMH), lacunes, cerebral microbleeds, superficial siderosis, perivascular spaces, and small subcortical or cortical microinfarcts, by means of clinical visual ratings and increasingly through quantitative methods [1]. However, advancements in neuroimaging technologies have also revealed that these discrete lesions are not the only consequence, suggesting instead that they often lead to widespread, rather than focal, alterations of microstructure and connectivity [9].

The integration of neuroimaging and machine learning (ML) techniques (**Supplementary BOX 2**) presents new avenues for understanding the intricate and multifactorial nature of CSVD and vascular contributors to cognitive impairment and dementia [10]. These possibilities include not only the computational quantification of neuroimaging markers of CSVD (e.g., through segmentation of lesions) [11–13] but also their predictive value for neurodegenerative diseases and dementia, which could ultimately facilitate early detection and personalised treatments. However, the contribution of CSVD to dementia and cognitive impairment using ML appears to be underdeveloped. According to a recent systematic review and meta-analysis conducted by the Imaging Working Group of the international DEMON Network on the application of neuroimaging and ML for dementia diagnosis and prognosis [14], only 2 out of 255 studies focused on vascular forms of dementia. Whilst that review did not consider cognitive changes other than dementia and most included studies leveraged the Alzheimer’s Disease Neuroimaging Initiative (ADNI)—a cohort with relatively minimal CSVD burden—it is surprising so few studies had been conducted in this field.

To map the significance of CSVD in ML-based detection of dementia and cognitive impairment more generally, we established a new subgroup of the DEMON Imaging Working group dedicated specifically to this topic. We conducted a systematic review and meta-analysis to (a) determine the use of CSVD neuroimaging markers and ML in the diagnosis and prognosis of cognitive impairment and dementia; (b) identify methodological shifts over time, particularly with recent deep learning advancements; and (c) pinpoint methodological barriers preventing the development and effective deployment of these strategies. Our primary focus was on papers addressing dementia-related diagnosis and prognosis rather than those solely centred on lesion segmentation. We aim for this review to inspire the development of more accurate and validated methods for predicting CSVD-related cognitive impairment, facilitating early detection and intervention.

## II. METHODS

### 2.1 Protocol registration

We registered this systematic review protocol with the International Prospective Register of Systematic Reviews (PROSPERO), registration number: CRD42022366767. We conducted this work following the Preferred Reporting Items for Systematic Reviews and Meta-Analysis (PRISMA) Statement [15]. The associated PRISMA checklist can be found in the supplementary material.

### 2.2 Search strategy

A medical librarian (VP) searched databases Medline (via Ovid), Embase (via Ovid), Cochrane Library, Emcare (via Ovid), Cinahl (via Ebscohost), PsycInfo (via Ebscohost), BNI (via ProQuest), Web of Science (Core Collection), and Scopus from inception to the date searches were conducted. The search strategy was peer-reviewed using the Peer Review of Electronic Search Strategies (PRESS) checklist [16], and evaluated against the PRISMA-S guidelines [17]. Databases were searched separately, rather than multiple databases being searched on the same platform. The search syntax was adapted for each database, and to account for variation between thesaurus terms/controlled vocabulary across each database. Results were limited to the English language in all databases. Results were exported to Endnote 20 for deduplication, using the method outlined by Bramer et al. [18].

All searches were originally conducted on September 20, 2023 and rerun on September 9, 2024 to include any papers published between the initial search and final submission.

### 2.3 PICOS framework

The parameters of this systematic review, as defined by the PICOS framework, were as follows:

● <u>P</u>articipants: Persons with cognitive impairment or a clinical diagnosis of dementia, as well as people with incident cognitive impairment or dementia.
● Index: Neuroimaging-derived CSVD data analysed with ML for diagnosis or prognosis.
● <u>C</u>omparator:
  ○ For diagnostic studies: persons without cognitive impairment or dementia.
  ○ For prognostic studies: prognostic factor (conversion to cognitive impairment or dementia vs no conversion).
● <u>O</u>utcome: Accuracy of diagnosis or prognosis of cognitive impairment or dementia based on CSVD burden.
● <u>S</u>tudy design: Original cross-sectional or prospective observational studies.

### 2.4 Inclusion and exclusion criteria

To be included, studies had to report on the model performance of the ML methods for the diagnosis or prognosis of cognitive impairment or dementia using imaging markers of CSVD and ML. We deemed eligible original studies published in English in peer-reviewed journals and excluded *in vitro* studies or animal studies. We also excluded studies that employed ML solely for image processing such segmentation.

### 2.5 Study selection

Study selection had two stages. First, each report was screened for eligibility by pairs of independent reviewers based on title and abstract using the screening tool Rayyan (https://www.rayyan.ai/). Second, each report that passed the initial filtering was reviewed by pairs of reviewers who independently conducted full-text screening. Conflicts arising at any of these two stages were resolved through discussions, with the assistance of a third independent senior reviewer when necessary.

### 2.6 Data extraction

Data from each included study were extracted independently by pairs of reviewers using a standard template. Once again, conflicts were resolved through discussion, with a third senior reviewer solving any remaining disagreements. The data extraction form captured (a) article information (first/last author, year, journal, country of first/last author’s affiliated institution, study type); (b) characteristics of the study population (sample size, age, sex, ethnicity, criteria for cognitive impairment and dementia, inclusion and exclusion criteria of study); (c) data analysis (ML approach used, covariates included in model, vascular neuroimaging features used; outcome measure); (d) results (measures of model performance (accuracy, sensitivity, specificity, area under the curve (AUC), positive predictive value, negative predictive value), other metrics reported (e.g. hazard or odds ratios), follow-up period (for prognostic studies only); and (e) risk of bias assessment.

### 2.7 Assessment of risk of bias

We assessed the quality of all individual studies using the QUality Assessment of Diagnostic Accuracy Studies (QUADAS-2) for diagnostic studies [19] and the Prediction model Risk Of Bias ASsessment Tool (PROBAST) for prognostic studies [20]. Pairs of reviewers independently conducted the critical appraisal of each paper and certainty of evidence rating. Disagreements were resolved through discussion.

### 2.8 Meta-analysis

We conducted a meta-analysis to provide a targeted evaluation of the ML models using vascular neuroimaging features. This meta-analysis focused on the two most common diagnostic tasks identified in the literature we reviewed: distinguishing between healthy controls and AD-dementia or all-cause dementia, as well as between healthy controls and cognitive impairment. Thus, we compared the performance of the various approaches using the AUC obtained from Receiver Operating Characteristic analyses.

We employed a random-effects model with the DerSimonian and Laird estimation method to calculate the pooled AUC values and confidence intervals. In cases of missing data, such as absent variability measures for the AUC, we reached out to the corresponding authors. In instances where authors did not respond to our inquiries and studies failed to report any measures of variability for the AUC, we estimated the standard error for the AUC based on Hanley and McNeil [21]. If different ML approaches were considered for the same database, we included the analysis utilising the ML model with the best model fit and the largest sample size. We quantified heterogeneity using Cohen’s Q statistics and I^2^ statistics. The meta-analysis was performed in R version 4.2.1 [22] using the package *metafor* [23].

## III. RESULTS

### 3.1 Search results

Our initial search on September 20, 2023, identified 4,956 potentially relevant records across all databases (**Figure 1**). After deduplication using Endnote and Rayyan, we retained 2,630 records. Of these, 256 passed the title and abstract screening, and 62 went on to pass full-text screening and were included in the systematic review [24–85]. On September 9, 2024, we conducted a rerun and identified 845 potentially new relevant records across the same databases (**Figure 1**). A total of 546 records remained after deduplication. Screening by title and abstract led to 35 eligible papers, of which 13 passed full-text screening and were included in the systematic review [86–98]. This brought the total number of included records to 75 [24–98]. We contacted the corresponding authors of 36 studies via email to request missing data. Of these, six responded: three provided additional data, while the other three were unable to do so. Finally, a total of 16 studies was suitable for meta-analysis [30, 32, 33, 47, 51, 58, 65, 79, 84, 85, 89, 91, 96, 98], of which seven classified healthy controls versus AD-dementia [30, 32, 47, 58, 65, 84, 89], and nine healthy controls versus cognitive impairment [32, 33, 51, 58, 79, 85, 91, 96, 98]. A flow chart of the identification and screening process is provided in **Figure 1**.

**Figure 1.**
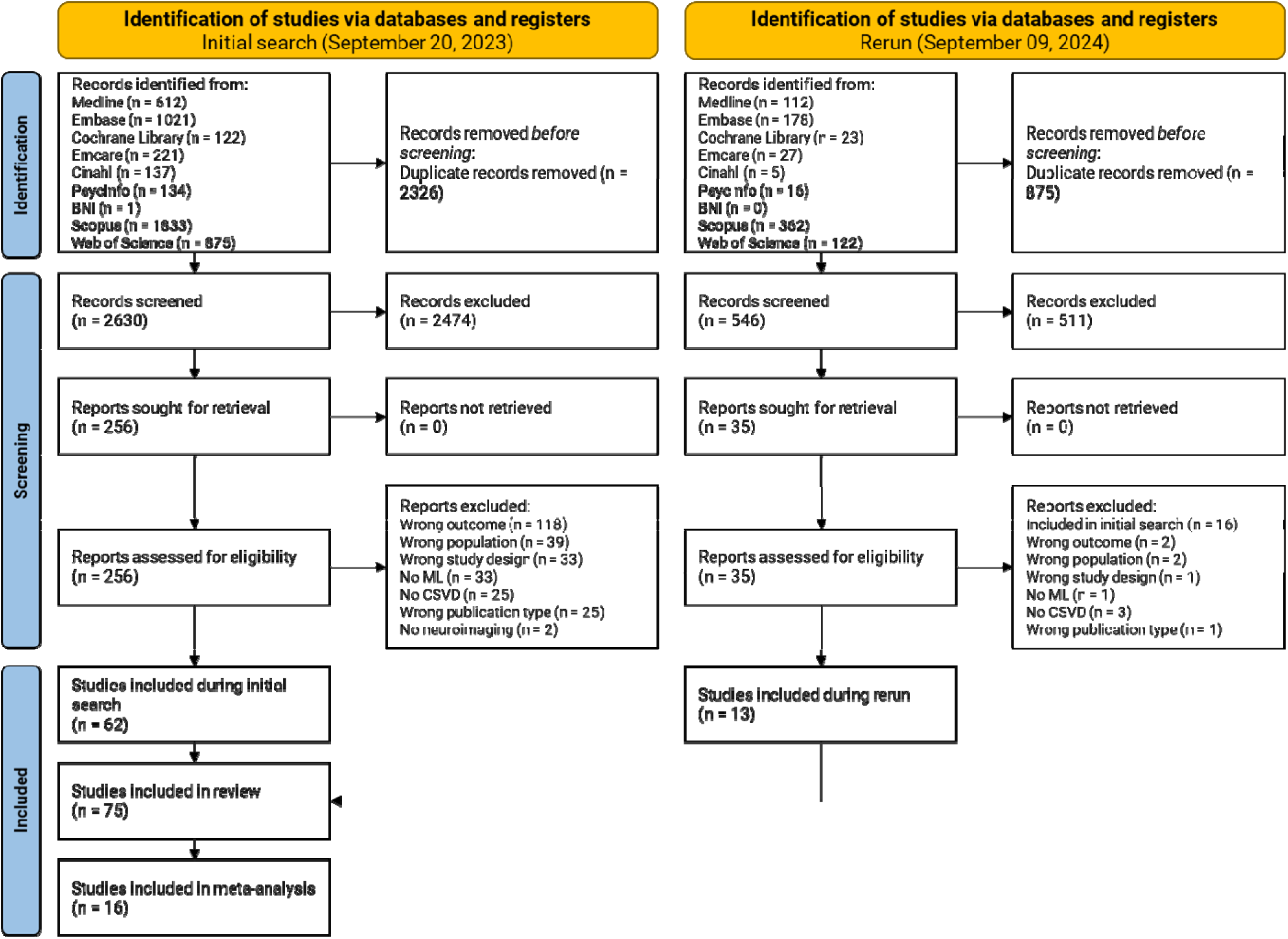
PRISMA flow chart outlining the number of studies identified, included, and excluded at each stage of the systematic review and meta-analysis.

### 3.2 Study characteristics

#### 3.2.1 Origin of studies

According to the affiliations of the first and last authors, the majority of included studies were from China (n=24) and the USA (n=16). The remaining studies originated primarily from Europe (n=33), followed by Asia (n=7, excluding China), and North America (n=3, excluding the USA). No studies were affiliated with institutions in South America, Africa, or Australia (**Figure 2**).

**Figure 2.**
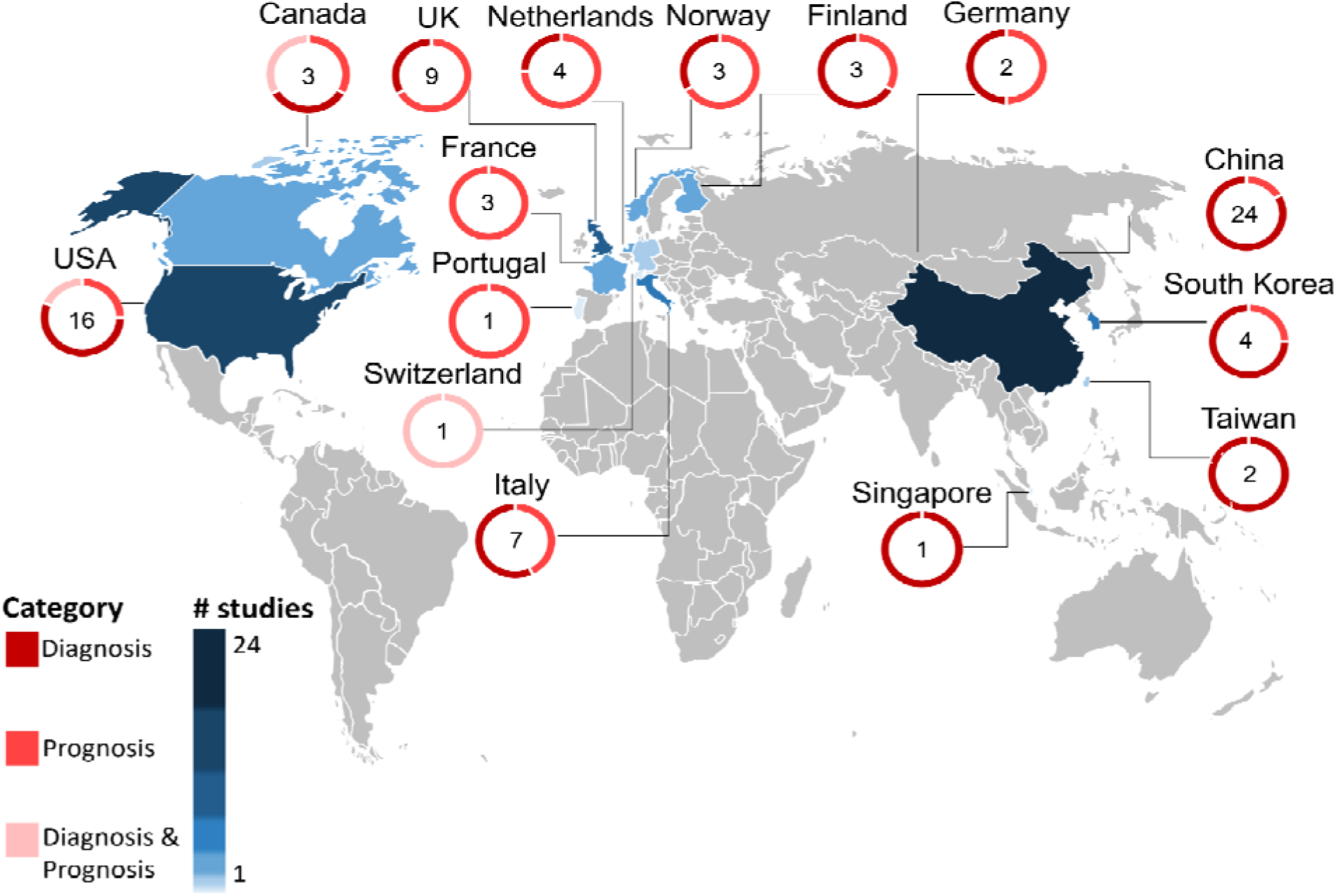
Countries of institutional affiliation for the first and last authors of each included publication. Note that the first and last authors had different affiliations in eight papers. The numbers on the map thus sum to 83 rather than 75 (total number of included studies).

#### 3.2.2 Study focus

Among the 75 included studies, 43 (57%) focused on diagnosis, 27 (36%) on prognosis, and five (7%) on both (**Supplementary Table 1**). More than two-thirds (70%) of the studies used private or local datasets (n=35 diagnosis, n=16 prognosis, n=2 both). When publicly available datasets were used (n=12 for diagnosis, n=14 for prognosis, n=4 for both), ADNI was the most frequent choice (n=17/30). Only 14 studies included two or more cohorts in their analyses (n=7 for diagnosis, n=5 for prognosis, n=2 for both). ADNI was the most commonly used dataset in these cases (n=9/14).

#### 3.2.3 Participant demographics

The mean age of participants in the studies was 71.7 years (SD 8.5), with a mean age of 69.9 years (SD 9.31) in diagnostic studies and 72.1 years (SD 8.3) in prognostic studies. There was a relatively balanced representation of men and women across the studies, with women making up 54% of participants overall (48.1% in diagnostic studies and 55% in prognostic studies).

Only six studies (8%) provided information about the ethnicity of the study population (n=3 diagnosis, n=3 prognosis). In these studies, 6217 (79%) participants were reported as White (n=1 diagnosis, n=3 prognosis), 209 (3%) as Asian (n=2 diagnosis), 934 (12%) as Black (n=1 prognosis), and 476 (6%) as “other” (n=2 diagnosis, n=2 prognosis).

### 3.3 ML methods

#### 3.3.1 Application of ML methods

A total of 23 different ML methods were employed for the diagnosis or prognosis of cognitive impairment and dementia based on vascular neuroimaging features (**Figure 3**). These methods spanned eight categories (**Supplementary BOX 2**). For diagnosis (**Figure 3A**), the most popular ML categories were instance-based, regression, and ensemble algorithms, with SVM (instance-based), logistic regression (regression), and random forest (ensemble) being the most commonly used models. For prognosis (**Figure 3B**), the top 3 categories remained the same, with Cox regression (regression), SVM (instance-based), and random forest (ensemble) being the most frequently used models.

**Figure 3.**
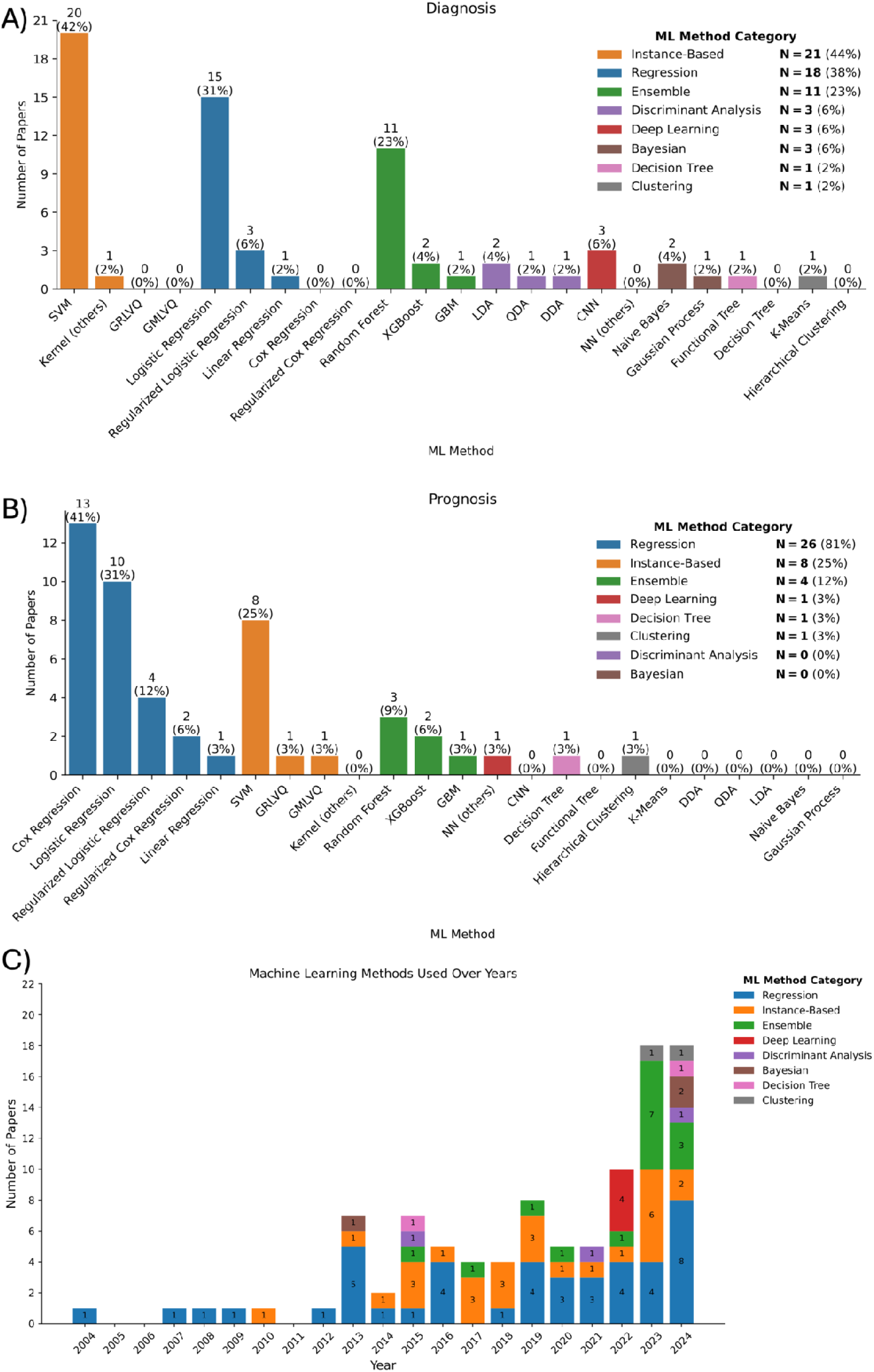
Number of papers using each machine learning (ML) method in our review, faceted by A) diagnosis or B) prognosis, and C) use of different ML categories over time. Different colours correspond to different ML categories. Note that as one paper may use multiple ML methods/categories, the percentages in A) and B) may not sum up to 100%. Kernel (others) = Kernel methods other than SVM; NN (others) = Neural networks other than CNN.

Overall, SVM was the most popular ML method, appearing in 26 (35%) of the 75 papers reviewed. This was followed closely by standard logistic regression, which was used in 24 (32%) papers. These two methods were applied almost twice as often as the next most common methods, namely random forest and Cox regression, which were featured in 14 (19%) and 13 (17%) papers, respectively. Other methods were much less common. Strikingly, despite the growing prominence of deep learning over the past decade, its application in diagnosing and predicting cognitive impairment and dementia based on vascular neuroimaging features remains limited. Only four (5%) of the papers employed deep learning, with three of them using CNNs. Very few studies used Bayesian or discriminant analysis methods, and we observed their application solely in diagnosis.

#### 3.3.2 Popularity over time

The application of ML techniques for diagnosis and prediction of cognitive impairment and dementia based on vascular neuroimaging features has experienced a significant growth over the past decade (**Figure 3C**). Cox regression was the first technique we identified that used neurovascular features, namely stroke lesions and WMH, to predict dementia [99]. Logistic regression and SVM have remained popular and widely used since their first application in 2007 and 2010, respectively. The application of random forests, on the other hand, was first identified in 2015 and has only recently reached its highest level of usage, as it was used in around one-third of the works in 2023 (36%). Neural networks and XGBoost appeared only in 2022.

#### 3.3.3 Performance assessment and generalisability

Adequately measuring the performance and generalisability of ML prediction algorithms is crucial for the translation of these models into real-world clinical settings. Out of a total of 75 papers (n=48 diagnosis; n=32 prognosis), eight combined datasets from different sources or assessed the out-of-sample performance of their models on external datasets (n=7 diagnosis; n=4 prognosis). The rest either divided a single dataset from the start into training and testing sets (n=3 diagnosis; n=2 prognosis), employed cross-validation (n=28 diagnosis; n=17 prognosis), or did not specify whether they derived performance metrics from a sample external to the training sample (n=10 diagnosis; n=9 prognosis).

### 3.4 Neuroimaging modalities and features

The papers included in this review assessed the diagnostic and prognostic capabilities of vascular neuroimaging features derived from structural, diffusion, and functional MRI (fMRI), as well as computed tomography and positron emission tomography (**Figure 4**). Structural MRI was by far the most widely used neuroimaging modality, appearing in 67 (89%) studies, whereas computed tomography was the least used, appearing in only one (1%) paper. A total of 27 (36%) studies leveraged two or more imaging modalities. When this occurred, structural MRI and diffusion-based MRI were the most common combination, used in 19 out of 27 (70%) papers.

**Figure 4.**
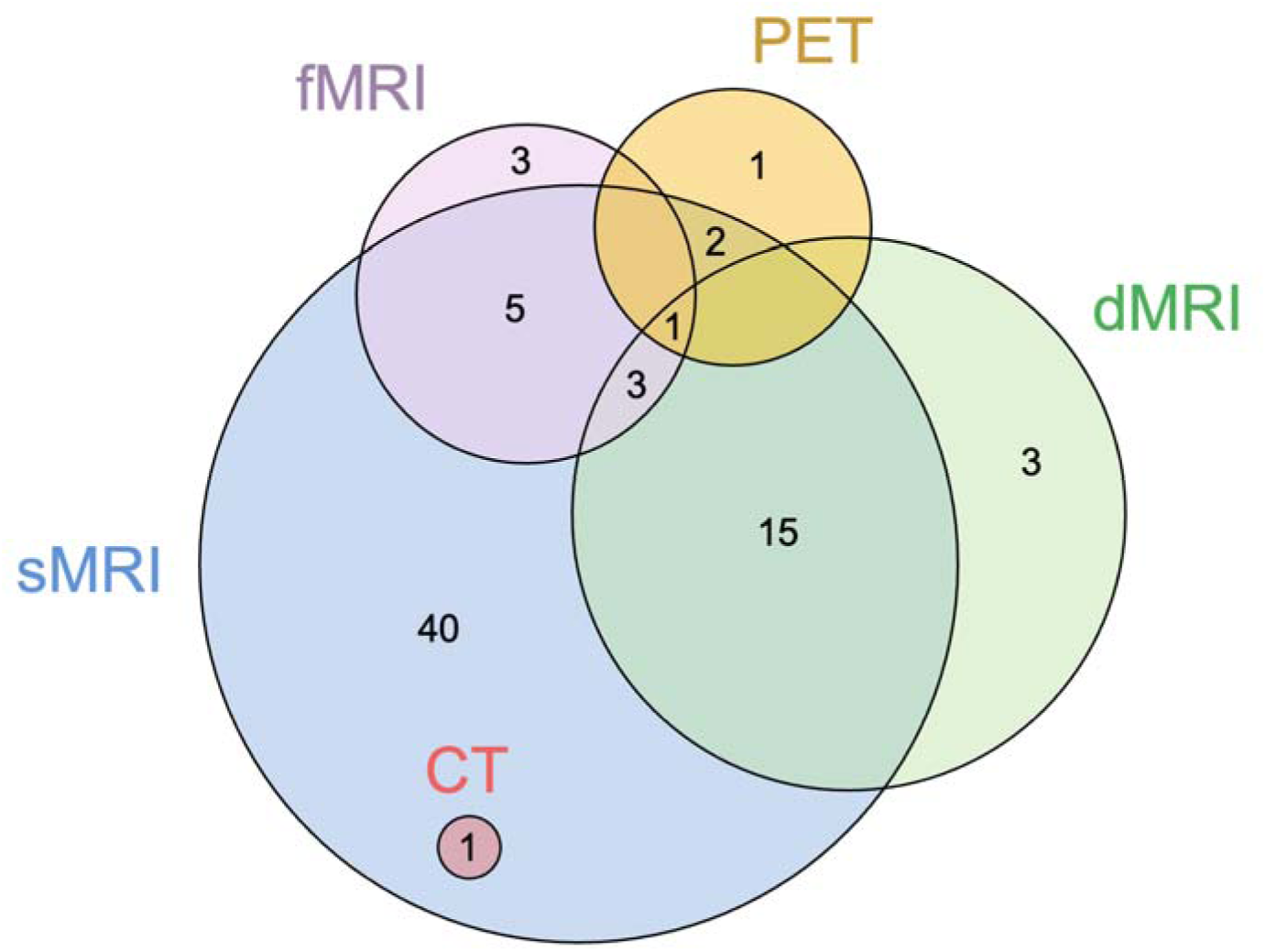
Neuroimaging techniques used to extract vascular neuroimaging features in the studies included in the review. Abbreviations: CT=computed tomography; dMRI=diffusion-based MRI; fMRI=functional MRI, sMRI=structural MRI; PET=positron emission tomography.

#### 3.4.1 Diagnosis

Around 40% of the diagnostic studies (n=20) leveraged vascular neuroimaging features obtained from structural MRI sequences. Every study that used structural MRI quantified WMH. Other neuroimaging features were less frequently considered: perivascular spaces (n=2), cerebral microbleeds (n=2), lacunes (n=3), and stroke lesion volume or type (n=2).

Non-lesion measurements were also common in many diagnostic studies. These included assessments of microstructural integrity from regional white matter (n=12), fMRI or diffusion MRI-based connectivity (n=8), or other, less conventional imaging features (n=7), such as tissue textures (n=2), white matter density (n=1), fMRI-derived amplitude of low-frequency fluctuation (n=2), FDG positron emission tomography derived “metabolic cognitive signature” (n=1), and iron deposition (n=1).

Of the 48 studies reporting diagnostic analyses, most used quantitative assessments of vascular neuroimaging features (n=40). Nine (19%) employed clinical visual ratings, of which two relied solely on visual ratings.

#### 3.4.2 Prognosis

Twenty-four (75%) prognostic studies used vascular neuroimaging features derived from structural MRI. WMH were assessed in every study. Evaluating the prognostic value of stroke lesions was also common in prognostic studies (n=6). Other neuroimaging features, including lacunes (n=1), perivascular spaces (n=2), cerebral microbleeds (n=1), and stroke lesion volume (n=2), were less relatively common.

Non-lesion measurements were less common in prognostic than in diagnostic studies. Only five studies used diffusion tensor imaging sampled and no studies investigated other diffusion-based signal modelling techniques. Two studies investigated connectivity measures: one using diffusion-based structural connectivity, and one using WMH-based disconnectome measures. Other vascular markers used in prognosis studies included tissue texture analysis (n=2), stroke aetiology and antecedents (n=2), and susceptibility-weighted imaging-based markers such as iron deposition (n=1).

Visual ratings were used in half of studies reporting prognostic results (16 out of 32). Most ratings were conducted on WMH burden, with one study reporting solely perivascular space ratings. Nine studies included additional quantitative measures alongside visual ratings, while the remaining relied solely on visual ratings (n=7). There was significant variation in the rating scales used across studies. For example, the six prognostic publications that used visual ratings of WMH employed a range of methods, including binary measures with different cut-offs as well as grading scales with 3, 4, 10, and 11 points.

#### 3.4.3 Temporal changes in vascular neuroimaging features

Studies using diffusion-based indices (e.g., fractional anisotropy and mean diffusivity) have decreased in recent years. Before 2020, diffusion imaging studies made up 41% of studies (n=16), compared to just 17% of studies published between 2020-2024 (n=6). Recent years have witnessed the adoption of novel techniques including graph theory, pattern analyses, and other methods of connectivity assessment, amplitude of low-frequency fluctuation, and composite brain signatures, making up a fifth (19%; n=7) of studies published in 2020-2024. Studies relying solely on clinical visual ratings have not changed in recent years (n=5 before 2020, n=4 since 2020).

#### 3.4.4 MRI scanner strength

MRI scanner strength details were reported in 65 (84%) studies (n=40 diagnostic, n=26 prognostic, n=5 both). Diagnostic studies were primarily carried out on 3T scanners. Most studies used a consistent scanner strength across participants and scans (n=33), with 3T being the most common (n=27), followed by 1.5T (n=5), and one study using a 4T scanner. A few studies (n=7) used scanners of different field strengths; six of these combined 1.5T and 3T scanners, while one study used both 1T and 1.5T scanners.

Most prognostic studies, on the other hand, were conducted on 1.5T scanners. Among the 27 prognostic studies that reported scanner strength, the majority used only 1.5T scanners (n=15), with about half as many relying exclusively on 3T scanners (n=7). A few studies (n=5) employed a mix of scanner strengths; two combined 0.5T and 1.5T scanners, and three studies used both 1.5T and 3T scanners.

The use of higher field strength scanners has increased over time. Before 2020, more than half of the studies (59%) relied predominantly on 1.5T scanners (n=23). Since 2020, reliance on 1.5T scanners has declined, with only 14% of studies using 1.5T scanners (n=4), while 62% (n=18) used 3T scanners exclusively.

### 3.5 Dementia

We identified 47 studies that focused on dementia, with 26 (55%) targeting diagnosis, 17 (36%) prognosis, and four (9%) both. Remarkably, despite the review’s strong emphasis on vascular aspects, diagnostic studies predominantly concentrated on AD (n=23). Other forms of dementia were also examined but with less frequency: vascular dementia (n=4), frontotemporal dementia (n=3), Lewy body dementia (n=3), behavioural variant frontotemporal dementia (n=1), Parkinson’s disease (n=1), post-stroke dementia (n=1), progressive non-fluent aphasia (n=1), and semantic dementia (n=1). Prognostic studies also focused largely on AD (n=10), with vascular dementia following (n=6). Mixed dementias were each examined by four prognostic studies, and a single prognostic study also explored frontotemporal dementia.

#### 3.5.1 Assessment criteria

Dementia diagnosis was primarily relied on published clinical criteria or via consensus diagnosis by experienced neurologists based on clinical evaluations, including cognitive tests, general neurological exams, and collateral information. Studies of AD employed no less than six different diagnostic criteria, namely Diagnostic and Statistical Manual of Mental Disorders (DSM versions III-R, IV or V) [100], National Institute of Neurological and Communicative Disorders and Stroke and the Alzheimer’s Disease and Related Disorders Association (NINCDS or NINCDS-ADRDA) [101], National Institute on Aging and Alzheimer’s Association (NIA-AA) [102, 103], National Institute on Aging– Alzheimer’s Disease Centers (NIA-ADRC), National Institute of Neurological Disorders and Stroke and Association Internationale pour la Recherche et l’Enseignement en Neurosciences (NINDS-AIREN) [104], and Alzheimer’s Disease Diagnostic and Treatment Centers (ADDTC) [105].

In addition to the diagnostic variability, six studies did not specify the criteria for diagnosing dementia (n=4 diagnosis, n=2 prognosis) and six diagnostic studies determined dementia diagnoses without adhering to standard criteria. Instead, they relied on specific thresholds from cognitive tests, including the Mini-Mental State Examination (MMSE) (n=1), the Clinical Dementia Rating scale (CDR) (n=1), the Montreal Cognitive Assessment (MoCA) scores (n=1), and various combinations of these test (n=3).

### 3.6 Cognitive impairment

We identified 45 articles addressing cognitive impairment (without a specific dementia diagnosis), with 26 (58%) focusing on diagnosis, 14 (31%) on prognosis, and five (11%) assessing both diagnosis and prognosis. The definition, subtype, and potential aetiology of cognitive impairment varied substantially across the studies.

Most diagnostic studies (52%) that studied cognitive impairment examined cognitive impairment linked to CSVD (n=11), such as strokes (n=1), WMH (n=4), subcortical ischemic vascular disease (n=4), and vascular cognitive impairment (n=3). However, a notable proportion of them (62%) also targeted mild cognitive impairment (MCI) (n=12) or its amnestic and non-amnestic subtypes (n=4). Cognitive impairment associated with Parkinson’s disease (n=1) and coronary artery disease (n=1) was also examined.

Prognostic studies mainly investigated either the progression of MCI to dementia (n=15) or the prognosis of cognitive impairment related to CSVD (n=3). Specific subtypes of MCI, such as amnestic MCI (n=1), as well as MCI subgroups, such as stable MCI and progressive MCI (n=7), were also investigated. For studies on CSVD-related cognitive impairment, the focus was mainly on strokes (n=3) or general accumulation of vascular lesions (n=1).

#### 3.6.1 Assessment criteria

While there was some consistency across studies, the diagnostic criteria consisted of a variety of definitions of cognitive impairment. Most papers that focused on cognitive impairment relied on neuropsychological tests (n=18 diagnosis, n=11 prognosis. For example, performance below 1.5 standard deviations from the mean on cognitive tests was deemed indicative of MCI. A significant variability in the choice of cognitive tests was noted. Some papers adopted established clinical criteria, such as those proposed by Petersen et al. [106, 107] or the DSM-5 [100], or utilised consensus diagnoses from multiple neurologists (n=1 diagnosis, n=4 prognosis). In the other instances, a combination of neuropsychological tests and clinical criteria was used (n=5 diagnosis, n=2 prognosis) with some studies also incorporating vascular neuroimaging measures, such as including WMH burden to support MCI diagnosis (n=5 diagnosis, n=1 prognosis). Three papers did not specify the criteria used to identify MCI.

### 3.7 Results meta-analysis

#### 3.7.1 Healthy controls versus Alzheimer’s dementia

Seven studies reported AUC measures for classifying AD-dementia versus healthy controls (see **Figure 5A**). Of these, five did not report any measures of variability; the standard error for these AUC values was therefore estimated. The pooled AUC was 0.88 [95% confidence interval (CI) 0.85 – 0.92] (**Figure 5A**), with significant heterogeneity across studies.

**Figure 5.**
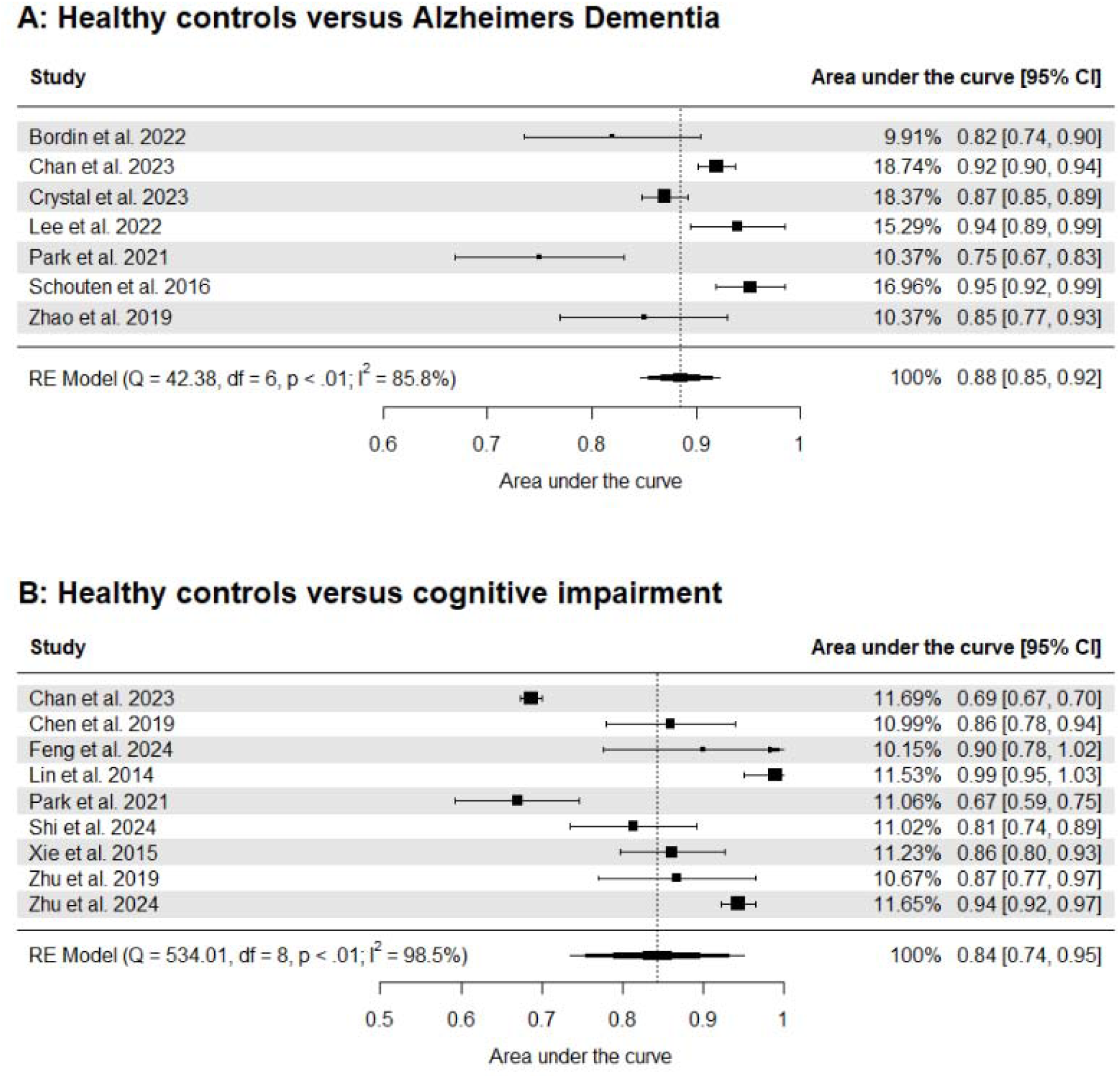
Meta-analysis of studies classifying Alzheimer’s Dementia or cognitive impairment vs healthy controls. Confidence intervals might exceed 1.00 because standard errors have been estimated due to missing data.

#### 3.7.2 Healthy controls versus cognitive impairment

Nine studies used ML algorithms to classify cognitive impairment versus healthy controls (see **Figure 5B**). Only two of these studies reported measures of variability, therefore all other standard errors for the AUC were estimated. The pooled AUC was 0.84 [95% CI 0.74 – 0.95], with significant heterogeneity among studies (**Figure 5B**) [32, 33, 51, 58, 79, 85, 91, 96, 98].

#### 3.7.3 Healthy controls versus all-cause dementia

Nine studies utilised ML methods to diagnose all-cause dementia versus healthy controls, of which two studies (Chan et al. 2023 [32] and Lee et al. 2022 [47]) made multiple comparisons. Here we selected the comparisons with the largest sample sizes included. Of these studies, three reported measures of variability for the AUC, the standard error for all remaining AUC values were therefore estimated. The pooled AUC was 0.88 [95% CI 0.83 – 0.93] (**Supplementary Figure 1A**), with significant heterogeneity across studies. Including the other two comparisons of Chan et al. 2023 [32] and Lee et al. 2022 [47] in the meta-analysis yielded similar results (**Supplementary Figure 1B**).

### 3.8 Risk of bias assessment

#### 3.8.1 Diagnosis

For diagnostic studies, we conducted risk of bias assessment using the QUADAS-2 framework to establish potential biases regarding patient selection, index method bias, reference accuracy, blinding, consistency of references, participant inclusion, and method applicability (**Supplementary Figure 2A and Table 2A**). Approximately 40% of the papers exhibited a low risk of bias across the aforementioned domains (flow and timing: n=21; reference standard: n=28; index test: n=31; patient selection=32). The patient selection domain had by far the highest risks of bias (n=19), with studies either using case-control designs or failing to adequately disclose patient selection details. Concerns about method applicability in diagnostic studies were minimal, with 83% of studies having low risk of bias.

#### 3.8.2 Prognosis

For prognostic studies, we conducted risk of bias assessment using the PROBAST framework to establish potential biases in prediction models regarding patient selection, predictor measurement, outcome measurement, and analysis and model evaluation (**Supplementary Figure 2B and Table 2B**). We rated 28% of the papers as showing a low risk of bias, 19% as unclear, and the remaining 53% as showing some risk of bias, primarily on the analysis domain. The primary reasons were unclear exclusion criteria from prognostic analyses and lack of clarity regarding how model overfitting and optimism in model performance were addressed. Concerns about applicability were minimal, with 94% of the studies showing low concerns.

## IV. DISCUSSION

This systematic review and meta-analysis summarise 75 studies examining the role of CSVD neuroimaging markers in ML-based diagnosis and prognosis of cognitive impairment and dementia. The field has grown substantially over the past two decades, with nearly 60% of studies being published in the last two years. Key findings reveal a mix of insights. Positively, most studies used local datasets with balanced sex representation, and ML models leveraging CSVD features achieved high diagnostic performance. However, many relied on single datasets, case-control designs, lacked external validation, or showed limited transparency regarding overfitting. Unexpectedly, most studies focused on AD rather than vascular dementia. Additionally, while XGBoost and neural networks are gaining traction, traditional methods like Cox regression, logistic regression, and SVM remain dominant. The following sections expand on these findings and place them within a broader context.

### 4.1 Vascular neuroimaging and its contributions to neurodegeneration

Most studies on dementia diagnosis and prognosis focused on AD (23/30 diagnostic; 10/21 prognostic) rather than vascular dementia (4/30 diagnostic; 6/21 prognostic). For example, seven of the nine studies that compared all-cause dementia with healthy controls in the meta-analysis focused on AD-related dementia. These seven studies that incorporated vascular features in ML models demonstrated strong performance (AUC 0.88 [95%-CI 0.85, 0.92]). This indicates not only the benefit of including CSVD in AD diagnostic classifiers, but also reflects a growing interest in the role of CSVD and vascular neuroimaging in dementia, particularly in AD-related dementia.

The focus on AD surprised us, given the review’s emphasis on vascular neuroimaging and CSVD, though aspects discussed in the literature may help explain this trend. AD is the leading cause of dementia, and CSVD is one of the most common conditions in clinics [4]. It is therefore expected that CSVD co-occurs and interacts with AD as well as other neurodegenerative disorders [108–113]. The role of vascular risk factors in the pathogenesis and progression of AD is well established [114], extending beyond their mere coexistence [115]. Potential mechanisms include impairment of the neurovascular unit [116, 117], disruption of the glymphatic system [118], and hypoperfusion and hypoxia [116, 119]. This coexistence is now formally recognised in the most recently proposed AD diagnostic and staging criteria, where vascular biomarkers are identified as indicators of non-AD co-pathology that may accelerate symptom progression [7].

Considering vascular risk factors in the progression of neurological conditions such as vascular dementia and AD creates opportunities for prevention [116, 120]. Modifiable CSVD risk factors, including diabetes, hypertension, smoking, obesity, and high low-density lipoprotein cholesterol, can be addressed with cost-effective treatments, such as hypertensive medications and lifestyle changes [121]. These measures may reduce vascular risks and delay or prevent cognitive decline, potentially alleviating the societal burden of dementia. Recent studies in birth cohorts show a decline in cerebrovascular pathology [122], which may explain the decrease in age-adjusted dementia incidence [123–125]. Understanding and identifying CSVD features associated with AD using ML approaches may therefore support early diagnosis and risk prediction, permitting early effective preventive strategies.

### 4.2 Future of ML and vascular neuroimaging

#### 4.2.1 Neuroimaging

The majority of studies in our review relied on structural MRI data, with only a single study using computed tomography. A significant shift in this trend is unlikely in the coming years, though it is probable that new and advanced imaging methods will begin to appear more frequently in the literature. While assessing the human cerebral microvasculature *in vivo* using conventional imaging technologies remains challenging, advancements in MRI with higher field strengths (7T and above) as well as other imaging technologies help us get closer to detecting subtle vascular changes and visualising small-calibre blood vessels [126, 127]. A promising candidate is ultrasound localization microscopy—a super-resolution ultrasound technique—which represents the first *in vivo* imaging technique capable of achieving capillary-scale (micron-level) resolution with an imaging penetration depth of several centimetres [128–130].

The recent incorporating of non-lesional outcomes into the assessment of CSVD in the STRIVE II criteria will likely enrich our understanding of CSVD and its contributions to neurodegenerative diseases. Investigating functional markers, such as those derived from fMRI [131], could enable determining the extent to which CSVD leads to impaired neurovascular coupling, neural network communication, and functional connectivity, and ultimate contributes to cognitive decline [132]. Multimodal imaging with optically pumped magnetometers, electroencephalography, magnetoelectroencephalography, and functional near infrared spectroscopy—which can simultaneously record electrophysiological and haemodynamic changes at fine-resolution—may enable the detection of the earliest vascular changes that occur in dementia [133]. Despite the value of these imaging techniques, it will likely take some time to build the necessary size of datasets for applying ML algorithms.

#### 4.2.2 Harmonisation

A key barrier to building large multimodal datasets is the harmonisation of data and sharing of protocols and data between centres. This may not be as important for novel methods, but improved data harmonisation techniques for existing modalities are desirable to support large-scale, multi-centre studies of early dementia identification. The HARNESS initiative, for instance, aims to develop tools for harmonising vascular neuroimaging data [134]. While this initiative primarily focuses on CSVD, the techniques developed are also applicable for harmonising data for other neurodegenerative dementias. Other examples of successful harmonisation to enhance consistency across WMH measures in healthy elderly cohorts (Whitehall II, 713 participants, and UK Biobank, 2295 participants) [135] and memory clinic cohorts [136] have already been published in the literature. A continued focus on the harmonisation of vascular neuroimaging data will allow much larger predictive studies to be conducted than has been possible hitherto.

#### 4.2.1 Treatments

Identifying and quantifying CSVD is relevant for the selection and stratification of dementia treatments, as illustrated in anti-amyloid therapies like Aducanumab or Lecanemab. Accurate identification of patients who are most likely to benefit from treatment, while minimising risks, is crucial. CSVD markers such as WMH [137–139], microbleeds [137–141], and perivascular spaces [142–144] play a pivotal role in the diagnosis of cerebral amyloid angiopathy, a major risk factor for adverse treatment outcomes [145–148]. AI models, with their ability to analyse large datasets and detect subtle neuroimaging features, offer a promising avenue for improving treatment planning [149]. These models may help predict which patients are at increased risk for conditions like amyloid-related imaging abnormalities [150], particularly in higher-risk patients such as those with WMH or cerebral amyloid angiopathy who require particularly careful consideration before being prescribed treatments like Aducanumab [151]. In addition to predicting complications, enhanced prognostic models incorporating CSVD changes in dementia may also help to identify those most likely to benefit from treatment. AI-based predictions could provide clinicians with valuable tools to tailor therapeutics, thus enhancing safety and efficacy while facilitating precision medicine approaches for patients [152–154].

### 4.3 Limitations of the field and recommendations

We have identified several limitations of the field and present corresponding recommendations, as summarised in **BOX 1**.

#### BOX 1 — Recommendations to move toward clinically useful, machine learning methods applied to vascular neuroimaging for cognitive impairment and dementia

##### Reporting

– Inclusion of measures of variance and/or confidence intervals for all performance metrics
– Full disclosure of all results, including those that do not reach statistical significance

##### Generalisability

– Use of external independent datasets for validation

##### From curated to real-world datasets

– Inclusion of diverse populations
– Investigation of a broad spectrum of neurodegenerative diseases, including a greater focus on vascular dementia

##### Fairness and representativeness

– Exploration of sex-specific and ethnicity-specific contributions of vascular features for neurodegeneration

#### 4.3.1 Reporting

Many studies did not report variances or confidence intervals for model performance, making it challenging to combine their findings in a meta-analysis. For future studies, *we strongly recommend the inclusion of confidence intervals for all performance metrics*, *following established reporting standards such as the TRIPOD+AI guideline* [155]. Careful consideration should also be given to the choice of metrics. For instance, in cases of pronounced class imbalance, reporting sensitivity and specificity alongside AUC can be crucial. Additionally, *we advocate for the full disclosure of all results, including those that do not achieve statistical significance, to enhance the completeness and unbiased representation of study findings*.

#### 4.3.2 Generalisability

Despite achieving high accuracy metrics, the translation of predictive ML algorithms to real-world clinical settings requires extensive external validation. Out of 75 studies included in this review, solely eleven trained and/or tested their models using datasets from different sources. A recent study by Chekround and colleagues found that, even with adequately cross-validated algorithms, the performance of these ML models was consistently lower in external datasets [156]. *We thus recommend the validation on external datasets to properly assess the true performance of these algorithms*. This is especially crucial for more complex non-linear models which may fit the specific idiosyncrasies of the data that they were trained on, in turn reduce their generalisability.

#### 4.3.3 From curated to real-world datasets

Neuropathological studies often show a complex constellation of brain pathologies across neurodegenerative dementias [157], which may exist to varying degrees alongside cerebrovascular disease [158]. These mixed or multiple pathologies can make it challenging for ML algorithms to accurately classify cases and predict disease progression, in turn lowering the robustness and generalisability of ML models. Hence, training ML algorithms that assess cerebrovascular disease in the presence of other neuropathological indicators, using features derived from well-characterised, multimodal datasets across neurodegenerative dementias, is highly desirable. *This underscores the importance of leveraging more representative, deeply phenotyped, real-world datasets that include diverse types of dementias*. As we move closer towards individualised, precision medicine approaches, such a strategy will optimise the translation of ML algorithms in clinical settings while ensuring their applicability across ethnically and socioeconomically diverse populations worldwide.

#### 4.3.4 Fairness and representativeness

Many included studies used case-control designs, which, while useful for comparisons, often introduce selection bias, especially when controls are not well-matched to cases [19]. Additionally, several studies lacked clear inclusion/exclusion criteria, undermining replicability and generalisability. Six studies also failed to clearly specify diagnostic criteria, further impacting repeatability and reliability. Although there was a balanced representation of men and women in the included studies, only one study assessed sex differences in the classification of dementia [72]. It is well-established that women are at higher risk of developing dementia and WMH [159, 160]. Similarly, while ethnicity can modulate developing dementia [161], only six studies reported the ethnicity of their participants. The inconsistent reporting of demographic factors, such as ethnicity, raises concerns about diversity in dementia research, given that most dementia cohorts tend to be well educated, and of higher socioeconomic status than what would be expected based on census data. This has important implications for accurately predicting dementia risk especially since ethnicity, socioeconomic background, and education are important factors that can modulate an individual’s cumulative risk for developing dementia [162–164]. Non-representative samples may limit the applicability of the findings to broader, more diverse populations. *We recommend that future cohort studies seek to collect and consistently report data from diverse populations, and that studies explore sex– and ethnicity-specific classifications of dementia*.

### 4.4 Strengths and limitations

This work has major strengths. We performed a systematic search across nine databases to provide a comprehensive summary of the existing evidence on the application of vascular neuroimaging in ML-based diagnosis and prognosis of cognitive impairment and dementia. Our approach followed a carefully structured methodology to ensure both transparency and reproducibility. This included pre-registering our protocol and adhering to PRISMA guidelines. All papers included in the title and abstract screening, full-text screening, data extraction, and risk of bias assessment phases— during both the initial review and the re-run—were independently reviewed by multiple reviewers, with each paper evaluated by two reviewers and a third consulted to resolve any outstanding conflicts. We systematically assessed the risk of bias and the overall quality of the studies using established quality assessment tools, specifically QUADAS-2 and PROBAST, to ensure a robust evaluation of the evidence included in our review. Through these rigorous standards, we aimed to synthesise high-quality evidence that can guide future research to accelerate the integration of ML into clinical practice.

This work has three main limitations. First, a limitation of our meta-analysis is the significant variation in covariates and sample sizes across studies, which may have led to heterogeneity in the predictive models and potentially affected AUC comparability. Second, we did not assess the added value of neuroimaging markers of CSVD in diagnosing or predicting dementia, as only a few studies compared the performance of ML models with and without CSVD markers. Third, the risk of bias assessment tools used here were not designed for AI studies, and while recent efforts have been made, such tools are still in their infancy [155, 165] — the rationale for using accepted risk of bias tools for the tasks, QUADAS-2 and PROBAST.

## V. CONCLUSION

CSVD markers are playing an increasing role in ML-based diagnosis and prognosis of dementia and cognitive impairment, with models leveraging these markers already demonstrating strong performance in distinguishing individuals with and without cognitive impairment and dementia. However, challenges remain of reporting standards, generalisability, and fairness that hinder the widespread adoption of these approaches. As the cerebrovascular interest subgroup of the international DEMON Network Imaging Working Group, we have collaboratively developed a set of targeted recommendations to drive progress in this field over the next decade. We firmly believe that adopting these recommendations will greatly accelerate the integration of ML methods into clinical practice, delivering meaningful benefits for patients.

## AUTHOR CONTRIBUTIONS

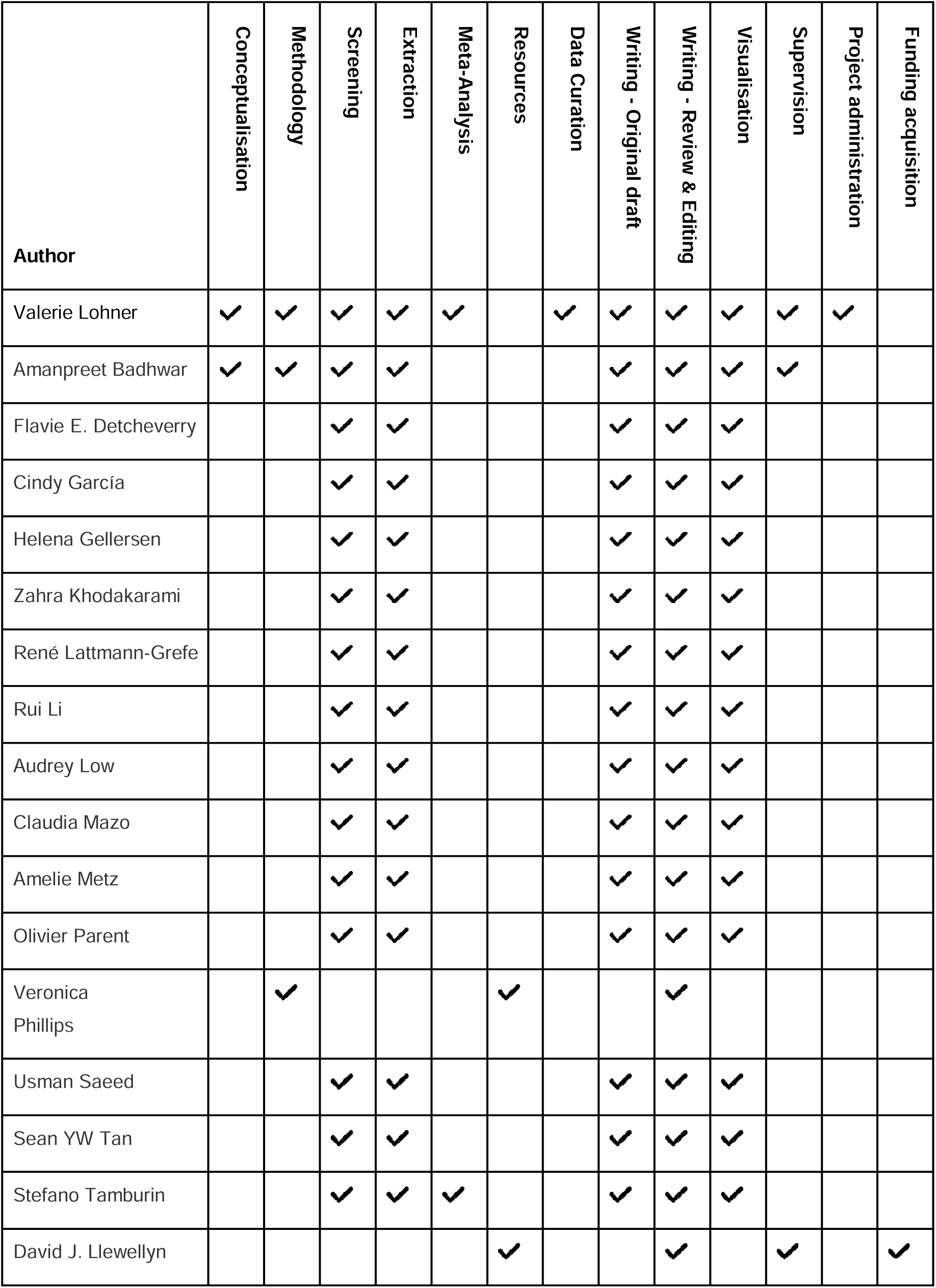
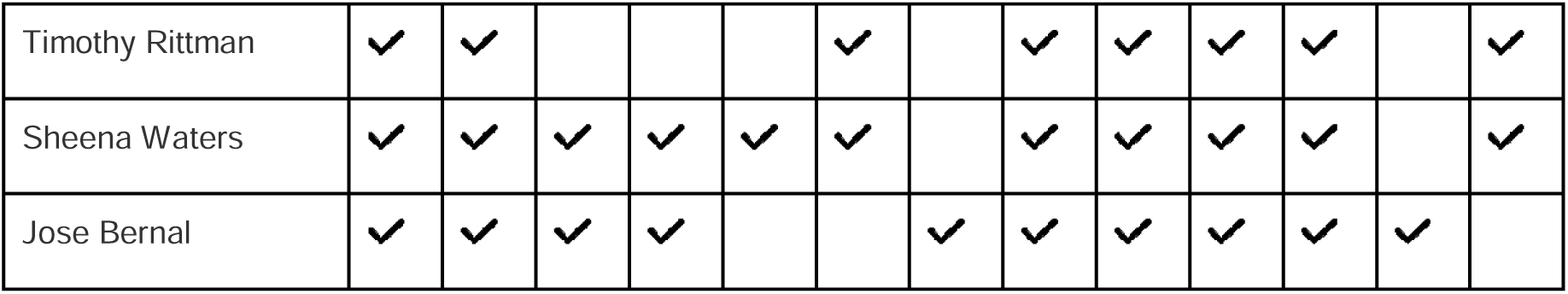

## Data Availability

All data produced in the present work are contained in the manuscript and in the supplementary material.

## ACKNOWLEDGEMENT

Funding: VL is supported by the Marga and Walter Boll Foundation, Kerpen, Germany. AB is supported by Fonds de Recherche Québec – Santé (FRQS) Chercheurs boursiers Junior 1 (2020–2024) and the Fonds de soutien à la recherche pour les neurosciences du vieillissement from the Fondation Courtois. FED is supported by Fonds de recherche du Québec – Santé (FRQS). CG acknowledges support from Fonds de recherche du Québec – Santé (FRQS), Healthy Brains Healthy Lives (HBHL) and McGill University. RL is supported by a PhD funding from Trinity College, Cambridge, UK. HMG is supported by the BrightFocus Foundation, the German Center for Neurodegenerative Diseases (DZNE) Foundation, the Joachim Herz Foundation, and St John’s College, University of Cambridge, UK. ZK is supported by National Institutes on Aging grant RF1 AG056014. AL is supported by a research fellowship from Race Against Dementia. AM is supported by Fonds de recherche du Québec – Santé (FRQ-S) and Healthy Brains Healthy Lives (HBHL). OP is supported by the Alzheimer Society of Canada. US acknowledges support from the Ontario Graduate Scholarship, University of Toronto, Canada. TR is supported by the NIHR Cambridge Biomedical Research Centre (BRC-1215-20014) and Alzheimer’s Research UK (ARUK-SRF2023B-005). DJL and the DEMON Network are supported by Alzheimer’s Research UK. DJL is also supported by National Institute for Health and Care Research (NIHR) Applied Research Collaboration South West Peninsula and the NIHR Exeter Biomedical Research Centre. SW is supported by the Global Parkinson’s Genetics Program (GP2) since April 2024. GP2 is funded by the Aligning Science Across Parkinson’s (ASAP) initiative and implemented by The Michael J. Fox Foundation for Parkinson’s Research (https://gp2.org). Prior to April 2024, SW was supported by UKRI Innovate UK. The views and opinions expressed are those of the authors and not necessarily those of the PIA membership, NIHR, ISTAART, the Alzheimer’s Association, or the Department of Health and Social Care.

## REFERENCES

[1] Duering M, Biessels GJ, Brodtmann A, Chen C, Cordonnier C, de Leeuw F-E, et al. Neuroimaging standards for research into small vessel disease-advances since 2013. The Lancet Neurology. 2023;22:602–18.

[2] Wardlaw JM, Smith C, Dichgans M. Small vessel disease: mechanisms and clinical implications. The Lancet Neurology. 2019;18:684–96.

[3] Ter Telgte A, Duering M. Cerebral Small Vessel Disease: Advancing Knowledge With Neuroimaging. Stroke. 2024;55:1686–8.

[4] Cannistraro RJ, Badi M, Eidelman BH, Dickson DW, Middlebrooks EH, Meschia JF. CNS small vessel disease: A clinical review. Neurology. 2019;92:1146–56.

[5] Clancy U, Appleton JP, Arteaga C, Doubal FN, Bath PM, Wardlaw JM. Clinical management of cerebral small vessel disease: a call for a holistic approach. Chin Med J (Engl). 2020;134:127–42.

[6] Yang HD, Kim DH, Lee SB, Young LD. History of Alzheimer’s Disease. Dement Neurocogn Disord. 2016;15:115–21.

[7] Jack Jr. CR, Andrews JS, Beach TG, Buracchio T, Dunn B, Graf A, et al. Revised criteria for diagnosis and staging of Alzheimer’s disease: Alzheimer’s Association Workgroup. Alzheimer’s & Dementia. 2024;20:5143–69.

[8] Hampel H, Elhage A, Cho M, Apostolova LG, Nicoll JAR, Atri A. Amyloid-related imaging abnormalities (ARIA): radiological, biological and clinical characteristics. Brain. 2023;146:4414–24.

[9] ter Telgte A, van Leijsen EMC, Wiegertjes K, Klijn CJM, Tuladhar AM, de Leeuw F-E. Cerebral small vessel disease: from a focal to a global perspective. Nat Rev Neurol. 2018;14:387–98.

[10] Gurol ME, Sacco RL, McCullough LD. Multiple Faces of Cerebral Small Vessel Diseases. Stroke. 2020;51:9–11.

[11] Jiang J, Wang D, Song Y, Sachdev PS, Wen W. Computer-aided extraction of select MRI markers of cerebral small vessel disease: A systematic review. NeuroImage. 2022;261:119528.

[12] Waymont JMJ, Valdés Hernández MdC, Bernal J, Duarte Coello R, Brown R, Chappell FM, et al. Systematic review and meta-analysis of automated methods for quantifying enlarged perivascular spaces in the brain. NeuroImage. 2024;297:120685.

[13] Hu X, Liu L, Xiong M, Lu J. Application of artificial intelligence-based magnetic resonance imaging in diagnosis of cerebral small vessel disease. CNS Neuroscience & Therapeutics. 2024;30:e14841.

[14] Borchert RJ, Azevedo T, Badhwar A, Bernal J, Betts M, Bruffaerts R, et al. Artificial intelligence for diagnostic and prognostic neuroimaging in dementia: A systematic review. Alzheimers Dement. 2023;19:5885–904.

[15] Page MJ, McKenzie JE, Bossuyt PM, Boutron I, Hoffmann TC, Mulrow CD, et al. The PRISMA 2020 statement: an updated guideline for reporting systematic reviews. Bmj. 2021;372:n71.

[16] McGowan J, Sampson M, Salzwedel DM, Cogo E, Foerster V, Lefebvre C. PRESS Peer Review of Electronic Search Strategies: 2015 Guideline Statement. J Clin Epidemiol. 2016;75:40–6.

[17] Rethlefsen ML, Kirtley S, Waffenschmidt S, Ayala AP, Moher D, Page MJ, et al. PRISMA-S: an extension to the PRISMA Statement for Reporting Literature Searches in Systematic Reviews. Syst Rev. 2021;10:39.

[18] Bramer WM, Giustini D, de Jonge GB, Holland L, Bekhuis T. De-duplication of database search results for systematic reviews in EndNote. J Med Libr Assoc. 2016;104:240–3.

[19] Whiting PF, Rutjes AW, Westwood ME, Mallett S, Deeks JJ, Reitsma JB, et al. QUADAS-2: a revised tool for the quality assessment of diagnostic accuracy studies. Ann Intern Med. 2011;155:529–36.

[20] Wolff RF, Moons KGM, Riley RD, Whiting PF, Westwood M, Collins GS, et al. PROBAST: A Tool to Assess the Risk of Bias and Applicability of Prediction Model Studies. Ann Intern Med. 2019;170:51–8.

[21] Hanley JA, McNeil BJ. The meaning and use of the area under a receiver operating characteristic (ROC) curve. Radiology. 1982;143:29–36.

[22] R Core Team. R: A language and environment for statistical computing.: R Foundation for Statistical Computing, Vienna, Austria; 2021.

[23] Viechtbauer W. Conducting meta-analyses in R with the metafor package. Journal of Statistical Software. 2010;36:1–48.

[24] Aam S, Einstad MS, Munthe-Kaas R, Lydersen S, Ihle-Hansen H, Knapskog AB, et al. Post-stroke Cognitive Impairment-Impact of Follow-Up Time and Stroke Subtype on Severity and Cognitive Profile: The Nor-COAST Study. Front Neurol. 2020;11:699.

[25] Aamodt EB, Schellhorn T, Stage E, Sanjay AB, Logan PE, Svaldi DO, et al. Predicting the Emergence of Major Neurocognitive Disorder Within Three Months After a Stroke. Frontiers in Aging Neuroscience. 2021;13.

[26] Altieri M, Di Piero V, Pasquini M, Gasparini M, Vanacore N, Vicenzini E, et al. Delayed poststroke dementia. Neurology. 2004;62:2193–7.

[27] Appel J, Potter E, Bhatia N, Shen Q, Zhao W, Greig MT, et al. Association of white matter hyperintensity measurements on brain MR imaging with cognitive status, medial temporal atrophy, and cardiovascular risk factors. AJNR Am J Neuroradiol. 2009;30:1870–6.

[28] Belathur Suresh M, Fischl B, Salat DH. Factors influencing accuracy of cortical thickness in the diagnosis of Alzheimer’s disease. Hum Brain Mapp. 2018;39:1500–15.

[29] Binzer M, Hammernik K, Rueckert D, Zimmer VA. Long-Term Cognitive Outcome Prediction in Stroke Patients Using Multi-task Learning on Imaging and Tabular Data. Cham: Springer Nature Switzerland; 2022. p. 137–48.

[30] Bordin V, Coluzzi D, Rivolta MW, Baselli G. Explainable AI Points to White Matter Hyperintensities for Alzheimer’s Disease Identification: a Preliminary Study. 2022 44th Annual International Conference of the IEEE Engineering in Medicine & Biology Society (EMBC) 2022. p. 484–7.

[31] Cajanus A, Hall A, Koikkalainen J, Solje E, Tolonen A, Urhemaa T, et al. Automatic MRI Quantifying Methods in Behavioral-Variant Frontotemporal Dementia Diagnosis. Dement Geriatr Cogn Dis Extra. 2018;8:51–9.

[32] Chan K, Fischer C, Maralani PJ, Black SE, Moody AR, Khademi A. Alzheimer’s and vascular disease classification using regional texture biomarkers in FLAIR MRI. NeuroImage: Clinical. 2023;38:103385.

[33] Chen H, Huang L, Yang D, Ye Q, Guo M, Qin R, et al. Nodal Global Efficiency in Front-Parietal Lobe Mediated Periventricular White Matter Hyperintensity (PWMH)-Related Cognitive Impairment. Front Aging Neurosci. 2019;11:347.

[34] Chen H, Sheng X, Qin R, Luo C, Li M, Liu R, et al. Aberrant White Matter Microstructure as a Potential Diagnostic Marker in Alzheimer’s Disease by Automated Fiber Quantification. Front Neurosci. 2020;14:570123.

[35] Chen H, Xu J, Lv W, Hu Z, Ke Z, Qin R, et al. Altered static and dynamic functional network connectivity related to cognitive decline in individuals with white matter hyperintensities. Behav Brain Res. 2023;451:114506.

[36] Chen Y, Sha M, Zhao X, Ma J, Ni H, Gao W, et al. Automated detection of pathologic white matter alterations in Alzheimer’s disease using combined diffusivity and kurtosis method. Psychiatry Res Neuroimaging. 2017;264:35–45.

[37] Ciulli S, Citi L, Salvadori E, Valenti R, Poggesi A, Inzitari D, et al. Prediction of Impaired Performance in Trail Making Test in MCI Patients With Small Vessel Disease Using DTI Data. IEEE J Biomed Health Inform. 2016;20:1026–33.

[38] Diciotti S, Ciulli S, Ginestroni A, Salvadori E, Poggesi A, Pantoni L, et al. Multimodal MRI classification in vascular mild cognitive impairment. 2015 37th Annual International Conference of the IEEE Engineering in Medicine and Biology Society (EMBC) 2015. p. 4278–81.

[39] Dyrba M, Ewers M, Wegrzyn M, Kilimann I, Plant C, Oswald A, et al. Robust automated detection of microstructural white matter degeneration in Alzheimer’s disease using machine learning classification of multicenter DTI data. PLoS One. 2013;8:e64925.

[40] Haller S, Bartsch A, Nguyen D, Rodriguez C, Emch J, Gold G, et al. Cerebral microhemorrhage and iron deposition in mild cognitive impairment: susceptibility-weighted MR imaging assessment. Radiology. 2010;257:764–73.

[41] Han L, Liu L, Hao Y, Zhang L. Diagnosis and Treatment Effect of Convolutional Neural Network-Based Magnetic Resonance Image Features on Severe Stroke and Mental State. Contrast Media & Molecular Imaging. 2021;2021:8947789.

[42] Jokinen H, Koikkalainen J, Laakso HM, Melkas S, Nieminen T, Brander A, et al. Global Burden of Small Vessel Disease–Related Brain Changes on MRI Predicts Cognitive and Functional Decline. Stroke. 2020;51:170–8.

[43] Joo L, Shim WH, Suh CH, Lim SJ, Heo H, Kim WS, et al. Diagnostic performance of deep learning-based automatic white matter hyperintensity segmentation for classification of the Fazekas scale and differentiation of subcortical vascular dementia. PLoS One. 2022;17:e0274562.

[44] Kandiah N, Mak E, Ng A, Huang S, Au WL, Sitoh YY, et al. Cerebral white matter hyperintensity in Parkinson’s disease: A major risk factor for mild cognitive impairment. Parkinsonism & Related Disorders. 2013;19:680–3.

[45] Lai Y, Xu L, Yao L, Wu X. Discriminative analysis of non-linear brain connectivity for leukoaraiosis with resting-state fMRI: SPIE; 2015.

[46] Lambert C, Zeestraten E, Williams O, Benjamin P, Lawrence AJ, Morris RG, et al. Identifying preclinical vascular dementia in symptomatic small vessel disease using MRI. NeuroImage: Clinical. 2018;19:925–38.

[47] Lee R, Choi H, Park K-Y, Kim J-M, Seok JW. Prediction of post-stroke cognitive impairment using brain FDG PET: deep learning-based approach. European journal of nuclear medicine and molecular imaging. 2022;49:1254–62.

[48] Li B, Zhang M, Riphagen J, Morrison Yochim K, Li B, Liu J, et al. Prediction of clinical and biomarker conformed Alzheimer’s disease and mild cognitive impairment from multi-feature brain structural MRI using age-correction from a large independent lifespan sample. NeuroImage: Clinical. 2020;28:102387.

[49] Li R, Lai Y, Zhang Y, Yao L, Wu X. Classification of Cognitive Level of Patients with Leukoaraiosis on the Basis of Linear and Non-Linear Functional Connectivity. Front Neurol. 2017;8.

[50] Liang L, Zhou P, Ye C, Yang Q, Ma T. Spatial–temporal patterns of brain disconnectome in Alzheimer’s disease. Hum Brain Mapp. 2023;44:4272–86.

[51] Lin C-J, Tu P-C, Chern C-M, Hsiao F-J, Chang F-C, Cheng H-L, et al. Connectivity Features for Identifying Cognitive Impairment in Presymptomatic Carotid Stenosis. PLOS ONE. 2014;9:e85441.

[52] Lindemer ER, Greve DN, Fischl B, Salat DH, Gomez-Isla T. White matter abnormalities and cognition in patients with conflicting diagnoses and CSF profiles. Neurology. 2018;90:e1461–e9.

[53] Ma J, Liu F, Wang Y, Ma L, Niu Y, Wang J, et al. Frequency-dependent white-matter functional network changes associated with cognitive deficits in subcortical vascular cognitive impairment. NeuroImage: Clinical. 2022;36:103245.

[54] Meng D, Hosseini AA, Simpson RJ, Shaikh Q, Tench CR, Dineen RA, et al. Lesion Topography and Microscopic White Matter Tract Damage Contribute to Cognitive Impairment in Symptomatic Carotid Artery Disease. Radiology. 2017;282:502–15.

[55] Mortamais M, Reynes C, Brickman AM, Provenzano FA, Muraskin J, Portet F, et al. Spatial distribution of cerebral white matter lesions predicts progression to mild cognitive impairment and dementia. PLoS One. 2013;8:e56972.

[56] Oppedal K, Eftestøl T, Engan K, Beyer MK, Aarsland D. Classifying dementia using local binary patterns from different regions in magnetic resonance images. Int J Biomed Imaging. 2015;2015:572567.

[57] Oppedal K, Engan K, Eftestøl T, Beyer M, Aarsland D. Classifying Alzheimer’s disease, Lewy body dementia, and normal controls using 3D texture analysis in magnetic resonance images. Biomedical Signal Processing and Control. 2017;33:19–29.

[58] Park G, Hong J, Duffy BA, Lee JM, Kim H. White matter hyperintensities segmentation using the ensemble U-Net with multi-scale highlighting foregrounds. Neuroimage. 2021;237:118140.

[59] Peters F, Villeneuve S, Belleville S. Predicting Progression to Dementia in Elderly Subjects with Mild Cognitive Impairment Using Both Cognitive and Neuroimaging Predictors. Journal of Alzheimer’s Disease. 2014;38:307–18.

[60] Provenzano FA, Muraskin J, Tosto G, Narkhede A, Wasserman BT, Griffith EY, et al. White Matter Hyperintensities and Cerebral Amyloidosis: Necessary and Sufficient for Clinical Expression of Alzheimer Disease? JAMA Neurology. 2013;70:455–61.

[61] Qin Q, Qu J, Yin Y, Liang Y, Wang Y, Xie B, et al. Unsupervised machine learning model to predict cognitive impairment in subcortical ischemic vascular disease. Alzheimers Dement. 2023;19:3327–38.

[62] Rabin JS, Neal TE, Nierle HE, Sikkes SAM, Buckley RF, Amariglio RE, et al. Multiple markers contribute to risk of progression from normal to mild cognitive impairment. NeuroImage: Clinical. 2020;28:102400.

[63] Rosano C, Aizenstein HJ, Wu M, Newman AB, Becker JT, Lopez OL, et al. Focal atrophy and cerebrovascular disease increase dementia risk among cognitively normal older adults. J Neuroimaging. 2007;17:148–55.

[64] Rosano C, Perera S, Inzitari M, Newman AB, Longstreth WT, Studenski S. Digit Symbol Substitution test and future clinical and subclinical disorders of cognition, mobility and mood in older adults. Age Ageing. 2016;45:688–95.

[65] Schouten TM, Koini M, de Vos F, Seiler S, van der Grond J, Lechner A, et al. Combining anatomical, diffusion, and resting state functional magnetic resonance imaging for individual classification of mild and moderate Alzheimer’s disease. NeuroImage: Clinical. 2016;11:46–51.

[66] Smith CD, Johnson ES, Van Eldik LJ, Jicha GA, Schmitt FA, Nelson PT, et al. Peripheral (deep) but not periventricular MRI white matter hyperintensities are increased in clinical vascular dementia compared to Alzheimer’s disease. Brain and Behavior. 2016;6:e00438.

[67] Stebbins GT, Nyenhuis DL, Wang C, Cox JL, Freels S, Bangen K, et al. Gray matter atrophy in patients with ischemic stroke with cognitive impairment. Stroke. 2008;39:785–93.

[68] Stephan BC, Tzourio C, Auriacombe S, Amieva H, Dufouil C, Alpérovitch A, et al. Usefulness of data from magnetic resonance imaging to improve prediction of dementia: population based cohort study. Bmj. 2015;350:h2863.

[69] Tang L, Wu X, Liu H, Wu F, Song R, Zhang W, et al. Individualized Prediction of Early Alzheimer’s Disease Based on Magnetic Resonance Imaging Radiomics, Clinical, and Laboratory Examinations: A 60-Month Follow-Up Study. J Magn Reson Imaging. 2021;54:1647–57.

[70] Tozer DJ, Zeestraten E, Lawrence AJ, Barrick TR, Markus HS. Texture Analysis of T1-Weighted and Fluid-Attenuated Inversion Recovery Images Detects Abnormalities That Correlate With Cognitive Decline in Small Vessel Disease. Stroke. 2018;49:1656–61.

[71] Tu MC, Huang SM, Hsu YH, Yang JJ, Lin CY, Kuo LW. Discriminating subcortical ischemic vascular disease and Alzheimer’s disease by diffusion kurtosis imaging in segregated thalamic regions. Hum Brain Mapp. 2021;42:2018–31.

[72] Twait EL, Andaur Navarro CL, Gudnason V, Hu Y-H, Launer LJ, Geerlings MI. Dementia prediction in the general population using clinically accessible variables: a proof-of-concept study using machine learning. The AGES-Reykjavik study. BMC Medical Informatics and Decision Making. 2023;23:168.

[73] Verdelho A, Madureira S, Ferro JM, Baezner H, Blahak C, Poggesi A, et al. Physical activity prevents progression for cognitive impairment and vascular dementia: results from the LADIS (Leukoaraiosis and Disability) study. Stroke. 2012;43:3331–5.

[74] Wan MD, Liu H, Liu XX, Zhang WW, Xiao XW, Zhang SZ, et al. Associations of multiple visual rating scales based on structural magnetic resonance imaging with disease severity and cerebrospinal fluid biomarkers in patients with Alzheimer’s disease. Front Aging Neurosci. 2022;14:906519.

[75] Wang J, Knol MJ, Tiulpin A, Dubost F, de Bruijne M, Vernooij MW, et al. Gray Matter Age Prediction as a Biomarker for Risk of Dementia. Proc Natl Acad Sci U S A. 2019;116:21213–8.

[76] Wang Y, Xu C, Park JH, Lee S, Stern Y, Yoo S, et al. Diagnosis and prognosis of Alzheimer’s disease using brain morphometry and white matter connectomes. Neuroimage Clin. 2019;23:101859.

[77] West NA, Windham BG, Knopman DS, Shibata DK, Coker LH, Mosley TH, Jr. Neuroimaging findings in midlife and risk of late-life dementia over 20 years of follow-up. Neurology. 2019;92:e917–e23.

[78] Williams OA, Zeestraten EA, Benjamin P, Lambert C, Lawrence AJ, Mackinnon AD, et al. Predicting Dementia in Cerebral Small Vessel Disease Using an Automatic Diffusion Tensor Image Segmentation Technique. Stroke. 2019;50:2775–82.

[79] Xie Y, Cui Z, Zhang Z, Sun Y, Sheng C, Li K, et al. Identification of Amnestic Mild Cognitive Impairment Using Multi-Modal Brain Features: A Combined Structural MRI and Diffusion Tensor Imaging Study. Journal of Alzheimer’s Disease. 2015;47:509–22.

[80] Yao M, Zhu YC, Soumaré A, Dufouil C, Mazoyer B, Tzourio C, et al. Hippocampal perivascular spaces are related to aging and blood pressure but not to cognition. Neurobiol Aging. 2014;35:2118–25.

[81] Zhang W, Li M, Zhou X, Huang C, Wan K, Li C, et al. Altered serum amyloid beta and cerebral perfusion and their associations with cognitive function in patients with subcortical ischemic vascular disease. Front Neurosci. 2022;16:993767.

[82] Zhang W, Zheng X, Li R, Liu M, Xiao W, Huang L, et al. Research on nonstroke dementia screening and cognitive function prediction model for older people based on brain atrophy characteristics. Brain Behav. 2022;12:e2726.

[83] Zhang Y, Tartaglia MC, Schuff N, Chiang GC, Ching C, Rosen HJ, et al. MRI Signatures of Brain Macrostructural Atrophy and Microstructural Degradation in Frontotemporal Lobar Degeneration Subtypes. Journal of Alzheimer’s Disease. 2013;33:431–44.

[84] Zhao J, Ding X, Du Y, Wang X, Men G. Functional connectivity between white matter and gray matter based on fMRI for Alzheimer’s disease classification. Brain and Behavior. 2019;9:e01407.

[85] Zhu W, Huang H, Yang S, Luo X, Zhu W, Xu S, et al. Dysfunctional Architecture Underlies White Matter Hyperintensities with and without Cognitive Impairment. Journal of Alzheimer’s Disease. 2019;71:461–76.

[86] Chen J, Yang J, Shen D, Wang X, Lin Z, Chen H, et al. A Predictive Model of the Progression to Alzheimer’s Disease in Patients with Mild Cognitive Impairment Based on the MRI Enlarged Perivascular Spaces. Journal of Alzheimer’s Disease. 2024;101:159–73.

[87] Chen Y, Lu P, Wu S, Yang J, Liu W, Zhang Z, et al. CD163-Mediated Small-Vessel Injury in Alzheimer’s Disease: An Exploration from Neuroimaging to Transcriptomics. Int J Mol Sci. 2024;25:2293.

[88] Chen Y, Tozer D, Li R, Li H, Tuladhar A, De Leeuw FE, et al. Improved Dementia Prediction in Cerebral Small Vessel Disease Using Deep Learning–Derived Diffusion Scalar Maps From T1. Stroke. 2024;55:2254–63.

[89] Crystal O, Maralani PJ, Black S, Fischer C, Moody AR, Khademi A. Detecting conversion from mild cognitive impairment to Alzheimer’s disease using FLAIR MRI biomarkers. Neuroimage Clin. 2023;40:103533.

[90] De Francesco S, Crema C, Archetti D, Muscio C, Reid RI, Nigri A, et al. Differential diagnosis of neurodegenerative dementias with the explainable MRI based machine learning algorithm MUQUBIA. Sci Rep. 2023;13:17355.

[91] Feng J, Hui D, Zheng Q, Guo Y, Xia Y, Shi F, et al. Automatic detection of cognitive impairment in patients with white matter hyperintensity and causal analysis of related factors using artificial intelligence of MRI. Comput Biol Med. 2024;178:108684.

[92] Keller JA, Sigurdsson S, Schmitz Abecassis B, Kant IMJ, Van Buchem MA, Launer LJ, et al. Identification of Distinct Brain MRI Phenotypes and Their Association With Long-Term Dementia Risk in Community-Dwelling Older Adults. Neurology. 2024;102:e209176.

[93] Lee M-W, Kim HW, Choe YS, Yang HS, Lee J, Lee H, et al. A multimodal machine learning model for predicting dementia conversion in Alzheimer’s disease. Scientific Reports. 2024;14:12276.

[94] Li R, Harshfield EL, Bell S, Burkhart M, Tuladhar AM, Hilal S, et al. Predicting incident dementia in cerebral small vessel disease: comparison of machine learning and traditional statistical models. Cerebral Circulation – Cognition and Behavior. 2023;5:100179.

[95] Marzi C, Scheda R, Salvadori E, Giorgio A, De Stefano N, Poggesi A, et al. Fractal dimension of the cortical gray matter outweighs other brain MRI features as a predictor of transition to dementia in patients with mild cognitive impairment and leukoaraiosis. Frontiers in Human Neuroscience. 2023;17.

[96] Shi Y, Deng J, Mao H, Han Y, Gao Q, Zeng S, et al. Macrophage Migration Inhibitory Factor as a Potential Plasma Biomarker of Cognitive Impairment in Cerebral Small Vessel Disease. ACS Omega. 2024;9:15339–49.

[97] Strain JF, Phuah C-L, Adeyemo B, Cheng K, Womack KB, McCarthy J, et al. White matter hyperintensity longitudinal morphometric analysis in association with Alzheimer disease. Alzheimer’s & Dementia. 2023;19:4488–97.

[98] Zhu XW, Liu SB, Ji CH, Liu JJ, Huang C. Machine learning-based prediction of mild cognitive impairment among individuals with normal cognitive function. Front Neurol. 2024;15.

[99] Altieri M, Di Piero V, Pasquini M, Gasparini M, Vanacore N, Vicenzini E, et al. Delayed poststroke dementia: a 4-year follow-up study. Neurology. 2004;62:2193–7.

[100] American Psychiatric Association D, American Psychiatric Association D. Diagnostic and statistical manual of mental disorders: DSM-5: American psychiatric association Washington, DC; 2013.

[101] McKhann G, Drachman D, Folstein M, Katzman R, Price D, Stadlan EM. Clinical diagnosis of Alzheimer’s disease: report of the NINCDS-ADRDA Work Group under the auspices of Department of Health and Human Services Task Force on Alzheimer’s Disease. Neurology. 1984;34:939–44.

[102] Jack Jr. CR, Bennett DA, Blennow K, Carrillo MC, Dunn B, Haeberlein SB, et al. NIA-AA Research Framework: Toward a biological definition of Alzheimer’s disease. Alzheimer’s & Dementia. 2018;14:535–62.

[103] Petersen RC, Wiste HJ, Weigand SD, Fields JA, Geda YE, Graff-Radford J, et al. NIA-AA Alzheimer’s Disease Framework: Clinical Characterization of Stages. Ann Neurol. 2021;89:1145–56.

[104] Román GC, Tatemichi TK, Erkinjuntti T, Cummings JL, Masdeu JC, Garcia JH, et al. Vascular dementia. Neurology. 1993;43:250-.

[105] Chui HC, Victoroff JI, Margolin D, Jagust W, Shankle R, Katzman R. Criteria for the diagnosis of ischemic vascular dementia proposed by the State of California Alzheimer’s Disease Diagnostic and Treatment Centers. Neurology. 1992;42:473-.

[106] Petersen RC, Smith GE, Waring SC, Ivnik RJ, Tangalos EG, Kokmen E. Mild cognitive impairment: clinical characterization and outcome. Arch Neurol. 1999;56:303–8.

[107] Petersen RC. Mild cognitive impairment as a diagnostic entity. Journal of Internal Medicine. 2004;256:183–94.

[108] Attems J, Jellinger KA. The overlap between vascular disease and Alzheimer’s disease--lessons from pathology. BMC Med. 2014;12:206.

[109] Dadar M, Manera AL, Ducharme S, Collins DL. White matter hyperintensities are associated with grey matter atrophy and cognitive decline in Alzheimer’s disease and frontotemporal dementia. Neurobiol Aging. 2022;111:54–63.

[110] Dadar M, Camicioli R, Duchesne S, Collins DL, Initiative ftAsDN. The temporal relationships between white matter hyperintensities, neurodegeneration, amyloid beta, and cognition. Alzheimer’s & Dementia: Diagnosis, Assessment & Disease Monitoring. 2020;12:e12091.

[111] Garnier-Crussard A, Bougacha S, Wirth M, Dautricourt S, Sherif S, Landeau B, et al. White matter hyperintensity topography in Alzheimer’s disease and links to cognition. Alzheimers Dement. 2022;18:422–33.

[112] Liu Y, Braidy N, Poljak A, Chan DKY, Sachdev P. Cerebral small vessel disease and the risk of Alzheimer’s disease: A systematic review. Ageing Res Rev. 2018;47:41–8.

[113] McKay E, Counts SE. Multi-Infarct Dementia: A Historical Perspective. Dement Geriatr Cogn Dis Extra. 2017;7:160–71.

[114] Kivipelto M, Helkala E-L, Laakso MP, Hänninen T, Hallikainen M, Alhainen K, et al. Midlife vascular risk factors and Alzheimer’s disease in later life: longitudinal, population based study. BMJ. 2001;322:1447.

[115] Sweeney MD, Montagne A, Sagare AP, Nation DA, Schneider LS, Chui HC, et al. Vascular dysfunction—The disregarded partner of Alzheimer’s disease. Alzheimer’s & Dementia. 2019;15:158–67.

[116] Huang W, Xia Q, Zheng F, Zhao X, Ge F, Xiao J, et al. Microglia-Mediated Neurovascular Unit Dysfunction in Alzheimer’s Disease. J Alzheimers Dis. 2023;94:S335–s54.

[117] Zimmerman B, Rypma B, Gratton G, Fabiani M. Age-related changes in cerebrovascular health and their effects on neural function and cognition: A comprehensive review. Psychophysiology. 2021;58:e13796.

[118] Nedergaard M, Goldman SA. Glymphatic failure as a final common pathway to dementia. Science. 2020;370:50–6.

[119] Laing KK, Simoes S, Baena-Caldas GP, Lao PJ, Kothiya M, Igwe KC, et al. Cerebrovascular disease promotes tau pathology in Alzheimer’s disease. Brain Commun. 2020;2:fcaa132.

[120] Backhouse EV, Boardman JP, Wardlaw JM. Cerebral Small Vessel Disease: Early-Life Antecedents and Long-Term Implications for the Brain, Aging, Stroke, and Dementia. Hypertension. 2024;81:54–74.

[121] Livingston G, Huntley J, Liu KY, Costafreda SG, Selbæk G, Alladi S, et al. Dementia prevention, intervention, and care: 2024 report of the Lancet standing Commission. The Lancet. 2024;404:572–628.

[122] Grodstein F, Leurgans SE, Capuano AW, Schneider JA, Bennett DA. Trends in Postmortem Neurodegenerative and Cerebrovascular Neuropathologies Over 25 Years. JAMA Neurology. 2023;80:370–6.

[123] Satizabal CL, Beiser AS, Chouraki V, Chêne G, Dufouil C, Seshadri S. Incidence of Dementia over Three Decades in the Framingham Heart Study. New England Journal of Medicine. 2016;374:523–32.

[124] Derby CA, Katz MJ, Lipton RB, Hall CB. Trends in Dementia Incidence in a Birth Cohort Analysis of the Einstein Aging Study. JAMA Neurol. 2017;74:1345–51.

[125] Tom SE, Phadke M, Hubbard RA, Crane PK, Stern Y, Larson EB. Association of Demographic and Early-Life Socioeconomic Factors by Birth Cohort With Dementia Incidence Among US Adults Born Between 1893 and 1949. JAMA Network Open. 2020;3:e2011094-e.

[126] Osuafor CN, Rua C, Mackinnon AD, Egle M, Benjamin P, Tozer DJ, et al. Visualisation of lenticulostriate arteries using contrast-enhanced time-of-flight magnetic resonance angiography at 7 Tesla. Scientific Reports. 2022;12:20306.

[127] Zwartbol MH, van der Kolk AG, Kuijf HJ, Witkamp TD, Ghaznawi R, Hendrikse J, et al. Intracranial vessel wall lesions on 7T MRI and MRI features of cerebral small vessel disease: The SMART-MR study. Journal of Cerebral Blood Flow & Metabolism. 2021;41:1219–28.

[128] Dencks S, Schmitz G. Ultrasound localization microscopy. Zeitschrift für Medizinische Physik. 2023;33:292–308.

[129] Song P, Rubin JM, Lowerison MR. Super-resolution ultrasound microvascular imaging: Is it ready for clinical use? Zeitschrift für Medizinische Physik. 2023;33:309–23.

[130] Desailly Y, Pierre J, Couture O, Tanter M. Resolution limits of ultrafast ultrasound localization microscopy. Phys Med Biol. 2015;60:8723–40.

[131] Hillman EM. Coupling mechanism and significance of the BOLD signal: a status report. Annu Rev Neurosci. 2014;37:161–81.

[132] Li K, Fu Z, Luo X, Zeng Q, Huang P, Zhang M, et al. The Influence of Cerebral Small Vessel Disease on Static and Dynamic Functional Network Connectivity in Subjects Along Alzheimer’s Disease Continuum. Brain Connect. 2021;11:189–200.

[133] Ru X, He K, Lyu B, Li D, Xu W, Gu W, et al. Multimodal neuroimaging with optically pumped magnetometers: A simultaneous MEG-EEG-fNIRS acquisition system. NeuroImage. 2022;259:119420.

[134] Smith EE, Biessels GJ, De Guio F, de Leeuw FE, Duchesne S, Düring M, et al. Harmonizing brain magnetic resonance imaging methods for vascular contributions to neurodegeneration. Alzheimers Dement (Amst). 2019;11:191–204.

[135] Bordin V, Bertani I, Mattioli I, Sundaresan V, McCarthy P, Suri S, et al. Integrating large-scale neuroimaging research datasets: Harmonisation of white matter hyperintensity measurements across Whitehall and UK Biobank datasets. Neuroimage. 2021;237:118189.

[136] Coenen M, Kuijf HJ, Huenges Wajer IMC, Duering M, Wolters FJ, Fletcher EF, et al. Strategic white matter hyperintensity locations for cognitive impairment: A multicenter lesion-symptom mapping study in 3525 memory clinic patients. Alzheimer’s & Dementia. 2023;19:2420–32.

[137] Lee S, Zimmerman ME, Narkhede A, Nasrabady SE, Tosto G, Meier IB, et al. White matter hyperintensities and the mediating role of cerebral amyloid angiopathy in dominantly-inherited Alzheimer’s disease. PLOS ONE. 2018;13:e0195838.

[138] Thanprasertsuk S, Martinez-Ramirez S, Pontes-Neto OM, Ni J, Ayres A, Reed A, et al. Posterior white matter disease distribution as a predictor of amyloid angiopathy. Neurology. 2014;83:794–800.

[139] Renard D, Tatu L, Demattei C, Hirtz C, Lehmann S, Thouvenot E. Characterizing Deep White Matter Hyperintensities in Patients with Symptomatic Isolated Cortical Superficial Siderosis. J Stroke Cerebrovasc Dis. 2017;26:465–9.

[140] Alber J, Arthur E, Goldfarb D, Drake J, Boxerman JL, Silver B, et al. The relationship between cerebral and retinal microbleeds in cerebral amyloid angiopathy (CAA): A pilot study. Journal of the Neurological Sciences. 2021;423:117383.

[141] Ii Y, Ishikawa H, Matsuyama H, Shindo A, Matsuura K, Yoshimaru K, et al. Hypertensive Arteriopathy and Cerebral Amyloid Angiopathy in Patients with Cognitive Decline and Mixed Cerebral Microbleeds. J Alzheimer’s Dis. 2020;78:1765–74.

[142] Bouvy WH, van Veluw SJ, Kuijf HJ, Zwanenburg JJ, Kappelle JL, Luijten PR, et al. Microbleeds colocalize with enlarged juxtacortical perivascular spaces in amnestic mild cognitive impairment and early Alzheimer’s disease: A 7 Tesla MRI study. Journal of Cerebral Blood Flow & Metabolism. 2020;40:739–46.

[143] Weller RO, Hawkes CA, Kalaria RN, Werring DJ, Carare RO. White Matter Changes in Dementia: Role of Impaired Drainage of Interstitial Fluid. Brain Pathology. 2015;25:63–78.

[144] Xiong L, Boulouis G, Charidimou A, Roongpiboonsopit D, Jessel MJ, Pasi M, et al. Dementia incidence and predictors in cerebral amyloid angiopathy patients without intracerebral hemorrhage. Journal of Cerebral Blood Flow & Metabolism. 2018;38:241–9.

[145] Sin M-K, Zamrini E, Ahmed A, Nho K, Hajjar I. Anti-Amyloid Therapy, AD, and ARIA: Untangling the Role of CAA. Journal of Clinical Medicine. 2023;12:6792.

[146] Beach TG, Sue LI, Scott S, Intorcia AJ, Walker JE, Arce RA, et al. Cerebral white matter rarefaction has both neurodegenerative and vascular causes and may primarily be a distal axonopathy. J Neuropathol Exp Neurol. 2023;82:457–66.

[147] Sperling R, Salloway S, Brooks DJ, Tampieri D, Barakos J, Fox NC, et al. Amyloid-related imaging abnormalities in patients with Alzheimer’s disease treated with bapineuzumab: a retrospective analysis. Lancet Neurol. 2012;11:241–9.

[148] Charidimou A, Boulouis G, Frosch MP, Baron JC, Pasi M, Albucher JF, et al. The Boston criteria version 2.0 for cerebral amyloid angiopathy: a multicentre, retrospective, MRI-neuropathology diagnostic accuracy study. Lancet Neurol. 2022;21:714–25.

[149] Jo T, Nho K, Saykin AJ. Deep Learning in Alzheimer’s Disease: Diagnostic Classification and Prognostic Prediction Using Neuroimaging Data. Frontiers in Aging Neuroscience. 2019;11.

[150] Reith FH, Mormino EC, Zaharchuk G. Predicting future amyloid biomarkers in dementia patients with machine learning to improve clinical trial patient selection. Alzheimer’s & Dementia: Translational Research & Clinical Interventions. 2021;7:e12212.

[151] Beshir SA, Aadithsoorya AM, Parveen A, Goh SSL, Hussain N, Menon VB. Aducanumab Therapy to Treat Alzheimer’s Disease: A Narrative Review. International Journal of Alzheimer’s Disease. 2022;2022:9343514.

[152] Bates DW, Levine D, Syrowatka A, Kuznetsova M, Craig KJT, Rui A, et al. The potential of artificial intelligence to improve patient safety: a scoping review. npj Digital Medicine. 2021;4:54.

[153] Yin J, Ngiam KY, Teo HH. Role of Artificial Intelligence Applications in Real-Life Clinical Practice: Systematic Review. J Med Internet Res. 2021;23:e25759.

[154] Choudhury A, Asan O. Role of Artificial Intelligence in Patient Safety Outcomes: Systematic Literature Review. JMIR Med Inform. 2020;8:e18599.

[155] Collins GS, Moons KGM, Dhiman P, Riley RD, Beam AL, Van Calster B, et al. TRIPOD+AI statement: updated guidance for reporting clinical prediction models that use regression or machine learning methods. Bmj. 2024;385:e078378.

[156] Chekroud AM, Hawrilenko M, Loho H, Bondar J, Gueorguieva R, Hasan A, et al. Illusory generalizability of clinical prediction models. Science. 2024;383:164–7.

[157] Robinson JL, Xie SX, Baer DR, Suh E, Van Deerlin VM, Loh NJ, et al. Pathological combinations in neurodegenerative disease are heterogeneous and disease-associated. Brain. 2023;146:2557–69.

[158] Rahimi J, Kovacs GG. Prevalence of mixed pathologies in the aging brain. Alzheimers Res Ther. 2014;6:82.

[159] Mielke MM. Sex and Gender Differences in Alzheimer’s Disease Dementia. Psychiatr Times. 2018;35:14–7.

[160] Lohner V, Pehlivan G, Sanroma G, Miloschewski A, Schirmer MD, Stöcker T, et al. Relation Between Sex, Menopause, and White Matter Hyperintensities: The Rhineland Study. Neurology. 2022;99:e935–e43.

[161] Shiekh SI, Cadogan SL, Lin LY, Mathur R, Smeeth L, Warren-Gash C. Ethnic Differences in Dementia Risk: A Systematic Review and Meta-Analysis. J Alzheimers Dis. 2021;80:337–55.

[162] Mayeda ER, Glymour MM, Quesenberry CP, Whitmer RA. Inequalities in dementia incidence between six racial and ethnic groups over 14 years. Alzheimer’s & Dementia. 2016;12:216–24.

[163] Hudomiet P, Hurd MD, Rohwedder S. Trends in inequalities in the prevalence of dementia in the United States. Proceedings of the National Academy of Sciences. 2022;119:e2212205119.

[164] Deckers K, Cadar D, van Boxtel MPJ, Verhey FRJ, Steptoe A, Köhler S. Modifiable Risk Factors Explain Socioeconomic Inequalities in Dementia Risk: Evidence from a Population-Based Prospective Cohort Study. J Alzheimer’s Dis. 2019;71:549–57.

[165] Sounderajah V, Ashrafian H, Rose S, Shah NH, Ghassemi M, Golub R, et al. A quality assessment tool for artificial intelligence-centered diagnostic test accuracy studies: QUADAS-AI. Nature Medicine. 2021;27:1663–5.

