## Supplementary Material for "Machine learning applications in vascular neuroimaging for the diagnosis and prognosis of cognitive impairment and dementia: a systematic review and meta-analysis"

### PRISMA checklist

| Section and Topic | Item # | Checklist item | Location where item is reported |
|---|---|---|---|
| **TITLE** | | | |
| Title | 1 | Identify the report as a systematic review. | p 1 |
| **ABSTRACT** | | | |
| Abstract | 2 | See the PRISMA 2020 for Abstracts checklist. | p 3 |
| **INTRODUCTION** | | | |
| Rationale | 3 | Describe the rationale for the review in the context of existing knowledge. | pp 5-6 |
| Objectives | 4 | Provide an explicit statement of the objective(s) or question(s) the review addresses. | p 6 |
| **METHODS** | | | |
| Eligibility criteria | 5 | Specify the inclusion and exclusion criteria for the review and how studies were grouped for the syntheses. | pp 7-9 |
| Information sources | 6 | Specify all databases, registers, websites, organisations, reference lists and other sources searched or consulted to identify studies. Specify the date when each source was last searched or consulted. | p 7 |
| Search strategy | 7 | Present the full search strategies for all databases, registers and websites, including any filters and limits used. | supplements |
| Selection process | 8 | Specify the methods used to decide whether a study met the inclusion criteria of the review, including how many reviewers screened each record and each report retrieved, whether they worked independently, and if applicable, details of automation tools used in the process. | pp 7-9 |
| Data collection process | 9 | Specify the methods used to collect data from reports, including how many reviewers collected data from each report, whether they worked independently, any processes for obtaining or confirming data from study investigators, and if applicable, details of automation tools used in the process. | p 9 |
| Data items | 10a | List and define all outcomes for which data were sought. Specify whether all results that were compatible with each outcome domain in each study were sought (e.g. for all measures, time points, analyses), and if not, the methods used to decide which results to collect. | pp 9-10 |
| | 10b | List and define all other variables for which data were sought (e.g. participant and intervention characteristics, funding sources). Describe any assumptions made about any missing or unclear information. | pp 9-10 |
| Study risk of bias assessment | 11 | Specify the methods used to assess risk of bias in the included studies, including details of the tool(s) used, how many reviewers assessed each study and whether they worked independently, and if applicable, details of automation tools used in the process. | p 9 |
| Effect measures | 12 | Specify for each outcome the effect measure(s) (e.g. risk ratio, mean difference) used in the synthesis or presentation of results. | pp 9-10 |
| Synthesis methods | 13a | Describe the processes used to decide which studies were eligible for each synthesis (e.g. tabulating the study intervention characteristics and comparing against the planned groups for each synthesis (item #5)). | pp 7-8 |
| | 13b | Describe any methods required to prepare the data for presentation or synthesis, such as handling of missing summary statistics, or data conversions. | pp 9-10 |
| | 13c | Describe any methods used to tabulate or visually display results of individual studies and syntheses. | N/A |

| Section and Topic | Item # | Checklist item | Location where item is reported |
|---|---|---|---|
| | 13d | Describe any methods used to synthesize results and provide a rationale for the choice(s). If meta-analysis was performed, describe the model(s), method(s) to identify the presence and extent of statistical heterogeneity, and software package(s) used. | pp 9-10 |
| | 13e | Describe any methods used to explore possible causes of heterogeneity among study results (e.g. subgroup analysis, meta-regression). | pp 9-10 |
| | 13f | Describe any sensitivity analyses conducted to assess robustness of the synthesized results. | pp 9-10 |
| Reporting bias assessment | 14 | Describe any methods used to assess risk of bias due to missing results in a synthesis (arising from reporting biases). | N/A |
| Certainty assessment | 15 | Describe any methods used to assess certainty (or confidence) in the body of evidence for an outcome. | pp 9-10 |
| **RESULTS** | | | |
| Study selection | 16a | Describe the results of the search and selection process, from the number of records identified in the search to the number of studies included in the review, ideally using a flow diagram. | pp 10-11 |
| | 16b | Cite studies that might appear to meet the inclusion criteria, but which were excluded, and explain why they were excluded. | N/A |
| Study characteristics | 17 | Cite each included study and present its characteristics. | pp 10-11 |
| Risk of bias in studies | 18 | Present assessments of risk of bias for each included study. | p 24, supplements |
| Results of individual studies | 19 | For all outcomes, present, for each study: (a) summary statistics for each group (where appropriate) and (b) an effect estimate and its precision (e.g. confidence/credible interval), ideally using structured tables or plots. | supplements |
| Results of syntheses | 20a | For each synthesis, briefly summarise the characteristics and risk of bias among contributing studies. | supplements |
| | 20b | Present results of all statistical syntheses conducted. If meta-analysis was done, present for each the summary estimate and its precision (e.g. confidence/credible interval) and measures of statistical heterogeneity. If comparing groups, describe the direction of the effect. | pp 22-24 |
| | 20c | Present results of all investigations of possible causes of heterogeneity among study results. | pp 22-24 |
| | 20d | Present results of all sensitivity analyses conducted to assess the robustness of the synthesized results. | pp 23-24 |
| Reporting biases | 21 | Present assessments of risk of bias due to missing results (arising from reporting biases) for each synthesis assessed. | N/A |
| Certainty of evidence | 22 | Present assessments of certainty (or confidence) in the body of evidence for each outcome assessed. | supplements |
| **DISCUSSION** | | | |
| Discussion | 23a | Provide a general interpretation of the results in the context of other evidence. | pp 25-28 |
| | 23b | Discuss any limitations of the evidence included in the review. | pp 28-31 |
| | 23c | Discuss any limitations of the review processes used. | pp 31-32 |
| | 23d | Discuss implications of the results for practice, policy, and future research. | pp 26-30 |
| **OTHER INFORMATION** | | | |
| Registration and protocol | 24a | Provide registration information for the review, including register name and registration number, or state that the review was not registered. | p 7 |
| | 24b | Indicate where the review protocol can be accessed, or state | p 7 |

| Section and Topic | Item # | Checklist item | Location where item is reported |
|---|---|---|---|
| | | that a protocol was not prepared. | |
| | 24c | Describe and explain any amendments to information provided at registration or in the protocol. | N/A |
| Support | 25 | Describe sources of financial or non-financial support for the review, and the role of the funders or sponsors in the review. | pp 34-35 |
| Competing interests | 26 | Declare any competing interests of review authors. | p 35 |
| Availability of data, code and other materials | 27 | Report which of the following are publicly available and where they can be found: template data collection forms; data extracted from included studies; data used for all analyses; analytic code; any other materials used in the review. | supplements |

#### Supplementary Box 1 – Neuroimaging features of cerebral small vessel disease

We provide an overview of the neuroimaging features of cerebral small vessel disease (CSVD) employed in the reviewed papers. For more detailed explanations, we direct readers to [1].

##### A. Lesions

**Cerebral white matter hyperintensities of presumed vascular origin** (WMH) are regions in the white matter that appear hyperintense on T2-weighted or fluid-attenuated inversion recovery (FLAIR) sequences and hypointense on T1-weighted images and computed tomography. Histopathology suggests that WMH include demyelination, axonal loss, and gliosis.

**Cerebral microbleeds** are focal brain haemorrhages likely caused by damage to small cerebral blood vessels and are associated with cerebral amyloid angiopathy. On T2*-weighted or susceptibility-weighted magnetic resonance imaging (SWI) sequences, they appear as small, round, hypointense spots.

**Lacunes** are ovoidal, fluid-filled cavities commonly found within the basal ganglia, thalamus or deep white matter. On neuroimaging, lacunes have a similar intensity profile to that of cerebrospinal fluid, appearing hyperintense on T2-weighted imaging and hypointense on T1-weighted or FLAIR images.

**Perivascular spaces** are fluid-filled cavities that surround small perforating blood vessels. On T1-weighted and FLAIR imaging, they appear hypointense, while on T2-weighted imaging, they appear hyperintense. Although their origin remains unclear, perivascular spaces may be related to neurotoxic or metabolic waste clearance.

**Strokes** are caused by disrupted blood flow to brain regions based on either a blocked (ischemia) or ruptured (haemorrhage) cerebral artery. Diffusion-weighted imaging (DWI) is the most sensitive MRI sequence for detecting acute ischemic strokes, as these appear hyperintense with respect to other structures. Strokes cause chronic structural changes that remain visible on T2-weighted MRI.

##### B. Non-lesion outcomes

Non-lesion outcomes capture the effects of CSVD that do not result in a discrete lesion. This involves computing regional statistics of specific measurements, such as the degree of free water in the white matter. Non-lesion outcomes include atrophy (although volume loss is not specific to CSVD), connectivity, microstructural integrity, and diffusion-based image metrics.

**Diffusion-based markers** enable the assessment of white matter microstructure. Examples of these markers include fractional anisotropy, along with axial, radial, and mean diffusivity. Fractional anisotropy and mean diffusivity provide information about the average and relative degree of water diffusion within a voxel, while axial and radial diffusivity measure water diffusion along specific directions. DWI signatures derived from white matter can be used to assess white matter integrity and axonal injury. A particular type of diffusion-based marker is kurtosis, which reflects deviations in water diffusion from the standard pattern seen in unrestricted, random diffusion.

**Connectivity** describes how brain regions are associated and interact with each other. Structural and functional connectivity are types of connectivity measurements. Structural connectivity reflects how different brain regions are physically connected by white matter tracts, while functional connectivity measures activity that might be correlated between regions during specific tasks or at rest.

#### Supplementary Box 2 — Machine learning methods

We provide an overview of the machine learning (ML) algorithms employed in the reviewed papers. In brief, ML is a branch of artificial intelligence dedicated to the study and development of algorithms that can learn patterns from data. These algorithms leverage the acquired knowledge to make predictions or decisions on new, unseen data without the need for explicit step-by-step programming. We categorised the identified ML algorithms into supervised learning, unsupervised learning, and advanced techniques. For more detailed explanations, we direct readers to [2, 3].

##### A. Supervised learning

Supervised learning algorithms use labelled input-output pairs (e.g., input: neuroimaging measures; output: dementia diagnosis) to infer relationships and make predictions on new data. Regression, instance-based methods, decision trees, discriminant analysis, Bayesian approaches, and ensemble algorithms belong to this category.

**Regression** algorithms model the relationship between dependent and independent variables by fitting a line, curve, or hyperplane that minimises prediction errors. Linear, logistic, and Cox regressions are used to predict continuous, categorical, and survival outcomes, respectively. They might be augmented with regularisation techniques to select important variables, prevent overfitting, and generally improve generalisability to unseen data, e.g., least absolute shrinkage and selection operator (LASSO), ridge regression, and elastic net.

**Instance-based** algorithms store specific training instances and compare them with new data to make predictions. A popular algorithm of this type is support vector machine (SVM), which finds the optimal hyperplane in a feature space for class separation based on the training instances closest to it from each class, known as the “support vectors”. Another subcategory of methods is learning vector quantization (LVQ), which learns a prototype for each class and makes predictions on new data based on its distance to each prototype. Depending on the specific form of distance metric, there are multiple types of LVQ, including generalised matrix LVQ (GMLVQ) and generalised relevance LVQ (GRLVQ).

**Decision tree** algorithms recursively split training data based on feature values to build tree-like structures, with internal nodes representing splitting rules and leaf nodes representing outcomes. Basic decision trees set a threshold for a single feature at each internal node, whereas functional trees allow functional models, such as linear regression, to set thresholds at each node. These algorithms may be used for both classification and regression.

**Discriminant analysis** algorithms are commonly employed for both classification and dimensionality reduction**.** A popular form is the linear discriminant analysis (LDA), which assumes that data points from each class follow Gaussian distributions with identical covariance, thus forming linear class boundaries. Quadratic discriminant analysis (QDA) extends this by allowing different covariance matrices for difference classes, thus forming quadratic class boundaries. Another approach is distance discriminant analysis (DDA), which classifies samples based on their distances to predefined class representatives.

**Bayesian algorithms**, based on Bayes' theorem, incorporate prior beliefs about outcome distributions into predictions. A popular method is the naive Bayes classifier, which calculates the posterior probability for each class by combining the prior probability with the likelihood of features, then assigns the class with the highest probability. A Gaussian Process (GP) is another Bayesian method that provides a flexible way to model distributions over functions, meaning that instead of assuming a fixed form of a function, it assumes a distribution over a range of possible functions that fit the data.

**Ensemble algorithms** combine multiple classifiers aiming to boost classification accuracy, stability, and robustness. By aggregating the predictions of various models, ensemble algorithms seek to correct for weaknesses of individual classifiers, reduce variance, and improve overall model performance. Random forests, Gradient boosting, and XGBoost, which typically combine weak classifiers like decision trees, are examples of this family of algorithms.

#### B. Unsupervised learning

Unlike supervised learning, unsupervised learning operates on unlabelled data to discover underlying relationships. **Clustering algorithms** are a subset of unsupervised learning strategies that organise unlabelled data into groups based on similarities. K-means, for instance, assigns data points to the nearest cluster, recalculates centroids, and repeats until assignments stabilise. In addition, hierarchical clustering is a method of cluster analysis that organises data into a nested sequence of clusters, represented as a tree, by progressively grouping similar items based on their similarity or proximity.

#### C. Advances algorithms

Advanced algorithms use sophisticated machine learning techniques to tackle extremely complex challenges and enhance model performance. **Deep learning**, inspired by biological neural networks, mimics biological learning processes through nodes (neurons) and connections (synapses). These connections strengthen in response to correlated outputs, analogous to how biological systems learn and form memories. Among the various deep learning architectures, convolutional neural networks (CNNs) are particularly suitable for image processing. CNNs preserve spatial relationships in images, mimicking how the human visual cortex processes visual inputs. They use convolutional filters to extract and analyse features such as edges and textures from images. This functionality is essential for tasks like image classification and segmentation, making CNNs highly effective in medical imaging for identifying and categorising features in anatomical structures.

### Search strategy

The search strategies used in each database are documented in full below.

**Medline (via Ovid)**

(dementia or alzheimer* or "lewy bod*" or "frontotemporal lobar degenerat* OR frontotemporal dementia" or "progressive supranuclear palsy" or huntington* or "corticobasal degeneration" or "vascular cognitive impairment" or "Cerebral Autosomal Dominant Arteriopathy with Subcortical Infarcts and Leukoencephalopathy" or CADASIL or "Cerebral autosomal recessive arteriopathy with subcortical infarcts and leukoencephalopathy" or CARASIL or "mild cognitive impairment" or "vascular parkinsonism" or "neurocognitive disorder*" or "cerebral amyloid angiopathy" or "small vessel disease*" or "cerebrovascular disease*" or "cerebrovascular damage" or "population-based" or "community-based" or "community-dwelling" or "independent living").ti,ab,kw. or Alzheimer Disease/ or Huntington Disease/ or Lewy Bodies/ or Frontotemporal Lobar Degeneration/ or corticobasal degeneration/ or dementia/ or neurocognitive disorders/ or neurodegenerative diseases/ or cerebral amyloid angiopathy/ or Cerebral Small Vessel Diseases/ or cerebrovascular disorders/ or independent living/

AND

("white matter hyperintensit*" or "white matter disease*" or "white matter lesion*" or leukoaraiosis* or lacune* or "lacunar infarct*" or "small subcortical infarct*" or "perivascular space*" or "virchow robin space*" or microbleed* or microinfarct* or Leukoencephalopath* or "cerebrovascular damage" or haemorrhag* or hemorrhag* or "cerebrovascular accident" or stroke).ti,ab,kw. or leukoaraiosis/ or Leukoencephalopathies/

AND

("Artificial Intelligence" or "neural network*" or "deep learning" or "machine learning" or "computer reasoning" or "machine intelligence" or "ridge regression" or "least absolute shrinkage and selection operator" or LASSO or "elastic net" or classifier or "support vector machine*" or "random forest*" or "reinforcement learning" or boost* or "nearest-neighbo*" or markov or clustering or "cluster analysis" or convolution or classification or "decision support system*" or automl or ensemble or unsupervi* or supervi* or "semi-supervi*" or "weakly supervi*" or "dimensionality reduction").ti,ab,kw. or Artificial Intelligence/ or Machine Learning/ or Deep Learning/ or Neural networks, computer/ or support vector machine/ or Markov chains/ or cluster analysis/

AND

(Neuroimag* or "Magnetic Resonance Imag*" or "Positron Emission Tomography" or "structural imaging" or "fluid attenuated inversion recovery" or "functional imaging" or "arterial spin labelling" or "dynamic imaging" or "DCE-MRI" or angiograph* or "computed tomography" or "CT perfusion" or "diffusion weighted imaging" or "proton density").ti,ab,kw. or Neuroimaging/ or Magnetic resonance imaging/ or Positron-emission tomography/ or angiography/ or Tomography, X-ray computed/ or diffusion magnetic resonance imaging/

**Embase (via Ovid)**

(dementia or alzheimer* or "lewy bod*" or "frontotemporal lobar degenerat* OR frontotemporal dementia" or "progressive supranuclear palsy" or huntington* or "corticobasal degeneration" or "vascular cognitive impairment" or "Cerebral Autosomal Dominant Arteriopathy with Subcortical Infarcts and Leukoencephalopathy" or CADASIL or "Cerebral autosomal recessive arteriopathy with subcortical infarcts and leukoencephalopathy" or CARASIL or "mild cognitive impairment" or "vascular parkinsonism" or "neurocognitive disorder*" or "cerebral amyloid angiopathy" or "small vessel disease*" or "cerebrovascular disease*" or "cerebrovascular damage" or "population-based" or "community-based" or "community-dwelling" or "independent living").ti,ab,kw. or dementia/ or alzheimer disease/ or lewy body/ or frontotemporal dementia/ or progressive supranuclear palsy/ or huntington chorea/ or corticobasal degeneration/ or CADASIL/ or mild cognitive impairment/ or vascular parkinsonism/ or "disorders of higher cerebral function"/ or cerebrovascular disease/ or community dwelling person/ or independent living/

AND

("white matter hyperintensit*" or "white matter disease*" or "white matter lesion*" or leukoaraiosis* or lacune* or "lacunar infarct*" or "small subcortical infarct*" or "perivascular space*" or "virchow robin space*" or microbleed* or microinfarct* or Leukoencephalopath* or "cerebrovascular damage" or haemorrhag* or hemorrhag* or "cerebrovascular accident" or stroke).ti,ab,kw. or white matter lesion/ or leukoaraiosis/ or lacunar infarction/ or perivascular space/ or lacunar stroke/ or Leukoencephalopathy/ or cerebrovascular accident/

AND

("Artificial Intelligence" or "neural network*" or "deep learning" or "machine learning" or "computer reasoning" or "machine intelligence" or "ridge regression" or "least absolute shrinkage and selection operator" or LASSO or "elastic net" or classifier or "support vector machine*" or "random forest*" or "reinforcement learning" or boost* or "nearest-neighbo*" or markov or clustering or "cluster analysis" or convolution or classification or "decision support system*" or automl or ensemble or unsupervi* or supervi* or "semi-supervi*" or "weakly supervi*" or "dimensionality reduction").ti,ab,kw. or artificial intelligence/ or artificial neural network/ or deep learning/ or machine learning/ or ridge regression/ or "least absolute shrinkage and selection operator"/ or support vector machine/ or random forest/ or k nearest neighbor/ or markov chain/ or markov decision process/ or decision support system/ or dimensionality reduction/

AND

(Neuroimag* or "Magnetic Resonance Imag*" or "Positron Emission Tomography" or "structural imaging" or "fluid attenuated inversion recovery" or "functional imaging" or "arterial spin labelling" or "dynamic imaging" or "DCE-MRI" or angiograph* or "computed tomography" or "CT perfusion" or "diffusion weighted imaging" or "proton density").ti,ab,kw. or Positron Emission Tomography/ or functional magnetic resonance imaging/ or neuroimaging/ or fluid attenuated inversion recovery imaging/ or functional magnetic resonance imaging/ or arterial spin labelling/ or angiography/ or brain angiography/ or computer assisted tomography/ or diffusion weighted imaging/

**Cochrane Library**

(dementia or alzheimer* or "lewy bod*" or "frontotemporal lobar degenerat* OR frontotemporal dementia" or "progressive supranuclear palsy" or huntington* or "corticobasal degeneration" or "vascular cognitive impairment" or "Cerebral Autosomal Dominant Arteriopathy with Subcortical Infarcts and Leukoencephalopathy" or CADASIL or "Cerebral autosomal recessive arteriopathy with subcortical infarcts and leukoencephalopathy" or CARASIL or "mild cognitive impairment" or "vascular parkinsonism" or "neurocognitive disorder*" or "cerebral amyloid angiopathy" or "small vessel disease*" or "cerebrovascular disease*" or "cerebrovascular damage" or "population-based" or "community-based" or "community-dwelling" or "independent living"):ti,ab,kw. or MeSH descriptor: [Alzheimer Disease] this term only or MeSH descriptor: [Huntington Disease] this term only or MeSH descriptor: [Lewy Bodies] this term only or MeSH descriptor: [Frontotemporal Lobar Degeneration] this term only or MeSH descriptor: [corticobasal degeneration] this term only or MeSH descriptor: [dementia] this term only or MeSH descriptor: [neurocognitive disorders] this term only or MeSH descriptor: [neurodegenerative diseases] this term only or MeSH descriptor: [cerebral amyloid angiopathy] this term only or MeSH descriptor: [Cerebral Small Vessel Diseases] this term only or MeSH descriptor: [cerebrovascular disorders] this term only or MeSH descriptor: [independent living] this term only

AND

("white matter hyperintensit*" or "white matter disease*" or "white matter lesion*" or leukoaraiosis* or lacune* or "lacunar infarct*" or "small subcortical infarct*" or "perivascular space*" or "virchow robin space*" or microbleed* or microinfarct* or Leukoencephalopath* or "cerebrovascular damage" or haemorrhag* or hemorrhag* or "cerebrovascular accident" or stroke):ti,ab,kw or MeSH descriptor: [leukoaraiosis] this term only or MeSH descriptor: [Leukoencephalopathies] this term only

AND

("Artificial Intelligence" or "neural network*" or "deep learning" or "machine learning" or "computer reasoning" or "machine intelligence" or "ridge regression" or "least absolute shrinkage and selection operator" or LASSO or "elastic net" or classifier or "support vector machine*" or "random forest*" or "reinforcement learning" or boost* or "nearest-neighbo*" or markov or clustering or "cluster analysis" or convolution or classification or "decision support system*" or automl or ensemble or unsupervi* or supervi* or "semi-supervi*" or "weakly supervi*" or "dimensionality reduction"):ti,ab,kw or MeSH descriptor: [Artificial Intelligence] this term only or MeSH descriptor: [Machine Learning] this term only or MeSH descriptor: [Deep Learning] this term only or MeSH descriptor: [Neural networks, computer] this term only or MeSH descriptor: [support vector machine] this term only or MeSH descriptor: [Markov chains] this term only or MeSH descriptor: [cluster analysis] this term only

AND

(Neuroimag* or "Magnetic Resonance Imag*" or "Positron Emission Tomography" or "structural imaging" or "fluid attenuated inversion recovery" or "functional imaging" or "arterial spin labelling" or "dynamic imaging" or "DCE-MRI" or angiograph* or "computed tomography" or "CT perfusion" or "diffusion weighted imaging" or "proton density"):ti,ab,kw. or MeSH descriptor: [Neuroimaging] this term only or MeSH descriptor: [Magnetic resonance imaging] this term only or MeSH descriptor: [Positron-emission tomography] this term only or MeSH descriptor: [angiography] this term only or MeSH descriptor: [Tomography, X-ray computed] this term only or MeSH descriptor: [diffusion magnetic resonance imaging] this term only

**Emcare (via Ovid)**

(dementia or alzheimer* or "lewy bod*" or "frontotemporal lobar degenerat* OR frontotemporal dementia" or "progressive supranuclear palsy" or huntington* or "corticobasal degeneration" or "vascular cognitive impairment" or "Cerebral Autosomal Dominant Arteriopathy with Subcortical Infarcts and Leukoencephalopathy" or CADASIL or "Cerebral autosomal recessive arteriopathy with subcortical infarcts and leukoencephalopathy" or CARASIL or "mild cognitive impairment" or "vascular parkinsonism" or "neurocognitive disorder*" or "cerebral amyloid angiopathy" or "small vessel disease*" or "cerebrovascular disease*" or "cerebrovascular damage" or "population-based" or "community-based" or "community-dwelling" or "independent living").ti,ab,kw. or Alzheimer Disease/ or Huntington Chorea/ or Lewy Body/ or Frontotemporal Dementia/ or corticobasal degeneration/ or dementia/ or degenerative diseases/ or CADASIL/ or mild cognitive impairment/ or progressive supranuclear palsy/ or vascular parkinsonism/ or vascular amyloidosis/ or cerebrovascular disease/ or community dwelling person or independent living/
AND
("white matter hyperintensit*" or "white matter disease*" or "white matter lesion*" or leukoaraiosis* or lacune* or "lacunar infarct*" or "small subcortical infarct*" or "perivascular space*" or "virchow robin space*" or microbleed* or microinfarct* or Leukoencephalopath* or "cerebrovascular damage" or haemorrhag* or hemorrhag* or "cerebrovascular accident" or stroke).ti,ab,kw. or white matter lesion/ or leukoaraiosis/ or lacunar infarction/ or perivascular space or Leukoencephalopathy/ or cerebrovascular accident/

AND

("Artificial Intelligence" or "neural network*" or "deep learning" or "machine learning" or "computer reasoning" or "machine intelligence" or "ridge regression" or "least absolute shrinkage and selection operator" or LASSO or "elastic net" or classifier or "support vector machine*" or "random forest*" or "reinforcement learning" or boost* or "nearest-neighbo*" or markov or clustering or "cluster analysis" or convolution or classification or "decision support system*" or automl or ensemble or unsupervi* or supervi* or "semi-supervi*" or "weakly supervi*" or "dimensionality reduction").ti,ab,kw. or Artificial Intelligence/ or artificial neural network/ or Machine Learning/ or ridge regression/ or Deep Learning/ or "least absolute shrinkage and selection operator"/ or support vector machine/ or random forest/ or k-nearest neighbor/ or markov chain/ or clustering algorithm/ or dimensionality reduction/

AND

(Neuroimag* or "Magnetic Resonance Imag*" or "Positron Emission Tomography" or "structural imaging" or "fluid attenuated inversion recovery" or "functional imaging" or "arterial spin labelling" or "dynamic imaging" or "DCE-MRI" or angiograph* or "computed tomography" or "CT perfusion" or "diffusion weighted imaging" or "proton density").ti,ab,kw. or Neuroimaging/ or functional magnetic resonance imaging/ or Positron emission tomography/ or angiography/ or computer assisted tomography/ or diffusion magnetic resonance imaging/ or arterial spin labelling/ or angiography/ or diffusion weighted imaging/

**Cinahl (via Ebscohost)**

TI (dementia or alzheimer* or "lewy bod*" or "frontotemporal lobar degenerat* OR frontotemporal dementia" or "progressive supranuclear palsy" or huntington* or "corticobasal degeneration" or "vascular cognitive impairment" or "Cerebral Autosomal Dominant Arteriopathy with Subcortical Infarcts and Leukoencephalopathy" or CADASIL or "Cerebral autosomal recessive arteriopathy with subcortical infarcts and leukoencephalopathy" or CARASIL or "mild cognitive impairment" or "vascular parkinsonism" or "neurocognitive disorder*" or "cerebral amyloid angiopathy" or "small vessel disease*" or "cerebrovascular disease*" or "cerebrovascular damage" or "population-based" or "community-based" or "community-dwelling" or "independent living") or AB (dementia or alzheimer* or "lewy bod*" or "frontotemporal lobar degenerat* OR frontotemporal dementia" or "progressive supranuclear palsy" or huntington* or "corticobasal degeneration" or "vascular cognitive impairment" or "Cerebral Autosomal Dominant Arteriopathy with Subcortical Infarcts and Leukoencephalopathy" or CADASIL or "Cerebral autosomal recessive arteriopathy with subcortical infarcts and leukoencephalopathy" or CARASIL or "mild cognitive impairment" or "vascular parkinsonism" or "neurocognitive disorder*" or "cerebral amyloid angiopathy" or "small vessel disease*" or "cerebrovascular disease*" or "cerebrovascular damage" or "population-based" or "community-based" or "community-dwelling" or "independent living") or (MH "Alzheimer's Disease") or (MH "Huntington's Disease") or (MH "Lewy Body Disease") or (MH "Frontotemporal Lobar Degeneration") or (MH "Corticobasal Degeneration") or (MH "Dementia") or (MH "Neurodegenerative Diseases") or (MH "Mild Cognitive Impairment") or (MH "Supranuclear Palsy, Progressive") or (MH "CADASIL") or (MH "Cerebral Amyloid Angiopathy") or (MH "Cerebral Small Vessel Diseases") or (MH "Community Living")

AND

TI ("white matter hyperintensit*" or "white matter disease*" or "white matter lesion*" or leukoaraiosis* or lacune* or "lacunar infarct*" or "small subcortical infarct*" or "perivascular space*" or "virchow robin space*" or microbleed* or microinfarct* or Leukoencephalopath* or "cerebrovascular damage" or haemorrhag* or hemorrhag* or "cerebrovascular accident" or stroke) or AB ("white matter hyperintensit*" or "white matter disease*" or "white matter lesion*" or leukoaraiosis* or lacune* or "lacunar infarct*" or "small subcortical infarct*" or "perivascular space*" or "virchow robin space*" or microbleed* or microinfarct* or Leukoencephalopath* or "cerebrovascular damage" or haemorrhag* or hemorrhag* or "cerebrovascular accident" or stroke) or (MH "Stroke, Lacunar") or (MH "Stroke")

AND

TI ("Artificial Intelligence" or "neural network*" or "deep learning" or "machine learning" or "computer reasoning" or "machine intelligence" or "ridge regression" or "least absolute shrinkage and selection operator" or LASSO or "elastic net" or classifier or "support vector machine*" or "random forest*" or "reinforcement learning" or boost* or "nearest-neighbo*" or markov or clustering or "cluster analysis" or convolution or classification or "decision support system*" or automl or ensemble or unsupervi* or supervi* or "semi-supervi*" or "weakly supervi*" or "dimensionality reduction") or AB ("Artificial Intelligence" or "neural network*" or "deep learning" or "machine learning" or "computer reasoning" or "machine intelligence" or "ridge regression" or "least absolute shrinkage and selection operator" or LASSO or "elastic net" or classifier or "support vector machine*" or "random forest*" or "reinforcement learning" or boost* or "nearest-neighbo*" or markov or clustering or "cluster analysis" or convolution or classification or "decision support system*" or automl or ensemble or unsupervi* or supervi* or "semi-supervi*" or "weakly supervi*" or "dimensionality reduction") or (MH "Artificial Intelligence") or (MH "Neural Networks (Computer)") or (MH "Deep Learning") or (MH "Machine Learning") or (MH "Support Vector Machine") or (MH "Random Forest")

AND

TI (Neuroimag* or "Magnetic Resonance Imag*" or "Positron Emission Tomography" or "structural imaging" or "fluid attenuated inversion recovery" or "functional imaging" or "arterial spin labelling" or "dynamic imaging" or "DCE-MRI" or angiograph* or "computed tomography" or "CT perfusion" or "diffusion weighted imaging" or "proton density") or AB (Neuroimag* or "Magnetic Resonance Imag*" or "Positron Emission Tomography" or "structural imaging" or "fluid attenuated inversion recovery" or "functional imaging" or "arterial spin labelling" or "dynamic imaging" or "DCE-MRI" or angiograph* or "computed tomography" or "CT perfusion" or "diffusion weighted imaging" or "proton density") or (MH "Positron-Emission Tomography") or (MH "Magnetic Resonance Imaging") or (MH "Angiography") or (MH "Computed Tomography Angiography")

**PsycInfo (via Ebscohost)**

TI (dementia or alzheimer* or "lewy bod*" or "frontotemporal lobar degenerat* OR frontotemporal dementia" or "progressive supranuclear palsy" or huntington* or "corticobasal degeneration" or "vascular cognitive impairment" or "Cerebral Autosomal Dominant Arteriopathy with Subcortical Infarcts and Leukoencephalopathy" or CADASIL or "Cerebral autosomal recessive arteriopathy with subcortical infarcts and leukoencephalopathy" or CARASIL or "mild cognitive impairment" or "vascular parkinsonism" or "neurocognitive disorder*" or "cerebral amyloid angiopathy" or "small vessel disease*" or "cerebrovascular disease*" or "cerebrovascular damage" or "population-based" or "community-based" or "community-dwelling" or "independent living") or AB (dementia or alzheimer* or "lewy bod*" or "frontotemporal lobar degenerat* OR frontotemporal dementia" or "progressive supranuclear palsy" or huntington* or "corticobasal degeneration" or "vascular cognitive impairment" or "Cerebral Autosomal Dominant Arteriopathy with Subcortical Infarcts and Leukoencephalopathy" or CADASIL or "Cerebral autosomal recessive arteriopathy with subcortical infarcts and leukoencephalopathy" or CARASIL or "mild cognitive impairment" or "vascular parkinsonism" or "neurocognitive disorder*" or "cerebral amyloid angiopathy" or "small vessel disease*" or "cerebrovascular disease*" or "cerebrovascular damage" or "population-based" or "community-based" or "community-dwelling" or "independent living") or

DE "Alzheimer's Disease" or DE "Huntingtons Disease" or DE "Dementia with Lewy Bodies" or DE "Frontotemporal Lobar Degeneration" or DE "Corticobasal Degeneration" or DE "Dementia" or DE "Neurocognitive Disorders" or DE "Neurodegenerative Diseases" or DE "Mild Cognitive Impairment" or DE "Progressive Supranuclear Palsy"

AND

TI ("white matter hyperintensit*" or "white matter disease*" or "white matter lesion*" or leukoaraiosis* or lacune* or "lacunar infarct*" or "small subcortical infarct*" or "perivascular space*" or "virchow robin space*" or microbleed* or microinfarct* or Leukoencephalopath* or "cerebrovascular damage" or haemorrhag* or hemorrhag* or "cerebrovascular accident" or stroke) or AB ("white matter hyperintensit*" or "white matter disease*" or "white matter lesion*" or leukoaraiosis* or lacune* or "lacunar infarct*" or "small subcortical infarct*" or "perivascular space*" or "virchow robin space*" or microbleed* or microinfarct* or Leukoencephalopath* or "cerebrovascular damage" or haemorrhag* or hemorrhag* or "cerebrovascular accident" or stroke) or DE "Leukoaraiosis" or DE "Leukoencephalopathy" or DE "Cerebrovascular Accidents"

AND

TI ("Artificial Intelligence" or "neural network*" or "deep learning" or "machine learning" or "computer reasoning" or "machine intelligence" or "ridge regression" or "least absolute shrinkage and selection operator" or LASSO or "elastic net" or classifier or "support vector machine*" or "random forest*" or "reinforcement learning" or boost* or "nearest-neighbo*" or markov or clustering or "cluster analysis" or convolution or classification or "decision support system*" or automl or ensemble or unsupervi* or supervi* or "semi-supervi*" or "weakly supervi*" or "dimensionality reduction") OR AB ("Artificial Intelligence" or "neural network*" or "deep learning" or "machine learning" or "computer reasoning" or "machine intelligence" or "ridge regression" or "least absolute shrinkage and selection operator" or LASSO or "elastic net" or classifier or "support vector machine*" or "random forest*" or "reinforcement learning" or boost* or "nearest-neighbo*" or markov or clustering or "cluster analysis" or convolution or classification or "decision support system*" or automl or ensemble or unsupervi* or supervi* or "semi-supervi*" or "weakly supervi*" or "dimensionality reduction") or DE "Artificial Intelligence" or DE "Neural Networks" or DE "Deep Neural Networks" or DE "Machine Learning" or DE "Markov Chains" or DE "Cluster Analysis" or DE "Decision Support Systems"

AND

TI (Neuroimag* or "Magnetic Resonance Imag*" or "Positron Emission Tomography" or "structural imaging" or "fluid attenuated inversion recovery" or "functional imaging" or "arterial spin labelling" or "dynamic imaging" or "DCE-MRI" or angiograph* or "computed tomography" or "CT perfusion" or "diffusion weighted imaging" or "proton density") or AB (Neuroimag* or "Magnetic Resonance Imag*" or "Positron Emission Tomography" or "structural imaging" or "fluid attenuated inversion recovery" or "functional imaging" or "arterial spin labelling" or "dynamic imaging" or "DCE-MRI" or angiograph* or "computed tomography" or "CT perfusion" or "diffusion weighted imaging" or "proton density") or DE "Positron Emission Tomography" or DE "Functional Magnetic Resonance Imaging" or DE "Neuroimaging" or DE "Angiography"

**BNI (via ProQuest)**

TIAB (dementia or alzheimer* or "lewy bod*" or "frontotemporal lobar degenerat* OR frontotemporal dementia" or "progressive supranuclear palsy" or huntington* or "corticobasal degeneration" or "vascular cognitive impairment" or "Cerebral Autosomal Dominant Arteriopathy with Subcortical Infarcts and Leukoencephalopathy" or CADASIL or "Cerebral autosomal recessive arteriopathy with subcortical infarcts and leukoencephalopathy" or CARASIL or "mild cognitive impairment" or "vascular parkinsonism" or "neurocognitive disorder*" or "cerebral amyloid angiopathy" or "small vessel disease*" or "cerebrovascular disease*" or "cerebrovascular damage" or "population-based" or "community-based" or "community-dwelling" or "independent living") or MAINSUBJECT.EXACT("Alzheimer's Disease") OR MAINSUBJECT.EXACT("Huntingtons Disease") OR MAINSUBJECT.EXACT("Dementia")

AND

TIAB ("white matter hyperintensit*" or "white matter disease*" or "white matter lesion*" or leukoaraiosis* or lacune* or "lacunar infarct*" or "small subcortical infarct*" or "perivascular space*" or "virchow robin space*" or microbleed* or microinfarct* or Leukoencephalopath* or "cerebrovascular damage" or haemorrhag* or hemorrhag* or "cerebrovascular accident" or stroke) or MAINSUBJECT.EXACT("Stroke")

AND

TIAB ("Artificial Intelligence" or "neural network*" or "deep learning" or "machine learning" or "computer reasoning" or "machine intelligence" or "ridge regression" or "least absolute shrinkage and selection operator" or LASSO or "elastic net" or classifier or "support vector machine*" or "random forest*" or "reinforcement learning" or boost* or "nearest-neighbo*" or markov or clustering or "cluster analysis" or convolution or classification or "decision support system*" or automl or ensemble or unsupervi* or supervi* or "semi-supervi*" or "weakly supervi*" or "dimensionality reduction") or MAINSUBJECT.EXACT("Artificial Intelligence") or MAINSUBJECT.EXACT("Neural networks") or MAINSUBJECT.EXACT("Deep learning") or MAINSUBJECT.EXACT("Machine learning") or MAINSUBJECT.EXACT("Support vector machines") or MAINSUBJECT.EXACT("Markov analysis") or MAINSUBJECT.EXACT("Decision support systems")

AND

TIAB (Neuroimag* or "Magnetic Resonance Imag*" or "Positron Emission Tomography" or "structural imaging" or "fluid attenuated inversion recovery" or "functional imaging" or "arterial spin labelling" or "dynamic imaging" or "DCE-MRI" or angiograph* or "computed tomography" or "CT perfusion" or "diffusion weighted imaging" or "proton density") or MAINSUBJECT.EXACT("Neuroimaging") or MAINSUBJECT.EXACT("Magnetic resonance imaging")

**Scopus**

Title-Abs-Key (dementia or alzheimer* or "lewy bod*" or "frontotemporal lobar degenerat* OR frontotemporal dementia" or "progressive supranuclear palsy" or huntington* or "corticobasal degeneration" or "vascular cognitive impairment" or "Cerebral Autosomal Dominant Arteriopathy with Subcortical Infarcts and Leukoencephalopathy" or CADASIL or "Cerebral autosomal recessive arteriopathy with subcortical infarcts and leukoencephalopathy" or CARASIL or "mild cognitive impairment" or "vascular parkinsonism" or "neurocognitive disorder*" or "cerebral amyloid angiopathy" or "small vessel disease*" or "cerebrovascular disease*" or "cerebrovascular damage" or "population-based" or "community-based" or "community-dwelling" or "independent living")

AND

Title-Abs-Key ("white matter hyperintensit*" or "white matter disease*" or "white matter lesion*" or leukoaraiosis* or lacune* or "lacunar infarct*" or "small subcortical infarct*" or "perivascular space*" or "virchow robin space*" or microbleed* or microinfarct* or Leukoencephalopath* or "cerebrovascular damage" or haemorrhag* or hemorrhag* or "cerebrovascular accident" or stroke)

AND

Title-Abs-Key ("Artificial Intelligence" or "neural network*" or "deep learning" or "machine learning" or "computer reasoning" or "machine intelligence" or "ridge regression" or "least absolute shrinkage and selection operator" or LASSO or "elastic net" or classifier or "support vector machine*" or "random forest*" or "reinforcement learning" or boost* or "nearest-neighbo*" or markov or clustering or "cluster analysis" or convolution or classification or "decision support system*" or automl or ensemble or unsupervi* or supervi* or "semi-supervi*" or "weakly supervi*" or "dimensionality reduction")

AND

Title-Abs-Key (Neuroimag* or "Magnetic Resonance Imag*" or "Positron Emission Tomography" or "structural imaging" or "fluid attenuated inversion recovery" or "functional imaging" or "arterial spin labelling" or "dynamic imaging" or "DCE-MRI" or angiograph* or "computed tomography" or "CT perfusion" or "diffusion weighted imaging" or "proton density")

**Web of Science (Core Collection)**

TS=(dementia or alzheimer* or "lewy bod*" or "frontotemporal lobar degenerat* OR frontotemporal dementia" or "progressive supranuclear palsy" or huntington* or "corticobasal degeneration" or "vascular cognitive impairment" or "Cerebral Autosomal Dominant Arteriopathy with Subcortical Infarcts and Leukoencephalopathy" or CADASIL or "Cerebral autosomal recessive arteriopathy with subcortical infarcts and leukoencephalopathy" or CARASIL or "mild cognitive impairment" or "vascular parkinsonism" or "neurocognitive disorder*" or "cerebral amyloid angiopathy" or "small vessel disease*" or "cerebrovascular disease*" or "cerebrovascular damage" or "population-based" or "community-based" or "community-dwelling" or "independent living")

AND

TS=("white matter hyperintensit*" or "white matter disease*" or "white matter lesion*" or leukoaraiosis* or lacune* or "lacunar infarct*" or "small subcortical infarct*" or "perivascular space*" or "virchow robin space*" or microbleed* or microinfarct* or Leukoencephalopath* or "cerebrovascular damage" or haemorrhag* or hemorrhag* or "cerebrovascular accident" or stroke)

AND

TS=("Artificial Intelligence" or "neural network*" or "deep learning" or "machine learning" or "computer reasoning" or "machine intelligence" or "ridge regression" or "least absolute shrinkage and selection operator" or LASSO or "elastic net" or classifier or "support vector machine*" or "random forest*" or "reinforcement learning" or boost* or "nearest-neighbo*" or markov or clustering or "cluster analysis" or convolution or classification or "decision support system*" or automl or ensemble or unsupervi* or supervi* or "semi-supervi*" or "weakly supervi*" or "dimensionality reduction")

AND

TS=(Neuroimag* or "Magnetic Resonance Imag*" or "Positron Emission Tomography" or "structural imaging" or "fluid attenuated inversion recovery" or "functional imaging" or "arterial spin labelling" or "dynamic imaging" or "DCE-MRI" or angiograph* or "computed tomography" or "CT perfusion" or "diffusion weighted imaging" or "proton density")

### Supplementary Table 1: Description of the studies

#### Diagnostic studies

| Reference | Dataset | N (% women) | | Age (SD) | | Ethnicity |
|---|---|---|---|---|---|---|
| | | Total | Per diagnosis | Total | Per diagnosis | |
| Ciulli et al., 2016 [4] | Local | 40 (34.0%) | MCI: 40 (34.0%) | 75.3 (6.8) | MCI: 75.3 (6.8) | - |
| Chan et al., 2023 [5] | CCNA# | 200 (52.0%) | HC: 48 (83.3%) \| MCI: 65 (44.6%) \| scVMCI: 44 (38.6%) \| AD: 21 (28.5%) \| Mixed: 22 (54.5%) | 74.7 | HC: 69.7 \| MCI: 71.9 \| scVMCI: 45.6 \| AD: 75.7 \| Mixed: 78.8 | - |
| Chen et al., 2019 [6] | Local | 112 (48.2%) | HC: 38 (47.3%) \| WMH-NC: 36 (58.3%) \| WMH-MCI: 38 (39.4%) | - | HC: 61.34 (1.16) \| WMH-NC: 65.03 (1.21) \| WMH-MCI: 64.84 (1.27) | - |
| Chen et al., 2023 [7] | Local | 178 (49.4%) | HC: 59 (37.2%) \| WMH-NC: 51 (50.9%) \| WMH-MCI: 68 (58.8%) | 63.7 (1.2) | HC: 64.56 (7.43) \| WMH-NC: 67.22 (6.88) \| WMH-MCI: 66.87 (6.89) | |
| Diciotti et al., 2015 [8] | Local | 87 | HC: 143 \| VCD: 58 | - | HC: 71.9 (6.1) \| VCD (low): 77.6 (5.6) \| VCD (high): 74.0 (7.0) | - |
| Dyrba et al., 2013 [9] | EDSD# | 280 (53.9%) | HC: 143 (50.3%) \| AD: 137 (57.6%) | - | HC: 69.2 (5.9) \| AD: 72.5 (8.3) | - |

| | | | | | | |
|---|---|---|---|---|---|---|
| Chen et al., 2017 [10] | Local | 53 (54.7%) | HC: 26 (57.7%) \| AD: 27 (51.8%) | - | HC: 66.0 (8.1) \| AD: 66.5 (7.7) | - |
| **Haller et al., 2010 [11]** | Local | 104 (62.5%) | HC: 35 (74.3%) \| sMCI: 40 (67.5%) \| pMCI: 29 (44.4%) | - | HC: 63.7 (5.1) \| sMCI: 65.4 (5.4) \| pMCI: 63.2 (4.2) | - |
| Han et al., 2021 [12] | Local | 208 (38.0%) | HC: 84 \| MCI: 124 | 40-80 | - | - |
| Zhu et al., 2019 [13] | Local | 131 | - | - | HC: 63.29 (6.50) \| WMH-NCI: 65.72 (5.94) \| WMH-MCI: 65.17 (6.65) | - |
| Qin et al. 2023 [14] | Local | 213 (38.0 %) | Training:<br>SVD-CI: 83 (37.4%) \| SVD-NC: 53 (39.6%)<br><br>Validation:<br>SVD-CI: 45 (27.8%) \| SVD-NC: 32 (40.6%) | - | Training:<br>SVD-CI: 65.9 (7.8) \|<br>SVD-NC: 63.4 (7.9)<br><br>Validation:<br>SVD-CI: 66.0 (7.4) \|<br>SVD-NC: 64.0 (8.2) | - |
| Smith et al., 2016 [15] | Local | 142 (63.3%) | VaD: 13 (61.5%) \| AD: 129 (69.0%) | - | VaD: 80.5 (6.4) \| AD: 75.1 (7.6) | - |
| Stebbins et al., 2008 [16] | Local | 91 (49.5%) | Stroke-NC: 51 (48.0%) \| Stroke-CI: 40 (51.2%) | - | Stroke-NC: 63.1 (8.2) \| Stroke-CI: 67.4 (9.7) | - |
| Tu et al., 2021 [17] | Local | 77 (49.4%) | HC: 24 (50.0%) \| scMCI: 23 (47.8%) \| AD: 30 (50.0%) | 73.2 (7.4) | HC: 65.9 (7.3) \| scMCI: 74.0 (8.5) \| AD: 78.3 (6.4) | - |
| Wan et al., 2022 [18] | Local | 438 (60.7%) | AD: 438 (early onset: 257 (65.0%); late onset: 181 (55.2)) | 64.8 | AD (early onset): 57.8 (5.7) \| AD (late onset): 74.7 (5.3) | - |

| | | | | | | |
|---|---|---|---|---|---|---|
| **Wang et al., 2019b [19]** | NHIS-IH, ADNI 2# | 390 (58.2%) | HC: 107 (NHIS-IH: 36 (88.9%) \| ADNI-2: 71 (60.6%))<br><br>MCI: 122 (NHIS-IH: 62 (61.3%) \| ADNI-2: 60 (33.3%))<br><br>AD: 158 (NHIS-IH: 110 (67.3%) \| ADNI-2: 48 (41.7%)) | 74.8 (7.1) | HC: NHIS-IH: 72.3 (7.0) \| ADNI-2: 72.6 (5.7)<br><br>MCI: NHIS-IH: 71.4 (8.6) \| ADNI-2: 72.6 (6.6)<br><br>AD: 80.0 (6.6) \| ADNI-2: 74.96 (8.6) | - |
| Joo et al., 2022 [20] | Local | 800 (59.9%) | Training: 596 (58.7%) \| Test: 204 (63.2%) | 67.2 (9.2) | Training: 69.1 (10.4) \| Test: 69.4 (10.8) | - |
| Lai et al., 2015 [21] | Local | 44 | HC: 10 \| MCI: 25 \| SVD-dementia: 9 | - | - | - |
| Lee et al., 2022 [22] | Training: ADNI#<br>Test: CARPET | 803 (45.6%) | HC: 401 (50.9%) \| Stroke-NC: 78 (37.2%) \| AD: 292 (40.8%) \| Pre-stroke dementia: 13 (61.5%) \| Post-stroke dementia: 19 (31.6%) | 72.1 (9.9) | HC: 73.8 (6.0) \| Stroke-NC: 70.1 (10.5) \| AD: 74.9 (8.0) \| Pre-stroke dementia: 79.5 (4.8) \| Post-stroke dementia: 75.5 (6.7) | - |
| Li et al., 2017 [23] | Local | 50 (50.0%) | HC: 21 (47.6%) \| MCI: 16 (62.5%) \| Dementia: 13 (38.5%) | - | HC: 58.8 (8.8) \| MCI: 64.2 (11.0) \| Dementia: 65.0 (13.3) | - |
| Lin et al., 2014 [24] | Local | 60 (45.0%) | HC: 30 (56.7%) \| ICA stenosis: 30 (43.3%) | - | HC: 69.8 (5.8) ICA stenosis: 70.8 (8.3) | - |
| **Lindemer et al., 2018 [25]** | ADNI-1# | 236 (34.7%) | HC: 56 (62.0%) \| MCI: 119 (34.0%) \| AD: 61 (44.0%) | - | HC: 75.2 \| MCI: 75.2 \| AD: 76.0 | - |
| Ma et al., 2022 [26] | Local | 72 (47.2%) | HC: 36 (58.3%) \| scMCI: 36 (36.1%) | - | HC: 57.3 (6.6) \| scMCI: 57.9 (6.6) | - |

| | | | | | | |
|---|---|---|---|---|---|---|
| Kandiah et al., 2013 [27] | Local | 91 (28.6%) | PD-NC: 67 (29.9%) \| PD-MCI: 24 (25.0%) | 64.9 | PD-NC: 63.4 (7.5) \| PD-MCI: 69.0 (6.1) | Asian: 81 (89.0%) \| Other: 10 (11.0%) |
| Meng et al., 2016 [28] | CAD imaging study | 108 (38.9%) | VCD-NC: 55 (40.0%) \| VCD-CI: 53 (37.7%) | 74.8 (8.9) | VCD-NC: 72.7 (9.5) \| VCD-CI: 76.6 (8.6) | - |
| Appel et al., 2009 [29] | Local | 192 | HC: 40 (67.5%) \| naMCI: 53 (40.7%) \| aMCI: 65 (46.7%) \| AD: 34 (53.3%) | - | HC: 71.0 (5.6) \| naMCI: 74.2 (6.1) \| aMCI: 75.3 (6.6) \| AD: 76.6 (6.5) | - |
| Park et al., 2021 [30] | Algorithm validated on an external dataset, ADNI# | 243 (49.4%) | HC: 73 (50.7%) \| MCI: 115 (51.3%) \| AD: 55 (43.6%) | 73.0 (7.5) | HC: 75.0 (6.5) \| MCI: 71.0 (7.9) \| AD: 76.0 (7.1) | White: 218 (90%) \| Other: 25 (10%) |
| Oppedal et al., 2017 [31] | DemWest, ParkWest (NC) | 182 | HC: 36 \| Mild dementia: 73 \| AD: 57 \| LBD: 16 | 70-75 | - | - |
| Bordin et al., 2022 [32] | OASIS-3# | 251 (53.4%) | HC: 212 \| AD: 39 | 70.3 (8.6) | - | - |
| Cajanus et al., 2018 [33] | Various local, ADNI# | 1161 | FTD: 22 \| LBD: 14 \| AD: 8 \| SMC: 6 | - | 62.5 | - |
| Oppedal et al., 2015 [34] | DemWest, ParkWest (for NC) | 182 | HC: 36 \| Mild dementia: 73 \| AD: 57 \| LBD: 16 | 70-75 | - | - |
| Chen et al., 2020 [35] | Local | 81 (45.7%) | HC: 18 (61.1%) \| MCI: 25 (35.6%) \| AD: 10 (41.7%) | - | HC: 62.0 (6.5) \| MCI: 67.1 (9.5) \| AD: 64.9 (8.1) | - |

| | | | | | | |
|---|---|---|---|---|---|---|
| Suresh et al., 2018 [36] | ADNI# | 406 (51.0%) | HC: 269 (55.4%) \| AD: 137 (42.3%) | - | HC: 72.9 \| AD: 74.2 | - |
| **Provenzano et al., 2013 [37]** | ADNI# | 100 (34.0%) | HC: 21 (38.0%) \| MCI: 59 (31.0%) \| AD: 20 (40.0%) | 75.0 (16.7) | HC: 76.2 (6.0) \| MCI: 75.7 (7.9) \| AD: 73.0 (8.6) | - |
| Xie et al., 2015 [38] | Local | 128 (58.0%) | HC: 64 (56.0%) \| MCI: 64 (59.0%) | 47-75 | HC: 64.8 (7.6) \| MCI: 67.1 (9.3) | Asian: 128 (100%) |
| Li et al., 2020 [39] | HCP#, ADNI# | 1043 (50.0%) | HCP: 272 (54.0%) \| HC (ADNI): 268 (55.0%) \| AD (ADNI): 136 (42.0%) \| HC (Validation): 25 (48.0%) \| AD (Validation): 66 (46.0%) \| Early MCI: 180 \| Late MCI: 96 | 69.4 (9.2) | HCP: 62.7 (16.8) \| HC (ADNI): 72.9 (6.0) \| AD (ADNI): 74.2 (8.2) \| HC (Validation): 68.5 (6.1) \| AD (Validation): 68.7 (9.0) | - |
| Zhang et al., 2022 [40] | Local | 148 (53.3%) | HC: 74 (59.5%) \| SIVD-NC: 38 (39.5) \| SIVD-MCI: 36 (47.2) | - | HC: 60.8 (8.1) \| SIVD-NC: 62.0 (4.5) \| SIVD-MCI: 63.1 (5.5) | - |
| Zhang et al., 2022 [41] | Local | 313 (51.8%) | HC: 156 (50.6%) \| Dementia: 157 (52.9%) AD: 80 \| CSVD: 46 \| FTD: 2 \| PD: 2 \| Mixed: 27 | - | HC: 70.32 (7.97) \| Dementia: 70.46 (8.16) | - |
| Zhang et al., 2013 [42] | Local | 44 (43.2%) | HC: 19 (47.4%) \| bvFTD: 13 (23.1%) \| SD: 6 (33.3%) \| PNFA: 6 (83.3%) | - | HC: 63.1 (7.6) \| bvFTD: 59.5 (6.5) \| SD: 64.8 (5.8) \| PNFA: 64.3 (5.0) | - |

| | | | | | | |
|---|---|---|---|---|---|---|
| Zhao et al., 2019 [43] | ADNI# | 90 (53.3%) | HC: 45 (55.6%) \| AD: 45 (51.1%) | - | HC: 74.3 (8.4) \| AD: 72.6 (7.1) | - |
| Schouten et al., 2016 [44] | PRODEM | 250 (58.0%) | HC: 173 (57.0%) \| AD: 77 (Mild AD: 22 (56.0%) \| Moderate AD: 24 (63.0%)) | 66.9 (8.6) | HC: 66.1 (8.71) \| Mild AD: 70.3 (7.85) \| Moderate AD: 66.9 (9.06) | - |
| **Crystal et al., 2023 [45]** | ADNI#, CCNA#, ONDRI# | 6063 | Cross- sectional analysis: HC: 589 \| AD: 589 \| sMCI: 89 \| cMCI: 89<br>Longitudinal analysis: HC: 1632 \| AD: 2344 \| sMCI: 543 \| cMCI: 188 | - | - | - |
| Chen et al., 2024 [46] | ADNI# | 497 (48.5%) | HC: 149 (54.4%) \| MCI: 304 (46.4%) \| 44 (43.2%) | 73.4 (7.5) | HC: 73.5 (6.0) \| MCI: 71.5 (7.4) \| Dementia: 75.2 (9.2) | - |
| Shi et al., 2024 [47] | Local | 171 (40.9%) | HC: 54 (42.6%) \| CSVD-HC: 57 (38.6%) \| CSVD-CI: 60 (41.6%) | 68.8 (6.0) | HC: 67.4 (7.5) \| CSVD-HC: 68.7 (5.8) \| CSVD-CI: 70.2 (4.8) | - |
| Strain et al., 2023 [48] | ADNI# | 228 (48.2%) | HC: 98 (53.0%) \| AD: 130 (44.6%) | 72 (6.4) | HC: 71 (6.0) \| AD: 73 (6.8) | - |
| Zhu et al., 2024 [49] | Local | 463 (48.8%) | HC: 232 (7.9) \| MCI: 231 (8.1) | 61.8 (8.1) | HC: 60.6 (7.9) \| MCI: 63.0 (8.1) | - |
| De Francesco et al., 2023 [50] | ADNI#, FTLDNI#, NACC#, PDBP#, Newcastle | 506 (36.0%) | HC: 108 (51.8%) \| AD: 110 (40.9%) \| FTD: 135 (40.0%) \| DLB: 153 (18.4%) | 71.4 (8.4) | HC: 74.3 (10.3) \| AD: 73.6 (7.7) \| FTD: 63.9 (6.9) \| DLB: 73.9 (8.6) | - |

| | | | | | | |
|---|---|---|---|---|---|---|
| Feng et al., 2024 [51] | Hospitals | Training: 79 \| Testing: 29 | Training set and internal testing set: WMH with CI: 42 (81.0%) \| WMH without CI: 37 (68.0%)<br><br>External testing set: WMH with CI: 15 (33.0%) \| WMH without CI: 14 (75.0%) | Training set and internal testing set: 141<br><br>External testing set: 70 | Training set and internal testing set: WMH with CI: 74 (49-84) \| WMH without CI: 67 (47-80)<br><br>External testing set: WMH with CI: 70 (53-79) \| WMH without CI: 69 (54-79) | - |

-: not specified/available; #: dataset publicly available; Bold: both diagnosis and prognosis

**Datasets:**
Canadian Consortium on Neurodegeneration in Aging (CCNA): https://ccna.loris.ca/
European DTI Study on Dementia (EDSD): https://www.gaaindata.org/partner/EDSD
National Health Insurance Service Ilsan Hospital (NHIS-IH)
ADNI (Alzheimer's Disease Neuroimaging Initiative): https://adni.loni.usc.edu/
ADNI-2: https://adni.loni.usc.edu/about/adni-go/
Cerebral Atherosclerosis Research with Positron Emission Tomography (CARPET)
Carotid artery disease (CAD) imaging study
OASIS-3 (Open Access Series of Imaging Studies): https://sites.wustl.edu/oasisbrains/
DemWest
ParkWest
HCP (Human Connectome Project) Lifespan-Aging cohort: https://www.humanconnectome.org/study/hcp-lifespan-aging/data-releases
PRODEM (Prospective registry on dementia)
ONDRI (Ontario Neurodegenerative Disease Research Initiative): https://braininstitute.ca/ondri
FTLDNI (Frontotemporal Lobar Degeneration Neuroimaging Initiative): http://adni.loni.usc.edu
NACC (National Alzheimer's Coordinating Center): https://naccdata.org/
PDBP (NIH Parkinson's Disease Biomarkers Program): https://pdbp.ninds.nih.gov

Prognostic studies

| Reference | Dataset | N (% women) | | Age (SD) | | Ethnicity |
|---|---|---|---|---|---|---|
| | | Total | Per diagnosis | Total | Per diagnosis | |
| **Haller et al., 2010 [11]** | Local | 104 (62.5%) | HC: 35 (74.3%) \| sMCI: 40 (67.5%) \| pMCI: 29 (44.4%) | - | HC: 63.7 (5.1) \| sMCI: 65.4 (5.4) \| pMCI: 63.2 (4.2) | - |
| Jokinen et al., 2020 [52] | LADIS | 560 (54.1%) | - | 73.5 (5.1) | - | - |
| Rabin et al., 2020 [53] | HABS# | 250 (58.0%) | HC: 227 (57.3%) \| MCI: 23 (65.2%) | 73.6 (6.0) | HC: 73.3 (5.9) \| MCI: 76.2 (6.1) | - |
| Stephan et al., 2015 [54] | The Three City Study# | 1721 (60.7%) | HC: 1602 (60.6%) \| Dementia: 119 (61.3%) | 73.5 (4.1) | HC: 72.2 (4.1) \| Dementia: 74.8 (4.0) | - |
| Tang et al., 2021 [55] | ADNI-GO#, ADNI-2# | 162 (40.7%) | pMCI: 68 \| sMCI: 94 | 72.0 | Training: 71.7 (7.3) \| Testing: 72.1 (6.1) | - |
| Tozer et al., 2018 [56] | SCANS#, GENIE (for HC) | 175 (35.4%) | HC: 54 (35.2%) \| SVD: 121 (35.54%) | 70.2 (9.6) | HC: 70.4 (9.2) \| SVD: 70.0 (9.8) | - |
| Twait et al., 2023 [57] | AGES-Reykjavik# | 4793 (59.0%) | HC: 3901 \| Dementia: 892 | 76.0 (6.0) | - | - |
| Verdelho et al., 2012 [58] | LADIS | 639 (55.0%) | HC: 351 \| MCI: 61 \| VCD: 86 \| VaD: 54 \| AD: 34 \| FTD: 2 | 74.1 (5.0) | - | - |
| Wang et al., 2019 [59] | Rotterdam study# | 5496 (54.9%) | HC: 5337 (54.8%) \| Dementia: 159 (57.9%) | 66.3 (10.6) | HC: 65.7 (10.3) \| Dementia: 77.3 (7.2) | - |

| | | | | | | |
|---|---|---|---|---|---|---|
| **Wang et al., 2019 [19]** | NHIS-IH, ADNI 2# | 390 (58.2%) | HC: 107 (NHIS-IH: 36 (88.9%) \| ADNI-2: 71 (60.6%))<br><br>MCI: 122 (NHIS-IH: 62 (61.3%) \| ADNI-2: 60 (33.3%))<br><br>AD: 158 (NHIS-IH: 110 (67.3%) \| ADNI-2: 48 (41.7%)) | 74.8 (7.1) | HC: NHIS-IH: 72.3 (7.0) \| ADNI-2: 72.6 (5.7)<br><br>MCI: NHIS-IH: 71.4 (8.6) \| ADNI-2: 72.6 (6.6)<br><br>AD: 79.95 (6.61) \| ADNI-2: 74.96 (8.59) | - |
| Lambert et al., 2018 [60] | SCANS | 119 (36.1%) | SVD-HC: 97 (39.2%) \| SVD-dementia: 22 (22.7%) | - | SVD-HC: 69.5 (9.5) \| SVD-dementia: 73.2 (9.4) | - |
| Liang et al., 2023 [61] | ADNI# | 225 (50.7%) | HC: 91 (52.7%) \| sMCI: 90 (51.1%) \| pMCI: 44 (45.5%) | - | HC: 74.0 (7.3) \| sMCI: 74.4 (7.1) \| pMCI: 75.2 (8.2) | - |
| **Lindemer et al., 2018 [25]** | ADNI-1# | 236 (34.7%) | HC: 56 (62%) \| MCI: 119 (34%) \| AD: 61 (44%) | - | HC: 75.2 \| MCI: 75.2 \| AD: 76.0 | - |
| Binzer et al., 2022 [62] | Local | 414 | HC: 332 \| Stroke: 82 | - | - | - |
| Mortamais et al., 2013 [63] | ESPRIT Project (local) | 426 (55.0%) | HC (at follow-up): 315 \| MCI (at follow-up): 100 \| dementia (at follow-up): 11 | 71.1 (4.0) | - | - |
| Aam et al., 2020 [64] | Nor-COAST | 617 (42.0%) | - | 72.0 (12.0) | - | - |
| Aamodt et al., 2021 [65] | Nor-COAST | 227 (43.6%) | Stroke-NC: 165 (69.7%) Stroke-CI: 62 (30.3%) | 71.7 (11.3) | Stroke-NC: 69.6 (10.8) \| Stroke-CI: 77.4 (10.4) | - |
| Altieri et al., 2004 [66] | Local | 191 (30.9%) | VaD: 41 (24.4%) \| Stroke: 150 (32.7%) | 71.3 (8.9) | VaD: 71 (6.9) \| Stroke: 63.2 (9.5) | - |

| | | | | | | |
|---|---|---|---|---|---|---|
| Peters et al., 2013 [67] | Local | 65 (62.0%) | HC: 20 (70.0%) \| sMCI: 22 (59.0%) \| pMCI: 18 (56.0%) | 71.8 (6.8) | HC: 72.0 (6.9) \| sMCI: 70.4 (7.1) \| pMCI: 72.9 (6.3) | - |
| **Provenzano et al., 2013 [37]** | ADNI# | 100 (34.0%) | HC: 21 (38.0%) \| MCI: 59 (31.0%) \| AD: 20 (40.0%) | 75.0 (16.7) | HC: 76.2 (6.0) \| MCI: 75.7 (7.9) \| AD: 73.0 (8.6) | - |
| West et al., 2019 [68] | ARIC# | 1881 (60.0%) | No high-grade cerebral abnormalities: 1028 (66.0%) \| At least 1 high-grade cerebral abnormality: 853 (53.0%) | 62.4 (4.5) | No high-grade cerebral abnormalities: 61.4 (4.4) \| At least 1 high-grade cerebral abnormality: 63.5 (4.4) | Black: 934 (49.7%) \| White: 947 (50.3%) |
| Williams et al., 2019 [69] | SCANS | 99 (34.0%) | SVD-dementia: 18 (22.2%) \| SVD-CN: 81 (35.8%) | 68.4 (10.0) | SVD-dementia: 70.6 (2.6) \| SVD-CN: 68.5 (1.1) | - |
| Yao et al., 2014 [70] | 3C-Dijon MRI Study | 1818 (61.0%) | HC: 1724 \| Dementia: 94 | 72.5 (4.1) | - | - |
| Rosano et al., 2007 [71] | Cardiovascular Health Study cohort# | 155 (38.7%) | HC: 116 (55.2%) \| AD: 39 (71.8%) | 77.4 (3.4) | HC: 76.9 (3.4) \| AD: 78.2 (3.4) | White: 127 (81.9%) \| Other: 28 (17.1%) |
| Rosano et al., 2016 [72] | Cardiovascular Health Study cohort# | 5338 (57.6%) | HC: 4130 (59.2%) \| Subclinical MCI: 579 (56.5%) \| MCI: 629 (53.7%) | 75.1 (5.5) | HC: 74.3 (4.9) \| Subclinical MCI: 76.1 (5.9) \| MCI: 78.4 (6.8) | White: 4925 (83.6%) \| Other: 413 (16.4%) |

| | | | | | | |
|---|---|---|---|---|---|---|
| **Crystal et al., 2023 [45]** | ADNI#, CCNA#, ONDRI# | 6063 | Cross- sectional analysis: HC: 589 \| AD: 589 \| sMCI: 89 \| cMCI: 89<br>Longitudinal analysis: HC: 1632 \| AD: 2344 \| sMCI: 543 \| cMCI: 188 | - | - | - |
| Chen et al., 2024 [73] | UKBiobank#, SCANS, RUN DMC, PRESERVE, NETWORKS | 6620 | Training: HC: 5998 (46.7%)<br>Testing: HC: 518 (42.0%) \| Dementia: 104 | - | - | - |
| Chen et al., 2024 [74] | ADNI# | 357 (40.1%) | MCI-HC: 181 (41.4%) \| MCI-converter:176 (38.6%) | 73.7 (7.5) | MCI-HC: 72.4 (8.0) \| MCI-converter: 74.9 (6.9) | - |
| Keller et al., 2024 [75] | AGES-Reykjavik# | 3056 (61.6%) | HC: 2799 \| MCI: 257 | 75.6 (5.2) | - | - |
| Lee et al., 2024 [76] | Local, ADNI# | 196 (56.6%) | sMCI: 149 (57%) \| pMCI: 47 (55.0%) | 72.3 (6.9) | sMCI: 71.7 (7.2) \| pMCI: 72.9 (6.6) | - |
| Marzi et al., 2023 [77] | VMCI Tuscany study | 64 (46.9%) | sMCI: 46 (47.8%) \| pMCI: 18 (44.4%) | 75.2 (6.7) | sMCI: 74.0 (6.7) \| pMCI: 76.3 (6.7) | - |
| Li et al., 2023 [78] | RUN DMC, SCANS, HARMONISATION | 750 (46.3%) | No dementia after 3 years: 702 \| Dementia after 3 years: 48 | 67.2 (9.1) | - | - |

-: not specified/available; #: dataset publicly available; Bold: both diagnosis and prognosis

**Datasets:**

LADIS (Leukoaraiosis And DISability)

HABS (Harvard Aging Brain Study): https://habs.mgh.harvard.edu/

The three-city study: https://neurodegenerationresearch.eu/jpnd-global-cohort-portal/how-to-use-the-portal-page/

ADNI-GO, ADNI-2 (Alzheimer's Disease Neuroimaging Initiative): https://adni.loni.usc.edu/about/adni-go/

ADNI (Alzheimer's Disease Neuroimaging Initiative): https://adni.loni.usc.edu/

ADNI-1 (Alzheimer's Disease Neuroimaging Initiative): https://adni.loni.usc.edu/about/adni1/

SCANS (St George's Cognition and Neuroimaging in Stroke): https://cheba.unsw.edu.au/consortia/strokog/studies/st-georges-cognition-and-neuroimaging-in-stroke-scans *(does not seem to be publicly available, but contact information)*

GENIE (St George's Neuropsychology and Imaging in the Elderly study)

AGES-Reykjavik (Age, Gene/Environment Susceptibility): https://www.maelstrom-research.org/study/ages

Rotterdam study: https://www.maelstrom-research.org/study/rs

National Health Insurance Service Ilsan Hospital (NHIS-IH)

ESPRIT project

Nor-COAST (Norwegian Cognitive Impairment After Stroke) study

ARIC (Subset of the original Atherosclerosis Risk in Communities) study: https://biolincc.nhlbi.nih.gov/studies/aric/

3C-Dijon MRI study

Cardiovascular health study: https://biolincc.nhlbi.nih.gov/studies/chs/

Canadian Consortium on Neurodegeneration in Aging (CCNA): https://ccna.loris.ca/

ONDRI (Ontario Neurodegenerative Disease Research Initiative): https://braininstitute.ca/ondri

UKBiobank: https://www.ukbiobank.ac.uk/

RUN DMC study (Radboud University Nijmegen Diffusion Tensor and Magnetic Resonance Cohort)

PRESERVE (Blood Pressure in Established Cerebral Small Vessel Disease)

### Supplementary Figure 1: Meta-analysis

#### Supplementary Figure 1A: Healthy controls vs. all-cause dementia

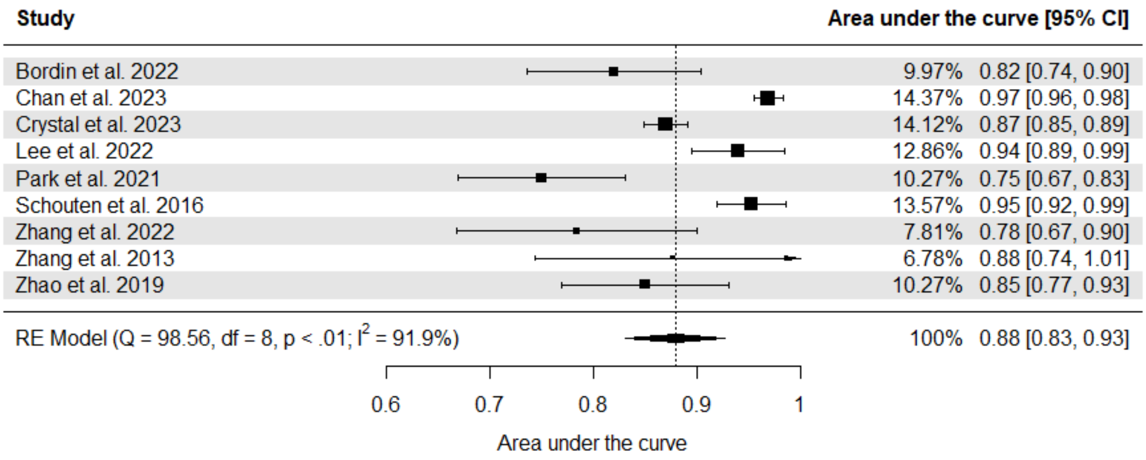

***Figure 1A.*** *Meta-analysis of studies classifying all-cause dementia vs healthy controls. Confidence intervals might exceed 1.00 because standard errors have been estimated due to missing data.*

#### Supplementary Figure 1B: Sensitivity analysis

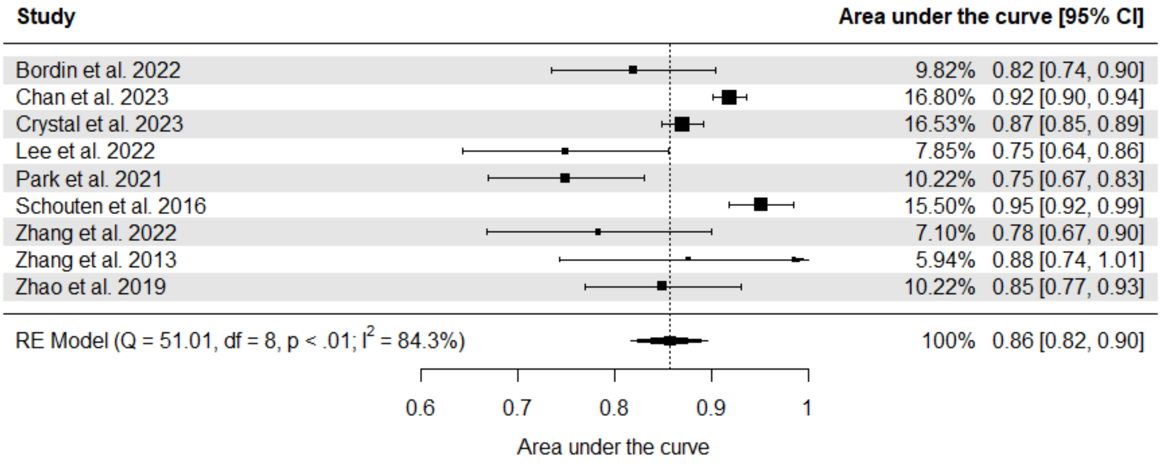

***Figure 1B.*** *Meta-analysis of studies classifying all-cause dementia vs healthy controls, including multiple classifiers from single papers. The results are similar whether these studies are included or excluded. Confidence intervals might exceed 1.00 because standard errors have been estimated due to missing data.*

#### Risk of bias assessment

Supplementary table 2A: Risk of bias assessment for each diagnostic study using the QUADAS-2 framework.

| Author, Year | Risk of Bias | | | | Applicability | | |
|---|---|---|---|---|---|---|---|
| | 1. Patient selection | 2. Index test | 3. Reference standard | 4. Flow and timing | 1. Patient selection | 2. Index test | 3. Reference standard |
| Ciulli et al., 2016 [4] | ☺ | ? | ☺ | ? | ☺ | ? | ☺ |
| Chan et al., 2023 [5] | ☺ | ? | ? | ☺ | ☺ | ? | ☺ |
| Chen et al., 2019 [6] | ☺ | ? | ☺ | ☺ | ☺ | ☺ | ☺ |
| Chen et al., 2023 [7] | ☺ | ? | ? | ? | ☺ | ☺ | ☺ |
| Diciotti et al., 2015 [8] | ☺ | ? | ? | ☺ | ☺ | ☺ | ☺ |
| Dyrba et al., 2013 [9] | ☹ | ☺ | ☺ | ☺ | ☺ | ☺ | ☺ |
| Chen et al., 2017 [10] | ☺ | ☺ | ☺ | ☺ | ☺ | ☺ | ☺ |
| Haller et al., 2010 [11] | ☹ | ☺ | ☺ | ☺ | ☺ | ☺ | ☺ |
| Han et al., 2021 [12] | ☹ | ? | ☹ | ☹ | ☹ | ☹ | ☹ |
| Zhu et al., 2019 [13] | ☹ | ☺ | ☺ | ☺ | ☺ | ☹ | ☺ |
| Qin et al., 2023 [14] | ☹ | ☺ | ☺ | ☺ | ☹ | ☺ | ☹ |
| Smith et al., 2016 [15] | ☺ | ☺ | ☺ | ☺ | ☺ | ☺ | ☺ |
| Stebbins et al., 2008 [16] | ☺ | ? | ? | ☺ | ☺ | ? | ? |
| Tu et al., 2021 [17] | ☺ | ☺ | ? | ☺ | ☺ | ☺ | ☺ |
| Wan et al., 2022 [18] | ☺ | ☺ | ☺ | ☺ | ☺ | ☺ | ☺ |
| Wang et al., 2019 [19] | ☺ | ☺ | ? | ☺ | ☺ | ☺ | ☺ |
| Joo et al., 2022 [20] | ☺ | ☺ | ☺ | ☺ | ☺ | ☺ | ☺ |
| Lai et al., 2015 [21] | ☹ | ☹ | ? | ☺ | ☺ | ☺ | ☺ |
| Lee at al., 2022 [22] | ☺ | ? | ☺ | ☺ | ☺ | ☺ | ☺ |
| Li et al., 2017 [23] | ☹ | ? | ☺ | ☺ | ☺ | ☺ | ☺ |

| | | | | | | | |
|---|---|---|---|---|---|---|---|
| Lin et al., 2014 [24] | ☹ | ☺ | ☹ | ☺ | ☺ | ☺ | ☹ |
| Lindemer et al., 2018 [25] | ? | ☺ | ☺ | ☹ | ☺ | ☺ | ☺ |
| Ma et al., 2022 [26] | ☹ | ☺ | ☺ | ? | ☺ | ☺ | ☺ |
| Kandiah et al., 2013 [27] | ☹ | ? | ☺ | ☺ | ☺ | ☺ | ☺ |
| Meng et al., 2017 [28] | ☺ | ? | ☺ | ☺ | ☺ | ? | ☺ |
| Appel et al., 2009 [29] | ? | ☹ | ☹ | ? | ☺ | ☺ | ☹ |
| Park et al., 2021 [30] | ☺ | ☺ | ☺ | ☺ | ☺ | ☺ | ☺ |
| Oppedal et al., 2017 [31] | ? | ☺ | ? | ☹ | ☺ | ☺ | ☺ |
| Bordin et al., 2022 [32] | ? | ? | ? | ☹ | ? | ☺ | ? |
| Cajanus et al., 2018 [33] | ☹ | ? | ? | ? | ? | ? | ? |
| Oppedal et al., 2015 [34] | ? | ☺ | ☺ | ☹ | ☺ | ☺ | ☺ |
| Chen et al., 2020 [35] | ? | ? | ☺ | ? | ☺ | ☺ | ☺ |
| Suresh et al., 2018 [36] | ☺ | ☹ | ? | ? | ☺ | ☺ | ☺ |
| Provenzano et al., 2013 [37] | ☺ | ? | ☺ | ☺ | ☺ | ☺ | ☺ |
| Li et al., 2020 [39] | ☹ | ☺ | ☺ | ☺ | ? | ☺ | ☺ |
| Zhang et al., 2022 [40] | ☹ | ☺ | ☺ | ☺ | ☺ | ☺ | ☺ |
| Zhang et al., 2022 [41] | ☺ | ☺ | ☺ | ☺ | ☺ | ☺ | ☺ |
| Zhang et al., 2013 [42] | ☹ | ☺ | ☺ | ☺ | ☹ | ☺ | ☺ |
| Zhao et al., 2019 [43] | ☹ | ☺ | ☺ | ☹ | ☺ | ☺ | ☺ |
| Schouten et al., 2016 [44] | ☹ | ☹ | ☺ | ☺ | ☺ | ☺ | ☺ |
| Xie et al., 2015 [38] | ☺ | ☺ | ☺ | ☺ | ☺ | ☺ | ☺ |
| Chen et al., 2024 [46] | ? | ☺ | ☺ | ? | ☺ | ☺ | ☺ |
| Crystal et al., 2023 [45] | ? | ? | ☹ | ☺ | ☺ | ☺ | ? |

| | | | | | | | |
|---|---|---|---|---|---|---|---|
| De Francesco et al., 2023 [50] | ☺ | ☺ | ☺ | ☺ | ☺ | ☺ | ☺ |
| Feng et al., 2024 [51] | ☺ | ☺ | ☺ | ☺ | ☺ | ☺ | ☺ |
| Shi et al., 2024 [47] | ☹ | ☺ | ☺ | ☺ | ☺ | ☺ | ☺ |
| Strain et al., 2023 [48] | ☹ | ☺ | ☹ | ☹ | ☺ | ☺ | ☺ |
| Zhu et al., 2024 [49] | ☹ | ☺ | ? | ? | ☺ | ☺ | ☺ |

Supplementary figure 2A: Summary of risk of bias assessment for diagnostic studies using the QUADAS-2 framework

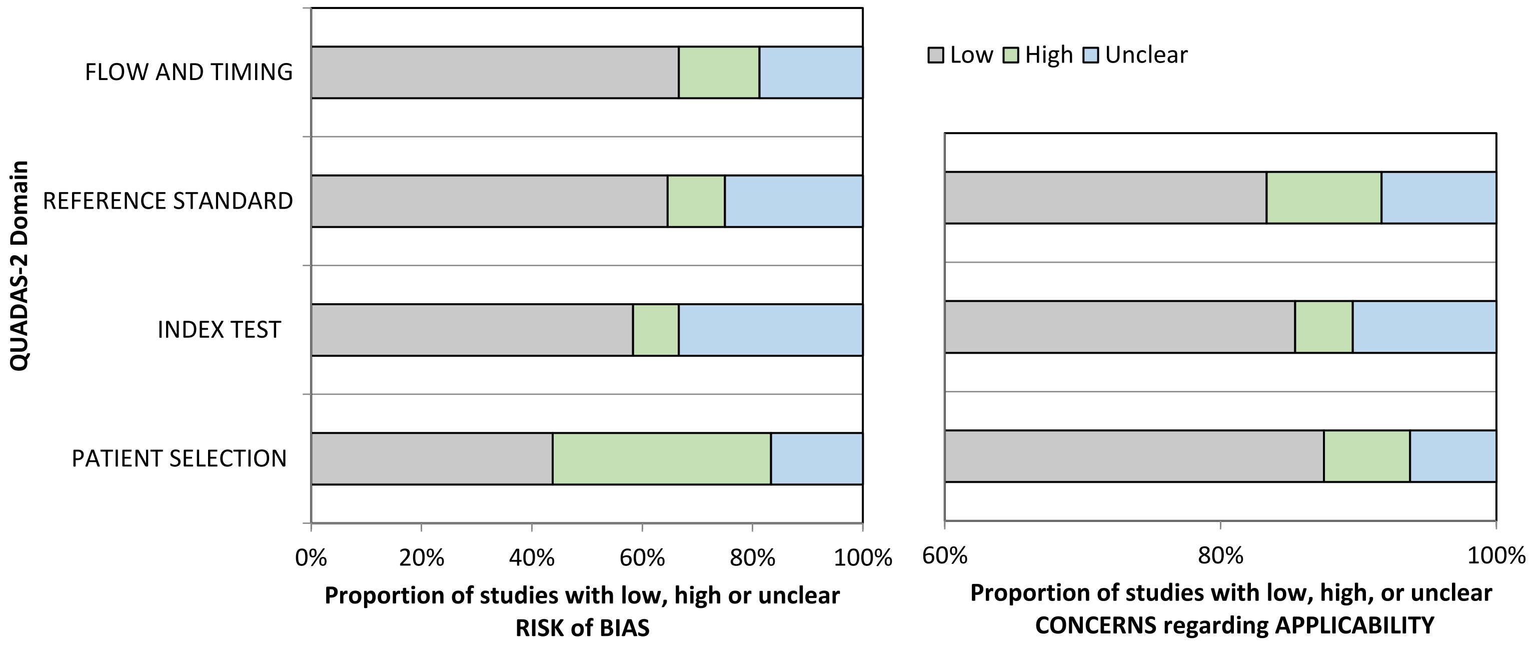

We provide summary statistics for each domain: flow and timing, reference standard, index test, and patient selection. The left plot displays the risk of bias and the right plot shows applicability concerns.

#### Supplementary table 2B: Risk of bias assessment for each prognostic study using the PROBAST framework studies

| Author, Year | Risk of Bias | | | | Applicability | | | Overall | |
|---|---|---|---|---|---|---|---|---|---|
| | 1. Participants | 2. Predictors | 3. Outcome | 4. Analysis | 1. Participants | 2. Predictors | 3. Outcome | **Risk of Bias** | **Applicability** |
| Jokinen et al., 2020 [52] | ☺ | ☺ | ☺ | ☺ | ☺ | ☺ | ☺ | ☺ | ☺ |
| Rabin et al., 2020 [53] | ☺ | ☺ | ☺ | ☺ | ☺ | ☺ | ☺ | ☺ | ☺ |
| Stephan et al., 2015 [54] | ? | ☺ | ☺ | ☹ | ☹ | ☺ | ☺ | ☹ | ☺ |
| Tang et al., 2021 [55] | ☹ | ☺ | ☺ | ? | ? | ☺ | ☺ | ? | ? |
| Tozer et al., 2018 [56] | ☺ | ☺ | ☺ | ☺ | ☺ | ☺ | ☺ | ☺ | ☺ |
| Twait et al., 2023 [57] | ☺ | ☺ | ☺ | ☺ | ☺ | ☺ | ☺ | ☺ | ☺ |
| Verdelho et al., 2012 [58] | ☺ | ☺ | ☺ | ☺ | ☺ | ☺ | ☺ | ☺ | ☺ |
| Wang et al., 2019 [59] | ☺ | ☺ | ☺ | ☺ | ☺ | ☺ | ☺ | ☺ | ☺ |
| Wang et al., 2019 [19] | ☺ | ☺ | ☺ | ☺ | ☺ | ☺ | ☺ | ☺ | ☺ |
| Lambert et al., 2018 [60] | ☺ | ☺ | ☺ | ☹ | ☺ | ☺ | ☺ | ☹ | ☺ |
| Lindemer et al., 2018 [25] | ? | ☺ | ☺ | ☹ | ☺ | ☺ | ☺ | ☹ | ☺ |
| Binzer et al., 2022 [62] | ☹ | ☺ | ? | ? | ☹ | ☺ | ? | ☹ | ☹ |
| Mortamais et al., 2013 [63] | ☺ | ☺ | ☺ | ☺ | ☺ | ☺ | ☺ | ☺ | ☺ |
| Aam et al., 2020 [64] | ☺ | ☺ | ☺ | ☹ | ☺ | ☺ | ☺ | ☹ | ☹ |
| Aamodt et al., 2021 [65] | ☺ | ☺ | ☺ | ? | ☺ | ☺ | ☺ | ? | ? |
| Altieri et al., 2004 [66] | ☺ | ☺ | ☺ | ☹ | ☺ | ☺ | ☺ | ☹ | ☹ |
| Peters et al., 2014 [67] | ☺ | ☺ | ☺ | ☺ | ☺ | ☺ | ☺ | ☹ | ☹ |
| Provenzano et al., 2013 [37] | ☺ | ☺ | ☺ | ☹ | ☺ | ☺ | ☺ | ? | ☹ |
| West et al., 2019 [68] | ☺ | ☺ | ☺ | ☺ | ☺ | ☺ | ☺ | ☺ | ☹ |
| Williams et al., 2019 [69] | ☺ | ☺ | ☺ | ☹ | ☺ | ☺ | ☺ | ☹ | ☹ |

| | | | | | | | | | |
|---|---|---|---|---|---|---|---|---|---|
| Yao et al., 2014 [70] | ☺ | ☹ | ☺ | ☹ | ☺ | ☺ | ☺ | ☹ | ☹ |
| Rosano et al., 2007 [71] | ☺ | ☺ | ☺ | ☹ | ☺ | ☺ | ☺ | ☹ | ☺ |
| Rosano et al., 2016 [72] | ☺ | ☺ | ☹ | ☹ | ☺ | ☺ | ☺ | ☹ | ☺ |
| Haller et al., 2010 [11] | ☺ | ☺ | ☺ | ☹ | ☺ | ☺ | ☺ | ☹ | ☺ |
| Chen et al., 2024 [73] | ? | ☺ | ? | ☺ | ☺ | ☺ | ☺ | ? | ☹ |
| Chen et al., 2024 [74] | ? | ☺ | ☺ | ? | ☺ | ☺ | ☺ | ? | ☹ |
| Crystal et al., 2023 [45] | ? | ☺ | ? | ? | ☺ | ☺ | ☺ | ? | ☹ |
| Keller et al., 2024 [75] | ☹ | ☺ | ☺ | ? | ☹ | ☺ | ☹ | ☹ | ☹ |
| Lee et al., 2024 [76] | ☺ | ? | ☹ | ☹ | ☺ | ☺ | ☺ | ☹ | ☺ |
| Marzi et al., 2023 [77] | ☺ | ☺ | ☺ | ☹ | ☺ | ☺ | ☺ | ☹ | ☺ |
| Li et al., 2023 [78] | ☺ | ☺ | ☺ | ☹ | ☺ | ☺ | ☺ | ☹ | ☺ |

Supplementary figure 2B: Summary of risk of bias assessment for prognostic studies using the PROBAST framework

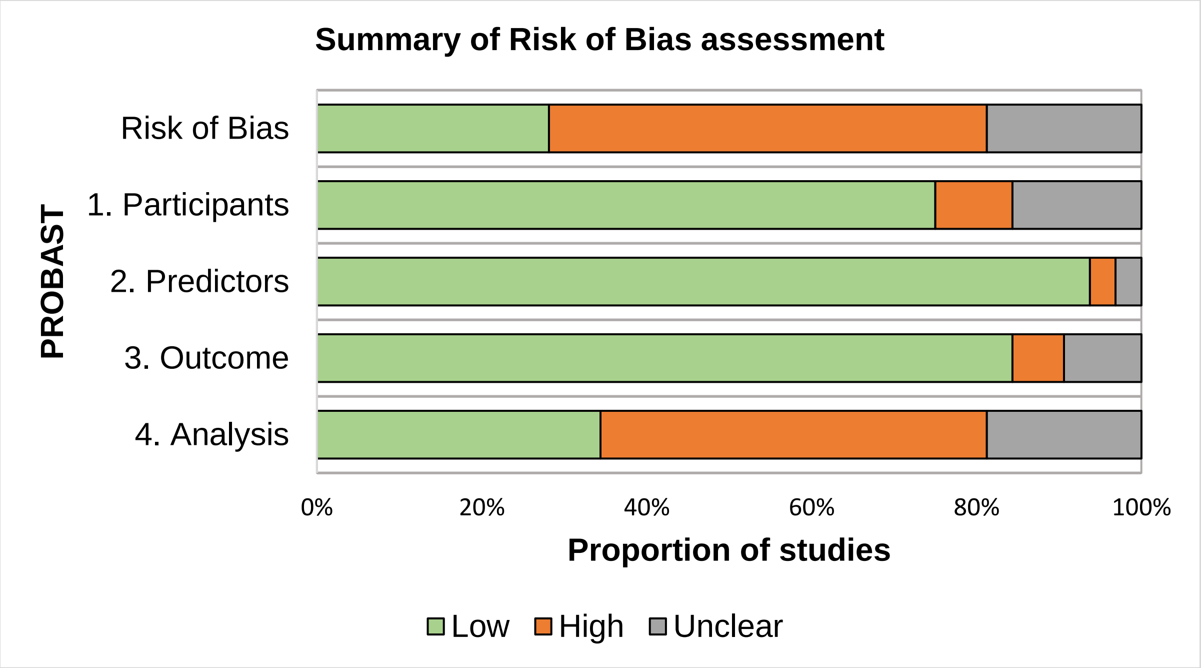

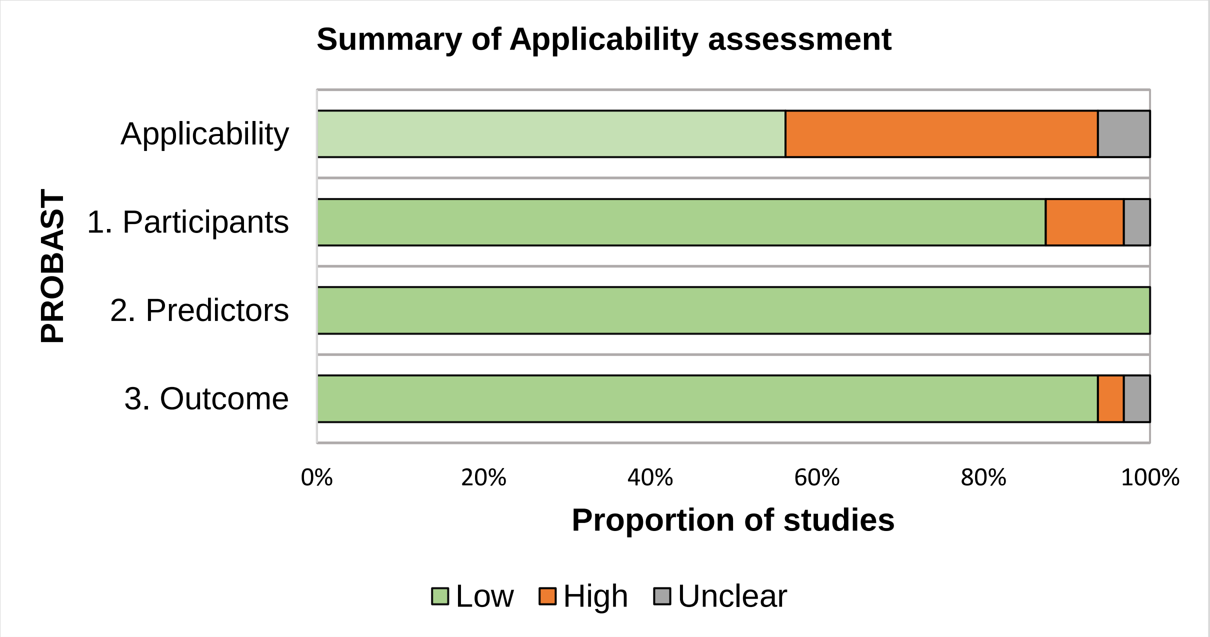

We provide summary statistics for each domain: participants, predictors, outcome, analysis, and applicability. The left plot displays the risk of bias and the right plot shows applicability concerns.

### References

[1] Duering M, Biessels GJ, Brodtmann A, Chen C, Cordonnier C, de Leeuw F-E, et al. Neuroimaging standards for research into small vessel disease-advances since 2013. The Lancet Neurology. 2023;22:602-18.
[2] Sarker IH. Machine Learning: Algorithms, Real-World Applications and Research Directions. SN Comput Sci. 2021;2:160.
[3] Jo T. Machine Learning Foundations. 2021.
[4] Ciulli S, Citi L, Salvadori E, Valenti R, Poggesi A, Inzitari D, et al. Prediction of Impaired Performance in Trail Making Test in MCI Patients With Small Vessel Disease Using DTI Data. IEEE J Biomed Health Inform. 2016;20:1026-33.
[5] Chan K, Fischer C, Maralani PJ, Black SE, Moody AR, Khademi A. Alzheimer’s and vascular disease classification using regional texture biomarkers in FLAIR MRI. NeuroImage: Clinical. 2023;38:103385.
[6] Chen H, Huang L, Yang D, Ye Q, Guo M, Qin R, et al. Nodal Global Efficiency in Front-Parietal Lobe Mediated Periventricular White Matter Hyperintensity (PWMH)-Related Cognitive Impairment. Front Aging Neurosci. 2019;11:347.
[7] Chen H, Xu J, Lv W, Hu Z, Ke Z, Qin R, et al. Altered static and dynamic functional network connectivity related to cognitive decline in individuals with white matter hyperintensities. Behav Brain Res. 2023;451:114506.
[8] Diciotti S, Ciulli S, Ginestroni A, Salvadori E, Poggesi A, Pantoni L, et al. Multimodal MRI classification in vascular mild cognitive impairment. 2015 37th Annual International Conference of the IEEE Engineering in Medicine and Biology Society (EMBC)2015. p. 4278-81.
[9] Dyrba M, Ewers M, Wegrzyn M, Kilimann I, Plant C, Oswald A, et al. Robust automated detection of microstructural white matter degeneration in Alzheimer's disease using machine learning classification of multicenter DTI data. PLoS One. 2013;8:e64925.
[10] Chen Y, Sha M, Zhao X, Ma J, Ni H, Gao W, et al. Automated detection of pathologic white matter alterations in Alzheimer's disease using combined diffusivity and kurtosis method. Psychiatry Res Neuroimaging. 2017;264:35-45.
[11] Haller S, Bartsch A, Nguyen D, Rodriguez C, Emch J, Gold G, et al. Cerebral microhemorrhage and iron deposition in mild cognitive impairment: susceptibility-weighted MR imaging assessment. Radiology. 2010;257:764-73.
[12] Han L, Liu L, Hao Y, Zhang L. Diagnosis and Treatment Effect of Convolutional Neural Network-Based Magnetic Resonance Image Features on Severe Stroke and Mental State. Contrast Media & Molecular Imaging. 2021;2021:8947789.
[13] Zhu W, Huang H, Yang S, Luo X, Zhu W, Xu S, et al. Dysfunctional Architecture Underlies White Matter Hyperintensities with and without Cognitive Impairment. Journal of Alzheimer’s Disease. 2019;71:461-76.
[14] Qin Q, Qu J, Yin Y, Liang Y, Wang Y, Xie B, et al. Unsupervised machine learning model to predict cognitive impairment in subcortical ischemic vascular disease. Alzheimers Dement. 2023;19:3327-38.
[15] Smith CD, Johnson ES, Van Eldik LJ, Jicha GA, Schmitt FA, Nelson PT, et al. Peripheral (deep) but not periventricular MRI white matter hyperintensities are increased in clinical vascular dementia compared to Alzheimer's disease. Brain and Behavior. 2016;6:e00438.
[16] Stebbins GT, Nyenhuis DL, Wang C, Cox JL, Freels S, Bangen K, et al. Gray matter atrophy in patients with ischemic stroke with cognitive impairment. Stroke. 2008;39:785-93.
[17] Tu MC, Huang SM, Hsu YH, Yang JJ, Lin CY, Kuo LW. Discriminating subcortical ischemic vascular disease and Alzheimer's disease by diffusion kurtosis imaging in segregated thalamic regions. Hum Brain Mapp. 2021;42:2018-31.
[18] Wan MD, Liu H, Liu XX, Zhang WW, Xiao XW, Zhang SZ, et al. Associations of multiple visual rating scales based on structural magnetic resonance imaging with disease severity and cerebrospinal fluid biomarkers in patients with Alzheimer's disease. Front Aging Neurosci. 2022;14:906519.

[19] Wang Y, Xu C, Park JH, Lee S, Stern Y, Yoo S, et al. Diagnosis and prognosis of Alzheimer's disease using brain morphometry and white matter connectomes. Neuroimage Clin. 2019;23:101859.
[20] Joo L, Shim WH, Suh CH, Lim SJ, Heo H, Kim WS, et al. Diagnostic performance of deep learning-based automatic white matter hyperintensity segmentation for classification of the Fazekas scale and differentiation of subcortical vascular dementia. PLoS One. 2022;17:e0274562.
[21] Lai Y, Xu L, Yao L, Wu X. Discriminative analysis of non-linear brain connectivity for leukoaraiosis with resting-state fMRI: SPIE; 2015.
[22] Lee R, Choi H, Park K-Y, Kim J-M, Seok JW. Prediction of post-stroke cognitive impairment using brain FDG PET: deep learning-based approach. European Journal of Nuclear Medicine and Molecular Imaging. 2022;49:1254-62.
[23] Li R, Lai Y, Zhang Y, Yao L, Wu X. Classification of Cognitive Level of Patients with Leukoaraiosis on the Basis of Linear and Non-Linear Functional Connectivity. Front Neurol. 2017;8.
[24] Lin C-J, Tu P-C, Chern C-M, Hsiao F-J, Chang F-C, Cheng H-L, et al. Connectivity Features for Identifying Cognitive Impairment in Presymptomatic Carotid Stenosis. PLOS ONE. 2014;9:e85441.
[25] Lindemer ER, Greve DN, Fischl B, Salat DH, Gomez-Isla T. White matter abnormalities and cognition in patients with conflicting diagnoses and CSF profiles. Neurology. 2018;90:e1461-e9.
[26] Ma J, Liu F, Wang Y, Ma L, Niu Y, Wang J, et al. Frequency-dependent white-matter functional network changes associated with cognitive deficits in subcortical vascular cognitive impairment. NeuroImage: Clinical. 2022;36:103245.
[27] Kandiah N, Mak E, Ng A, Huang S, Au WL, Sitoh YY, et al. Cerebral white matter hyperintensity in Parkinson's disease: A major risk factor for mild cognitive impairment. Parkinsonism & Related Disorders. 2013;19:680-3.
[28] Meng D, Hosseini AA, Simpson RJ, Shaikh Q, Tench CR, Dineen RA, et al. Lesion Topography and Microscopic White Matter Tract Damage Contribute to Cognitive Impairment in Symptomatic Carotid Artery Disease. Radiology. 2017;282:502-15.
[29] Appel J, Potter E, Bhatia N, Shen Q, Zhao W, Greig MT, et al. Association of white matter hyperintensity measurements on brain MR imaging with cognitive status, medial temporal atrophy, and cardiovascular risk factors. AJNR Am J Neuroradiol. 2009;30:1870-6.
[30] Park G, Hong J, Duffy BA, Lee JM, Kim H. White matter hyperintensities segmentation using the ensemble U-Net with multi-scale highlighting foregrounds. Neuroimage. 2021;237:118140.
[31] Oppedal K, Engan K, Eftestøl T, Beyer M, Aarsland D. Classifying Alzheimer's disease, Lewy body dementia, and normal controls using 3D texture analysis in magnetic resonance images. Biomedical Signal Processing and Control. 2017;33:19-29.
[32] Bordin V, Coluzzi D, Rivolta MW, Baselli G. Explainable AI Points to White Matter Hyperintensities for Alzheimer's Disease Identification: a Preliminary Study. 2022 44th Annual International Conference of the IEEE Engineering in Medicine & Biology Society (EMBC)2022. p. 484-7.
[33] Cajanus A, Hall A, Koikkalainen J, Solje E, Tolonen A, Urhemaa T, et al. Automatic MRI Quantifying Methods in Behavioral-Variant Frontotemporal Dementia Diagnosis. Dement Geriatr Cogn Dis Extra. 2018;8:51-9.
[34] Oppedal K, Eftestøl T, Engan K, Beyer MK, Aarsland D. Classifying dementia using local binary patterns from different regions in magnetic resonance images. Int J Biomed Imaging. 2015;2015:572567.
[35] Chen H, Sheng X, Qin R, Luo C, Li M, Liu R, et al. Aberrant White Matter Microstructure as a Potential Diagnostic Marker in Alzheimer's Disease by Automated Fiber Quantification. Front Neurosci. 2020;14:570123.
[36] Belathur Suresh M, Fischl B, Salat DH. Factors influencing accuracy of cortical thickness in the diagnosis of Alzheimer's disease. Hum Brain Mapp. 2018;39:1500-15.

[37] Provenzano FA, Muraskin J, Tosto G, Narkhede A, Wasserman BT, Griffith EY, et al. White Matter Hyperintensities and Cerebral Amyloidosis: Necessary and Sufficient for Clinical Expression of Alzheimer Disease? JAMA Neurology. 2013;70:455-61.
[38] Xie Y, Cui Z, Zhang Z, Sun Y, Sheng C, Li K, et al. Identification of Amnestic Mild Cognitive Impairment Using Multi-Modal Brain Features: A Combined Structural MRI and Diffusion Tensor Imaging Study. Journal of Alzheimer’s Disease. 2015;47:509-22.
[39] Li B, Zhang M, Riphagen J, Morrison Yochim K, Li B, Liu J, et al. Prediction of clinical and biomarker conformed Alzheimer’s disease and mild cognitive impairment from multi-feature brain structural MRI using age-correction from a large independent lifespan sample. NeuroImage: Clinical. 2020;28:102387.
[40] Zhang W, Li M, Zhou X, Huang C, Wan K, Li C, et al. Altered serum amyloid beta and cerebral perfusion and their associations with cognitive function in patients with subcortical ischemic vascular disease. Front Neurosci. 2022;16:993767.
[41] Zhang W, Zheng X, Li R, Liu M, Xiao W, Huang L, et al. Research on nonstroke dementia screening and cognitive function prediction model for older people based on brain atrophy characteristics. Brain Behav. 2022;12:e2726.
[42] Zhang Y, Tartaglia MC, Schuff N, Chiang GC, Ching C, Rosen HJ, et al. MRI Signatures of Brain Macrostructural Atrophy and Microstructural Degradation in Frontotemporal Lobar Degeneration Subtypes. Journal of Alzheimer’s Disease. 2013;33:431-44.
[43] Zhao J, Ding X, Du Y, Wang X, Men G. Functional connectivity between white matter and gray matter based on fMRI for Alzheimer's disease classification. Brain and Behavior. 2019;9:e01407.
[44] Schouten TM, Koini M, de Vos F, Seiler S, van der Grond J, Lechner A, et al. Combining anatomical, diffusion, and resting state functional magnetic resonance imaging for individual classification of mild and moderate Alzheimer's disease. NeuroImage: Clinical. 2016;11:46-51.
[45] Crystal O, Maralani PJ, Black S, Fischer C, Moody AR, Khademi A. Detecting conversion from mild cognitive impairment to Alzheimer's disease using FLAIR MRI biomarkers. Neuroimage Clin. 2023;40:103533.
[46] Chen Y, Lu P, Wu S, Yang J, Liu W, Zhang Z, et al. CD163-Mediated Small-Vessel Injury in Alzheimer’s Disease: An Exploration from Neuroimaging to Transcriptomics. Int J Mol Sci. 2024;25:2293.
[47] Shi Y, Deng J, Mao H, Han Y, Gao Q, Zeng S, et al. Macrophage Migration Inhibitory Factor as a Potential Plasma Biomarker of Cognitive Impairment in Cerebral Small Vessel Disease. ACS Omega. 2024;9:15339-49.
[48] Strain JF, Phuah C-L, Adeyemo B, Cheng K, Womack KB, McCarthy J, et al. White matter hyperintensity longitudinal morphometric analysis in association with Alzheimer disease. Alzheimer's & Dementia. 2023;19:4488-97.
[49] Zhu XW, Liu SB, Ji CH, Liu JJ, Huang C. Machine learning-based prediction of mild cognitive impairment among individuals with normal cognitive function. Front Neurol. 2024;15.
[50] De Francesco S, Crema C, Archetti D, Muscio C, Reid RI, Nigri A, et al. Differential diagnosis of neurodegenerative dementias with the explainable MRI based machine learning algorithm MUQUBIA. Sci Rep. 2023;13:17355.
[51] Feng J, Hui D, Zheng Q, Guo Y, Xia Y, Shi F, et al. Automatic detection of cognitive impairment in patients with white matter hyperintensity and causal analysis of related factors using artificial intelligence of MRI. Comput Biol Med. 2024;178:108684.
[52] Jokinen H, Koikkalainen J, Laakso HM, Melkas S, Nieminen T, Brander A, et al. Global Burden of Small Vessel Disease–Related Brain Changes on MRI Predicts Cognitive and Functional Decline. Stroke. 2020;51:170-8.
[53] Rabin JS, Neal TE, Nierle HE, Sikkes SAM, Buckley RF, Amariglio RE, et al. Multiple markers contribute to risk of progression from normal to mild cognitive impairment. NeuroImage: Clinical. 2020;28:102400.
[54] Stephan BC, Tzourio C, Auriacombe S, Amieva H, Dufouil C, Alpérovitch A, et al. Usefulness of data from magnetic resonance imaging to improve prediction of dementia: population based cohort study. Bmj. 2015;350:h2863.
[55] Tang L, Wu X, Liu H, Wu F, Song R, Zhang W, et al. Individualized Prediction of Early Alzheimer's Disease Based on Magnetic Resonance Imaging Radiomics, Clinical, and

Laboratory Examinations: A 60-Month Follow-Up Study. J Magn Reson Imaging. 2021;54:1647-57.
[56] Tozer DJ, Zeestraten E, Lawrence AJ, Barrick TR, Markus HS. Texture Analysis of T1-Weighted and Fluid-Attenuated Inversion Recovery Images Detects Abnormalities That Correlate With Cognitive Decline in Small Vessel Disease. Stroke. 2018;49:1656-61.
[57] Twait EL, Andaur Navarro CL, Gudnason V, Hu Y-H, Launer LJ, Geerlings MI. Dementia prediction in the general population using clinically accessible variables: a proof-of-concept study using machine learning. The AGES-Reykjavik study. BMC Medical Informatics and Decision Making. 2023;23:168.
[58] Verdelho A, Madureira S, Ferro JM, Baezner H, Blahak C, Poggesi A, et al. Physical activity prevents progression for cognitive impairment and vascular dementia: results from the LADIS (Leukoaraiosis and Disability) study. Stroke. 2012;43:3331-5.
[59] Wang J, Knol MJ, Tiulpin A, Dubost F, de Bruijne M, Vernooij MW, et al. Gray Matter Age Prediction as a Biomarker for Risk of Dementia. Proc Natl Acad Sci U S A. 2019;116:21213-8.
[60] Lambert C, Zeestraten E, Williams O, Benjamin P, Lawrence AJ, Morris RG, et al. Identifying preclinical vascular dementia in symptomatic small vessel disease using MRI. NeuroImage: Clinical. 2018;19:925-38.
[61] Liang L, Zhou P, Ye C, Yang Q, Ma T. Spatial–temporal patterns of brain disconnectome in Alzheimer's disease. Hum Brain Mapp. 2023;44:4272-86.
[62] Binzer M, Hammernik K, Rueckert D, Zimmer VA. Long-Term Cognitive Outcome Prediction in Stroke Patients Using Multi-task Learning on Imaging and Tabular Data. Cham: Springer Nature Switzerland; 2022. p. 137-48.
[63] Mortamais M, Reynes C, Brickman AM, Provenzano FA, Muraskin J, Portet F, et al. Spatial distribution of cerebral white matter lesions predicts progression to mild cognitive impairment and dementia. PLoS One. 2013;8:e56972.
[64] Aam S, Einstad MS, Munthe-Kaas R, Lydersen S, Ihle-Hansen H, Knapskog AB, et al. Post-stroke Cognitive Impairment-Impact of Follow-Up Time and Stroke Subtype on Severity and Cognitive Profile: The Nor-COAST Study. Front Neurol. 2020;11:699.
[65] Aamodt EB, Schellhorn T, Stage E, Sanjay AB, Logan PE, Svaldi DO, et al. Predicting the Emergence of Major Neurocognitive Disorder Within Three Months After a Stroke. Frontiers in Aging Neuroscience. 2021;13.
[66] Altieri M, Di Piero V, Pasquini M, Gasparini M, Vanacore N, Vicenzini E, et al. Delayed poststroke dementia. Neurology. 2004;62:2193-7.
[67] Peters F, Villeneuve S, Belleville S. Predicting Progression to Dementia in Elderly Subjects with Mild Cognitive Impairment Using Both Cognitive and Neuroimaging Predictors. Journal of Alzheimer’s Disease. 2014;38:307-18.
[68] West NA, Windham BG, Knopman DS, Shibata DK, Coker LH, Mosley TH, Jr. Neuroimaging findings in midlife and risk of late-life dementia over 20 years of follow-up. Neurology. 2019;92:e917-e23.
[69] Williams OA, Zeestraten EA, Benjamin P, Lambert C, Lawrence AJ, Mackinnon AD, et al. Predicting Dementia in Cerebral Small Vessel Disease Using an Automatic Diffusion Tensor Image Segmentation Technique. Stroke. 2019;50:2775-82.
[70] Yao M, Zhu YC, Soumaré A, Dufouil C, Mazoyer B, Tzourio C, et al. Hippocampal perivascular spaces are related to aging and blood pressure but not to cognition. Neurobiol Aging. 2014;35:2118-25.
[71] Rosano C, Aizenstein HJ, Wu M, Newman AB, Becker JT, Lopez OL, et al. Focal atrophy and cerebrovascular disease increase dementia risk among cognitively normal older adults. J Neuroimaging. 2007;17:148-55.
[72] Rosano C, Perera S, Inzitari M, Newman AB, Longstreth WT, Studenski S. Digit Symbol Substitution test and future clinical and subclinical disorders of cognition, mobility and mood in older adults. Age Ageing. 2016;45:688-95.
[73] Chen Y, Tozer D, Li R, Li H, Tuladhar A, De Leeuw FE, et al. Improved Dementia Prediction in Cerebral Small Vessel Disease Using Deep Learning–Derived Diffusion Scalar Maps From T1. Stroke. 2024;55:2254-63.

[74] Chen J, Yang J, Shen D, Wang X, Lin Z, Chen H, et al. A Predictive Model of the Progression to Alzheimer’s Disease in Patients with Mild Cognitive Impairment Based on the MRI Enlarged Perivascular Spaces. Journal of Alzheimer’s Disease. 2024;101:159-73.
[75] Keller JA, Sigurdsson S, Schmitz Abecassis B, Kant IMJ, Van Buchem MA, Launer LJ, et al. Identification of Distinct Brain MRI Phenotypes and Their Association With Long-Term Dementia Risk in Community-Dwelling Older Adults. Neurology. 2024;102:e209176.
[76] Lee M-W, Kim HW, Choe YS, Yang HS, Lee J, Lee H, et al. A multimodal machine learning model for predicting dementia conversion in Alzheimer’s disease. Scientific Reports. 2024;14:12276.
[77] Marzi C, Scheda R, Salvadori E, Giorgio A, De Stefano N, Poggesi A, et al. Fractal dimension of the cortical gray matter outweighs other brain MRI features as a predictor of transition to dementia in patients with mild cognitive impairment and leukoaraiosis. Frontiers in Human Neuroscience. 2023;17.
[78] Li R, Harshfield EL, Bell S, Burkhart M, Tuladhar AM, Hilal S, et al. Predicting incident dementia in cerebral small vessel disease: comparison of machine learning and traditional statistical models. Cerebral Circulation - Cognition and Behavior. 2023;5:100179.
